# A serum thyroid hormone metabolite test for molecular diagnostic evaluation of thyroid nodules

**DOI:** 10.64898/2026.09.25.26363992

**Authors:** Mengzhe Guo, Houfa Geng, Qiang Sun, Hong Wan, Ping Zhou, Yongfeng Song, Caiyan Zou, Xiu Zang, Hailin Xi, Tian Liao, Xuekui Liu, Gangshan Peng, Yu Wang, Mingzhao Xing, Jun Liang, “THMs Group”

## Abstract

This was a cohort study of 1,663 subjects (1196 women, 467 men; median age 48 years, IQR 37-57) with thyroid nodules recruited from 2020 to 2025 in five tertiary medical centers. Cytological/histopathological diagnoses were established by an independent review committee and preoperative serum thyroid hormone metabolites (**THMs**) were tested using liquid chromatography tandem mass spectrometry. A diagnostic model on this serum THM test to distinguish between differentiated thyroid cancer (**DTC**) and benign thyroid nodule (**BTN**) was trained on 872 retrospective subjects (454 DTC and 418 BTN), followed, prospectively, by model validation on 392 subjects (195 DTC and 197 BTN) and a second validation on 399 subjects (300 DTC and 99 BTN). Following successful diagnostic model training, the model validation demonstrated a diagnostic specificity of 99.5% (196/197), sensitivity of 92.3% (180/195), positive predictive value (**PPV**) of 99.4%, negative predictive value (**NPV**) of 92.9%, and diagnostic accuracy of 95.9% (95% CI, 90.8 to 100.0%) for DTC. Combination of the two validation cohorts showed a diagnostic specificity of 99.7% (295/296), sensitivity of 92.1% (456/495), PPV of 99.8%, NPV of 88.3%, and diagnostic accuracy of 94.9% (95% CI, 91.5 to 98.0%) for DTC. The test on indeterminate Bethesda categories III-IV (139 DTC, 87 BTN) showed a diagnostic specificity of 100.0% (87/87), sensitivity of 93.5% (130/139), PPV of 100.0%, NPV 90.6% and diagnostic accuracy of 96.0% (95% CI, 89.2 to 100.0%) for DTC. This study develops a novel molecular blood test of serum THMs for preoperative diagnostic evaluation of thyroid nodules, which displays a high PPV and decent NPV with an excellent overall diagnostic accuracy for DTC.

---

Dear Editor,

Thyroid cancer is a common endocrine malignancy. The most common histopathological type is papillary thyroid cancer (PTC), followed by follicular thyroid cancer (FTC) and oncocytic thyroid cancer (OCC). These cancers are collectively classified as differentiated thyroid cancer (DTC) and account for > 95% of all thyroid malignancies.^1,2^ The diagnosis of DTC typically begins with the evaluation of thyroid nodules, which occur in ∼60%–70% of the general population; 5%–10% of these nodules are DTCs, whereas the remainder are benign thyroid nodules (BTNs).^3,4^ The primary goal of the diagnostic evaluation of thyroid nodules is to distinguish malignant from benign nodules. The current diagnostic mainstays for thyroid nodules are ultrasonography and fine-needle aspiration biopsy (FNAB). For ultrasonography, the American College of Radiology Thyroid Imaging Reporting and Data System (ACR TI-RADS) is widely used to estimate the risk of malignancy. It comprises five incremental categories, ranging from 1 to 5, with categories 1–2 indicating benign/non-suspicious nodules and category 5 indicating nodules that are highly suspicious for malignancy.^5^ For FNAB, the Bethesda System for Reporting Thyroid Cytopathology (TBSRTC) is widely used; it comprises six diagnostic categories, each associated with an estimated risk of malignancy, with category I being non-diagnostic, II being benign, III being atypia of undetermined significance, IV being follicular neoplasm, V being suspicious for malignancy, and VI being malignant.^6^ However, these diagnostic modalities have limited accuracy, particularly for Bethesda categories III–IV, which encompass cytologically indeterminate thyroid nodules that occur in 25%–30% of cases and pose a major diagnostic challenge in current thyroid nodule management.^3,4,7^

Various molecular tests for thyroid diagnosis have been investigated, and the most commonly used are RNA- or DNA-based tests performed on FNAB specimens.^3,4,8^ These tests have improved the diagnostic accuracy of thyroid nodules, particularly as “rule-out” tests because of their high sensitivities and negative predictive values (NPVs) for malignancy. However, they share common limitations, with low specificity and positive predictive values (PPVs) being the most prominent limitations and hence have low “rule-in” power for malignancy. Moreover, they all require invasive FNAB to obtain specimens.^9,10^ Ideally, a noninvasive molecular test should have high specificity and decent sensitivity, thus providing excellent overall diagnostic accuracy.

The detection of serum thyroid hormone metabolites could potentially fulfill this need. Thyroid hormones, including thyroxine (T^4^), 3,5,5′-triiodothyronine (T^3^), and 3,3′,5′-triiodothyronine (rT^3^), are classical serum biomarkers of thyroid function but have no diagnostic value for thyroid cancer. Several well-characterized thyroid hormone metabolites (THMs), particularly those detectable in serum, include 3,5-diiodo-L-thyronine (3,5-T^2^); 3′, 5′-diiodo-L-thyronine (3′,5′-T^2^); 3,3′-diiodo-L-thyronine (3,3′-T^2^); 3’-iodo-L-thyronine (3′-T^1^); 3-iodo-L-thyronine (3-T^1^); L-thyronine (T^0^); and 3-iodothyronamine (3-T^1^AM), which are generated through the metabolic pathways of thyroid hormones.^11^ These THMs are produced through reactions catalyzed by various enzymes, including selenoenzymes such as deiodinase 3 (DIO3) and ornithine decarboxylase (ODC) (Supplementary information, Fig. S1). Interestingly, the expression of thyroid hormone-metabolizing enzymes, such as DIO3, is robustly increased in DTC tumors promoted by oncogenic signaling pathways compared with normal tissues.^12^

Like thyroid hormones, THMs are released into the blood, albeit at low concentrations, normally in the pg/mL range. The abundantly expressed thyroid hormone-metabolizing enzymes — particularly deiodinases — in DTC tumors are present in a local thyroid environment that is highly enriched in thyroid hormone substrates. This creates a unique condition that actively and robustly drives THM production, perhaps even in relatively small DTC tumors. Consequently, the presence of a DTC tumor may result in the continuous release of large amounts of THMs from this active production source into the bloodstream, significantly altering the serum THM concentrations that would otherwise remain low. This could make blood-based THM detection a sensitive diagnostic test for distinguishing DTC from BTN. We tested this hypothesis in the present double-blind clinical study using liquid chromatography-tandem mass spectrometry (LC-MS/MS), a sensitive and stable method commonly used to measure serum THMs,^13^ with the goal of establishing a novel molecular blood test for the preoperative diagnostic evaluation of thyroid nodules.

A logistic regression model (LRM) incorporating three diagnostically informative serum THMs — T^0^, 3-T^1^AM, and 3-T^1^ — was built and trained using the retrospective 872-sample dataset, validated using the prospective 392-sample dataset, and further validated using a second prospective 399-sample dataset for the diagnosis of DTC (Fig. 1a, b; Supplementary information, Fig. S2). Patient baseline characteristics, testing methods, and other information are presented in Supplementary information, Tables S1–S14 and S19. Model training yielded a diagnostic specificity of 87.1% (364/418), sensitivity of 93.0% (422/454), PPV of 88.7%, NPV of 91.9%, and diagnostic accuracy of 90.1% (95% CI, 87.0%–92.9%) for DTC. The modelvalidation demonstrated a diagnostic specificity of 99.5% (196/197), sensitivity of 92.3% (180/195), PPV of 99.4%, NPV of 92.9%, and diagnostic accuracy of 95.9% (95% CI, 90.8%–100%) for DTC. The second validation demonstrated a diagnostic specificity of 100% (99/99), sensitivity of 92.0% (276/300), PPV of 100%, NPV of 80.5%, and diagnostic accuracy of 94.0% (95% CI, 89.5%–97.6%) for DTC. Combining the two validation cohorts yielded a diagnostic specificity of 99.7% (295/296), sensitivity of 92.1% (456/495), PPV of 99.8%, NPV of 88.3%, and diagnostic accuracy of 94.9% (95% CI, 91.5%–98.0%) for DTC (Supplementary information, Table S15).

**Fig. 1.**
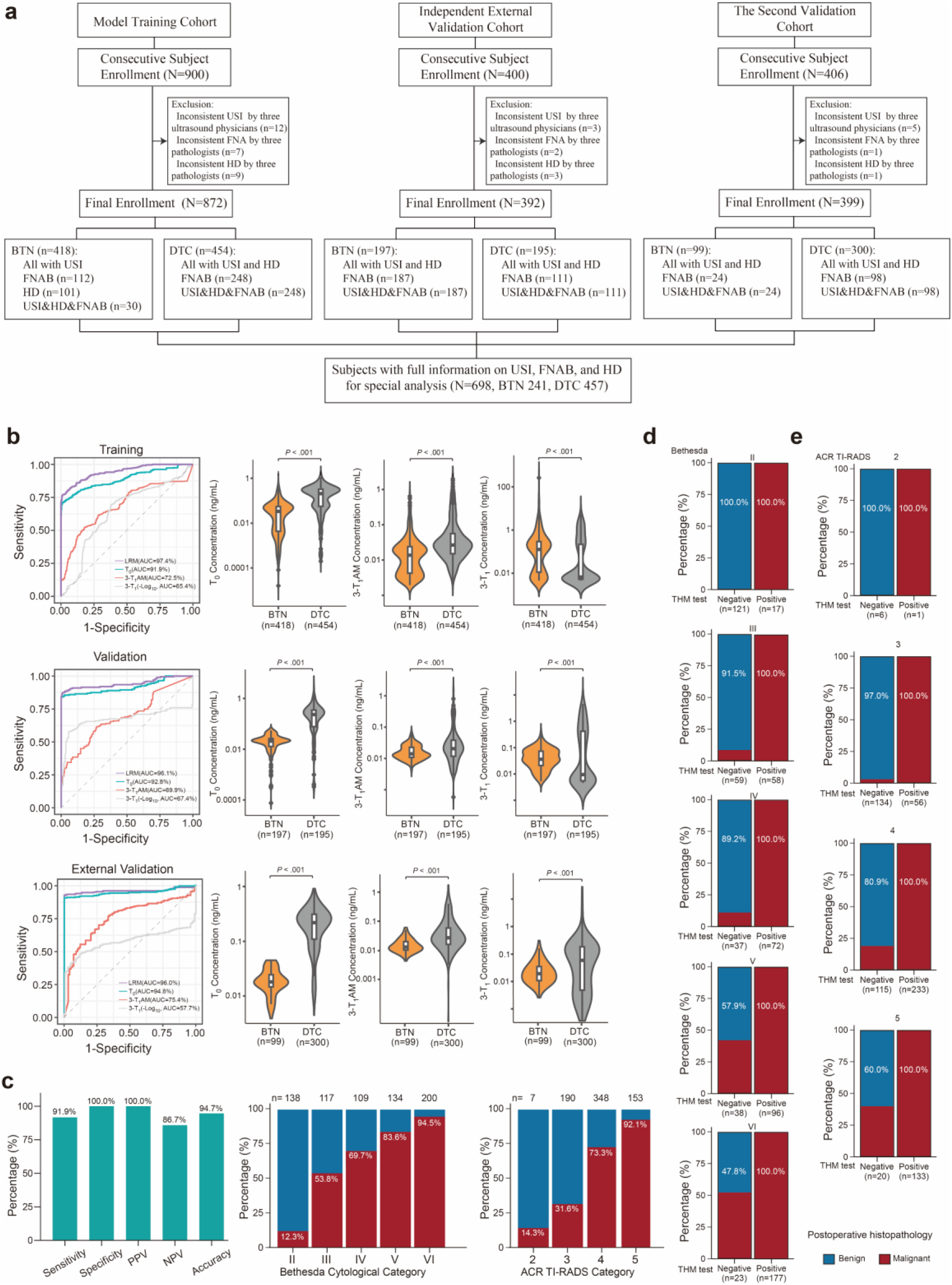
**a** Subject enrollment, study design, and workflow. **b** Differences in the levels of three thyroid hormone metabolites between subjects with DTC and those with BTN, and receiver operating characteristic (ROC) curves for the three metabolites and the LRM in the model training, independent external-validation, and second -validationcohorts. **c** Sensitivity, specificity, PPV, NPV, and accuracy, together with the distributions of histopathologically benign and malignant thyroid nodules across the Bethesda cytological and ACR TI-RADS categories. **d** Distribution of histopathologically benign and malignant thyroid nodules among THM model-negative and -positive Bethesda II – VI categories; **e** Distribution of histopathologically benign and malignant thyroid nodules among THM-negative and -positive ACR TI-RADS 2 – 5 categories. USI, ultrasound imaging; HD, histopathological diagnosis; TBSRTC, the Bethesda System for Reporting Thyroid Cytopathology.

We performed a subgroup analysis of 698 subjects with complete ultrasonographic, cytological, and histopathological diagnostic data (Fig. 1c, d). As expected, the histopathologically confirmed malignancy rates increased consistently from Bethesda II to VI. Among Bethesda II nodules, all THM test-negative cases (121/121; 100%) were histopathologically benign. For Bethesda III and IV nodules, 91.5% (54/59) and 89.2% (33/37) of THM test-negative cases were histopathologically benign, respectively. Among Bethesda V and VI nodules, 22 and 11 cases, respectively, were histopathologically confirmed as benign, and all tested negative on the THM test.

Supplementary information, Table S16 summarizes the diagnostic performance of the THM test in individual Bethesda categories. All (100%) THM test-positive cases were histopathologically malignant in all Bethesda categories. Overall, the THM test showed a diagnostic specificity of 100% (98.4%–100%), sensitivity of 91.9% (89.0%–94.1%), PPV of 100% (99.1%–100%), NPV of 86.7% (82.2%–90.2%), and diagnostic accuracy of 94.7% (91.1%–97.8%). For the 226 cases in the indeterminate Bethesda categories III and IV, the THM test showed a diagnostic specificity of 100% (95.8%–100%), sensitivity of 93.5% (88.2%–96.6%), PPV of 100% (97.1%–100%),NPV of 90.6% (83.1%–95.0%), and diagnostic accuracy of 96.0% (89.2%–100%).

In these 698 subjects with complete ultrasonographic, cytological, and histopathological diagnoses, increasing rates of histopathologically confirmed malignancy were observed from ACR TI-RADS categories 2–5 as expected (Fig. 1e). For ACR TI-RADS category 2 thyroid nodules, all THM test-negative cases (6/6; 100%) were histopathologically benign. For ACR TI-RADS categories 3 and 4, 97.0% (130/134) and 80.9% (93/115) of THM test-negative cases were histopathologically benign, respectively. For ACR TI-RADS category 5 thyroid nodules, 12 cases were histopathologically benign, all of which were negative on the THM test. Among THM test-positive cases, 100% were histopathologically malignant across all ACR TI-RADS categories (Supplementary information, Table S17).

There were 189 subjects with both DTC and HT; THM testing showed no effect of HT on the test results (Supplementary information, Fig. S3a). THM testing in 340 HV subjects showed a clear difference between HV and DTC subjects (Supplementary information, Fig. S3b). Tissue microarray immunohistochemical staining showed robust increases in DIO3 and ODC protein expression in DTC compared with adjacent normal tissues (*P* < 0.001 for DIO3 and *P* < 0.01 for ODC). DIO3 and ODC were both significantly increased in DTC compared with BTN (*P* = 0.005 and < 0.001, and *P* = 0.01 for ODC) (Supplementary information, Fig. S4). The concentrations of T^0^ and 3-T^1^AM were significantly higher in DTC and adjacent normal tissues than in BTN and adjacent normal tissues, whereas 3-T^1^ was correspondingly lower, as expected, in DTC and adjacent normal tissues than in BTN and adjacent tissues (Supplementary information, Fig. S5 and Table S18). T^0^ concentration was significantly positively correlated with DTC tumor size (Supplementary information, Fig. S6) and was positively correlated with 3-T^1^AM and negatively correlated with 3-T^1^, as expected (Supplementary information, Fig. S7). The three THMs did not differ significantly between patients with DTC with and without lymph node metastasis (Supplementary information, Fig. S8). There was no significant difference in urinary iodine content (UIC) among the DTC, BTN, and HV groups tested (Supplementary information, Fig. S9), and UIC may not alter serum THM levels unless it causes measurable dysfunction of the thyroid gland (Supplementary information, Fig. S10). The THM test would therefore be most reliable when performed in naturally euthyroid patients with normal iodine nutritional status.

This study developed a THM-based molecular test for thyroid nodules that consistently demonstrates, across clinical settings, high diagnostic specificity and PPV, reasonably high sensitivity and NPV, and an excellent overall diagnostic accuracy. Of particular note, this test performs well in indeterminate Bethesda III-IV thyroid nodules, making THM detection an excellent “rule-in” but suboptimal “rule-out” test for DTC^8-9^.

Unlike invasive FNAB-based molecular tests, THM detection is based on a convenient blood test. This approach provides the first preoperative diagnostic blood test for DTC. Given its nearly 100% PPV and “rule-in” power, a positive THM test result would be sufficient to diagnose DTC without the need for additional diagnostic measures. Given its suboptimal NPV, a negative THM test would largely rule out DTC, particularly in low Bethesda or ACR TI-RADS categories. In high Bethesda or ACR TI-RADS categories, which are much less common clinically, a negative THM test needs to be considered within the clinical context, and additional use of current FNAB-based molecular diagnostic tests with high NPV could provide complementary information.^3,4,7^

The small number of THMs required to achieve a high diagnostic efficiency makes this test economical. The ability of LC-MS/MS to sensitively detect low serum THM concentrations (pg/mL) was consistent with previous reports.^12^ The LC-MS/MS was also accurate and stable, as demonstrated by the remarkable consistency between clinical test results and LC-MS/MS measurements of T^4^ and T^3^, as well as among multiple interday THM measurements (Supplementary information, Tables S13 and S14). Although thyroid function parameters were all within normal ranges in this study, they were somewhat lower in patients with DTC than in those with BTN, consistent with previous reports.^13^ This may have led to an underestimation of the diagnostic sensitivity of serum THMs for DTC. Indeed, the diagnostic sensitivity of the test was enhanced when FT^4^ was adjusted for as a confounding factor in the LRM (Supplementary information, Fig. S11). The results regarding the relationship between expression of deiodinases in DTC and serum thyroid function parameters are presented in Supplementary information, Fig. S12.

It is worth noting that the concentrations of T^0^ and 3-T^1^AM in DTC tissues were three orders of magnitude higher than those in serum (Supplementary information, Fig. S5 and Table S18), consistent with abundant production of THMs by highly expressed thyroid hormone-metabolizing deiodinases in DTC tumors within the thyroid hormone substrate-enriched local environment of the thyroid gland. In addition to confirming previous reports of increased expression of DIO3 in DTC,^14,15^ the present study also observed markedly increased ODC expression compared with normal tissues, as well as markedly higher expression of both proteins in DTC than in BTN. As expected, we found an association between DTC tumor size and serum THMs, particularly T^0^, a late metabolite in the thyroid hormone metabolic pathway driven by several deiodinases (Supplementary information, Figs. S1, S6). We found no impact of lymph node metastases on serum THMs, as expected, since, unlike the thyroid hormone-enriched thyroid gland, lymph nodes are not rich in thyroid hormones that can serve as substrates for deiodinases. The THM test might not be suitable for individuals with inadequate or excessive iodine intake, but this was not an issue in the present study given the similar urinary iodine levels among the different patient groups. The test may also not be suitable for rare individuals with deiodinase gene mutations. We also recalculated the predictive values by applying cohort-specific sensitivity and specificity to assumed disease prevalence (Supplementary information, Table S20).

The current results may apply primarily to PTC given the relatively small number of FTC/OCC cases included in the study, even though the latter cases, like PTC, all tested positive on the THM test. Future studies including larger numbers of FTC/OCC cases would provide further validation of the test in these tumor types. Some BTN subjects in the retrospective model training cohort had BTN diagnoses based only on cytology or ultrasonography. It is likely that some malignant nodules existed in such “BTN” subjects, which could have caused “false positives” on the THM test, artificially reducing diagnostic specificity. Indeed, the performance of the THM test was superior in the two model validation cohorts, in which all cases of DTC and BTN had histopathologically confirmed diagnoses, compared with that in the model training cohort. The applicability of the THM test to rare types of thyroid cancer also remains to be investigated.

In summary, this study developed a novel blood test of serum THMs for the molecular diagnostic evaluation of thyroid nodules, which demonstrates excellent rule-in and less robust rule-out performance for DTC, particularly PTC. This is the first blood test for effective molecular diagnostic evaluation of thyroid nodules and accurate preoperative diagnosis of DTC, particularly PTC, with the potential to have a major clinical impact.

## Contributions

J. L., M.X. and Y.W. conceived and supervised the study. H.G., H.W., P.Z., Y.S., T.L., H.X. performed sample collection. M.G., X.Z., C.Z., and G.P. performed LC-MS analysis. M.G., Q.S., X.L. analyzed the data. M.G., H.G. drafted the manuscript. J.L. and M.X. edited the manuscript.

## Ethics approval

The study was registered in the Chinese Clinical Trial Registry (ChiCTR2300070912) and approved by the ethics committee of participating medical centers, including Xuzhou Central Hospital (XZXY-LJ-20200724-029), Xuzhou, Jiangsu; Liyang People’s Hospital (2023005), Changzhou, Jiangsu; First Affiliated Hospital of Anhui Medical University (PJ-YX2022-014F1), Hefei, Anhui; Fudan University Cancer Center (050432-4-2108*), Shanghai; and Shandong First Medical University Affiliated Endocrine and Metabolic Disease Hospital (EMDH202112001), Jinan, Shandong. All patients signed the informed consent form.

## Acknowledgements

This study was supported by the National Natural Science Foundation of China (82072951, 82172892, and 81973346), the Natural Science Foundation of Jiangsu Province (BK20220672), the Science and Technology Commission of Shanghai Municipality (22Y21900100), the Shanghai Anticancer Association (SACA-AX202213), Xuzhou Health Summit Team (2025DF07), Xuzhou “Peng Cheng Ying Cai” medical talent program (XWRCHT20220059), Xuzhou Municipal Science and Technology Bureau (KC21231, KC23145).

## Data availability

All data are available in the main text or supplementary information.

