## Supplemental Materials for "A serum thyroid hormone metabolite test for molecular diagnostic evaluation of thyroid nodules"

**of**

**Acknowledgement**

We acknowledge Drs. Wu Di and Zhu Jiefei at Xuzhou Central Hospital; Zhang Yan and Jin Anqi at Fudan University Shanghai Cancer Center as expert pathologists contributed to managing the cytopathological/histopathological aspects of the study. Drs. Lou Kexin and Lyu Nan at Xuzhou Central Hospital; Wan Xiaochun at Fudan University Shanghai Cancer Center as senior ultrasound radiologists on the independent central review committee contributed to blinded ACR TI-RADS review of all the thyroid ultrasound images used in this study.

**The THMs** (**T**hyroid **H**ormone **M**etabolites **S**tudy) Group dedicated to investigating the diagnostic potential and underlying mechanisms of thyroid hormone metabolites in thyroid cancer and related diseases, as well as conducting clinical validation and translation. Members include: Jun Liang, Mingzhao Xing, Huaidong Song, Yu Wang, Xiaohong Chen, Mengzhe Guo, Houfa Geng, Qiang Sun, Yongfeng Song, Hong Wan, Ping Zhou, Xuekui Liu, Caiyan Zou, Xiu Zang, Gangshan Peng, Hailin Xi, Kexin Lou.

**Data Supplement Methods**

Data Supplement Method 1: Subjects and Study Design

Data Supplement Method 2: Diagnostic Model Training, Validation, and a Second Validation

Data Supplement Method 3: Statistics

Data Supplement Method 4: Ultrasound examination

Data Supplement Method 5: Fine-needle aspiration

Data Supplement Method 6: The histopathological diagnostic methods

Data Supplement Method 7: Inclusion and exclusion criteria

Data Supplement Method 8: Patient and public involvement

Data Supplement Method 9: Materials

Data Supplement Method 10: Sample preparation method

Data Supplement Method 11: LC-MS/MS analysis method

Data Supplement Method 12: LC-MS/MS method validation

Data Supplement Method 13: Logistic regression model

Data Supplement Method 14: Immunohistochemical (**IHC**) staining for DIO3, ODC, and TG

Data Supplement Method 15: Test of $T_0$, 3-$T_1$AM, 3-$T_1$, and $T_4$ in differentiated thyroid cancer (**DTC**), benign thyroid nodules (**BTN**), and adjacent normal tissues

Data Supplement Method 16: Urine iodine test

Data Supplement Method 17: Prevalence-adjusted PPV and NPV for THM Testing

Data Supplement Method 18: Analysis of the correlation relationship between the expression of thyroid hormone metabolizing enzymes in DTC and serum $FT_4$, TSH, and $T_0$.

### 1. Subjects and Study Design

A total of 1,706 subjects were consecutively recruited from five tertiary medical centers in China from August, 2020 to May, 2025, from whom 1,663 subjects [1196 women, 467 men; median age 48 years, interquartile ranges (**IQR**) 37-57] were finally enrolled, including 872 retrospective subjects (454 DTC and 418 BTN) for logistic regression diagnostic model training, 392 prospective subjects (195 DTC and 197 BTN) for the independent external model validation, and another 399 prospective subjects (300 DTC and 99 BTN) for a second external validation.

The study was registered in the Chinese Clinical Trial Registry (registration number: ChiCTR2300070912) and approved by the ethics committee of participating medical centers (IRB#), including Xuzhou Central Hospital (XZXY-LJ-20200724-029), Xuzhou, Jiangsu Province; Liyang People's Hospital (2023005), Changzhou, Jiangsu Province; First Affiliated Hospital of Anhui Medical University (PJ-YX2022-014F1), Hefei, Anhui Province; Fudan University Cancer Center (050432-4-2108*), Shanghai; and Shandong First Medical University Affiliated Endocrine and Metabolic Disease Hospital (EMDH202112001), Jinan, Shandong Province (**Table S1**). All subjects provided informed written consent.

All the 949 DTC cases (including 922 PTC, 19 FTC and 8 OCC) in this study and all the 296 BTN cases in the prospective validation cohorts were histopathologically confirmed for the diagnosis. The study included a total of 714 BTN cases, including 128 follicular thyroid adenoma (formally named solitary adenoma), 264 thyroid follicular nodular disease (formally named multinodular goiter), 2 oncocytic thyroid adenoma, and 3 Hashimoto's thyroiditis (**HT**). The 418 BTN cases in the retrospective model training cohort included 101 confirmed histopathologically, 112 by FNAB, and 205 in ACR TI-RADS categories 1 or 2 clinically treated as benign.[1] A group of 698 cases (457 DTC and 241 BTN) with complete information on ultrasonographic, FNAB, and histopathological diagnoses were used for analysis. Thyroid cytopathological/histopathological diagnoses were made by an independent central review committee of three senior pathologists according to the WHO criteria[2] as described in the **Table S2-6**. ACR TI-RADS diagnoses were made by an independent central review committee of three senior ultrasound radiologists (**Table S7, S8**). Demographic characteristics of the subjects are presented in **Table S9** and **S19**.

A tissue microarray was built using 47 DTC and 22 BTN tumors with paired adjacent normal tissues for immunohistochemical assessment of DIO3 and ODC protein expression. Urine iodine was examined in the cohort used for the THM model validation, in whom urine specimens were available. We also tested 340 randomly recruited healthy volunteers (**HV**) (**Table S10**).

The study was conducted in a double-blinded manner: $T_4$; $T_3$; $rT_3$; $T_0$; 3-$T_1$; 3'-$T_1$; 3,5-$T_2$; 3,3'-$T_2$; 3',5'-$T_2$ and 3-$T_1$AM were detected using LC-MS/MS by team members blinded to the clinical diagnosis; the diagnosing pathologists and ultrasound radiologists were blinded to the LC-MS/MS test results. The methodology is described in **Tables S11** and **S12** and validated for precision and stability in **Tables S13** and **S14**. Routine clinical test results of $T_4$ and $T_3$ were available in all subjects and were used to compare with the detection results of LC-MS/MS tests.

**2. Diagnostic Model Training, Validation, and a Second Validation**

Among the seven common serum THMs tested, $T_0$, 3-$T_1$AM, and 3-$T_1$ differed significantly between DTC and BTN (**Fig. 1 and Fig. S2**) and were selected for the diagnostic model training and validation. The model training and building were performed on 872 retrospective subjects, followed by the external model validation on 392 prospective subjects and a second validation on another 399 prospective subjects.

**3. Statistics**

Continuous data were summarized using medians and IQR for non-normally distributed variables or mean ± SD for normally distributed variables; categorical data were summarized using frequencies and percentages. Wilcoxon-Mann-Whitney test was used for non-normally distributed continuous variables and independent *t* test was used for normally distributed continuous variables. Categorical variables were compared using $\chi^2$ test. The logistic regression was performed to build the prediction model discriminating DTC from BTN on the training dataset, followed by test on the first and second validation datasets. Computed area under the curve (**AUC**) values were generated using the ROCR package in R for the evaluation of the discriminating performance of THMs-based model. [3] Specificity, sensitivity, PPV, NPV, diagnostic accuracy, and their 95% confidence intervals (**95% CI**) were calculated using standard statistical methods. When the actual value was equal to 0 or 100%, the 95% CI was calculated using the "exact method". [4] Hypothesis testing was done in a two-sided manner, with $P<0.05$ considered to be significant. The R statistical software (version 4.1.3) was used for the statistical analyses.

**4. Ultrasound examination**

Ultrasound examinations were performed by experienced radiologists specialized in thyroid imaging using 5-12 MHz linear-array probes (iU22, Philips Healthcare, Bothell, WA, USA; Logic 9, GE Healthcare, Wauwatosa, WI, the USA). The study documented key ultrasound characteristics indicative of the risk level of thyroid malignancy according to the ACR TI-RADS classification system, [1] including, for example, echogenicity, compositions, calcification states, margin characteristics, and shapes. Each ultrasound feature for an individual nodule was assigned points. All thyroid images underwent review by a central committee consisting of three senior ultrasound radiologists to confirm the diagnosis. Each radiologist independently evaluated the ultrasonography results. In case of disagreement among the radiologists, the decision was made based on the majority consensus or the case was excluded if all three pathologists differed in opinion.

**5. Thyroid fine needle aspiration biopsy and cytological diagnosis**

Fine needle aspiration biopsy (FNAB) was performed mainly on nodules with ACR TI-RADS categories 3 to 5 and other clinical risk indications. FNABs were carried out by experienced radiologists using ultrasound guidance. The FNAB specimen was smeared onto glass slides and fixed with 95% alcohol for cytopathology analysis. Pathologists analyzed the FNAB specimens and reported the results according to the Bethesda cytology classification system. [5] The treatment for patients with Bethesda II thyroid nodules followed the American Association of Endocrine Surgeons Guidelines, which suggest that observation is safe and surgery may be considered for symptomatic cases, such as those with local compressive symptoms caused by a large nodule, or upon the patient's preference. [6] All cytological results of FNAB were reviewed by a central committee consisting of three senior pathologists to confirm the diagnosis. Each pathologist independently evaluated the FNAB results. In case of disagreement among the pathologists, the decision was made based on the majority consensus or the case was excluded if all three pathologists differed in opinion.

**6. Thyroid histopathological diagnosis**

Thyroid histopathological diagnoses were made according to the definitions and criteria of the recent WHO classification. [2] Slides of formalin-fixed and paraffin-embedded thyroid histopathological specimens were H&E stained following standard procedures and reviewed by a central committee consisting of three senior pathologists. Each pathologist independently evaluated each case. In case of disagreement among the pathologists, the finalized results were determined by the majority consensus. The case was excluded if the three pathologists differed in opinion.

**7. Inclusion and Exclusion criteria**

Subjects with thyroid nodules at age 18-85 years, who did not have the following conditions were included in this study:

(1) Present/past history of thyroid dysfunction; (2) A history of chemoradiotherapy; (3) Present or past history of non-thyroid malignancy; (4) A history of surgery or blood transfusion in the past six months; (5) Significant other organ diseases; (6) Metabolic

diseases, such as diabetes mellitus; (7) Immune system diseases, such as lupus erythematosus; (8) Other major medical conditions; (9) A history of thyroid surgery; and (10) Currently taking thyroid hormone medication.

Healthy volunteers (HV) had normal thyroid function without thyroid nodules on ultrasonography. They also met the above inclusion and exclusion criteria.

**8. Patient and public involvement**

This research was conducted involving recruitment of patients who were fully informed of the current clinical problem to be addressed by the study, the nature, means, purpose, expectations, and other aspects of the study. With understanding of these and other research-related aspects as well as their enthusiastic interest and intention to help advance medicine, patients signed informed consent to participate and get involved in this study. They were actively involved in the research also by providing or allowing the use of their biospecimens (e.g., blood specimens, thyroid tumor samples, urine specimens, etc) and medical records. They were also involved in and supported this research by cooperating with the research team on follow-up activities. The study was registered in the Chinese Clinical Trial Registry (registration number: ChiCTR2300070912) and publicly posted. The information on the nature, purpose, goal, and other aspects of the program was publicized widely among patients, research team members, hospital staff, management leaders, family members, hospital visitors, post readers, and other interested people. Therefore, this research has received good public awareness and involvement.

**9. Materials**

Ascorbic acid, dithiothreitol, and citric acid were purchased from Sun Chemical Reagent Co., Ltd (Shanghai, China) and were each prepared into a solution with the concentration of 25 mg/mL. L-thyroxine ($T_4$) was purchased from Sun Chemical Reagent Co., Ltd (Shanghai, China). 3, 3', 5-triiodine-l-thyronine ($T_3$) and 3, 3', 5'-triiodine-l-thyronine ($rT_3$) were purchased from Sigma-Aldrich (Merk, the USA). 3, 5-

diiodine-l-thyronine (3, 5-$T_2$); 3', 5'-diiodine-l-thyronine (3', 5'-$T_2$); 3, 3'-diiodine-l-thyronine (3, 3'-$T_2$); 3'-iodo-L-thyronine (3'-$T_1$); 3-iodo-L-thyronine (3-$T_1$); L-thyronine ($T_0$); 3-iodothyronamine (3-$T_1$AM); and 3, 3', 5-triiodine-L-thyroxine-$^{13}C_6$ isotope were purchased from Toronto Research Chemicals (Toronto, Canada).

**10. Sample Preparation**

One mL of serum was added with 40 μL of each ascorbic acid, dithiothreitol, and citric acid solution (25 mg/mL). Ten ng of $^{13}$C-$T_3$ was added as the internal standard, followed by addition of 6 mL of prechilled methanol/acetonitrile (50/50, *v/v*) to mix with the serum. The mixture solution was placed in the ice bath for 20 min for protein precipitation, followed by centrifugation at 20,000 g for 10 min, and the supernatant was collected and vacuum frozen-dried. One mL of prechilled ethyl acetate with 3% formic acid was then added and the mixture solution was centrifuged at 20,000 g for 10 min for further removal of substrate. The supernatant was collected and vacuum frozen-dried. The dried sample was re-dissolved in 1 mL distilled water and enriched by HLB C18 SPE column (30 mg HLB C18 in 1-mL column, Waters, the USA), and 1 mL methanol with 1% formic acid was used as eluate. The eluted solution was dried and re-dissolved with 70 μL of initial mobile phase (methanol/water, 2/3, *v/v,* with 0.1% formic acid) for analysis.

**11. LC-MS/MS Analysis**

The LC-MS/MS analysis was performed by LC-30 ultra-performance liquid chromatography (Shimadzu, Japan) coupled with AB 5500 triple quadrupole tandem mass spectrometry (AB Sciex, the USA). The column used was the ACQUITY UPLC BEH C18 column (2.1 mm×100 mm, 1.7 μm, Waters, the USA), and the column temperature was 40 °C for LC-MS/MS analysis. The compounds were separated at a flow rate of 0.3 mL/min. Two types of mobile phase were used, containing 0.1% formic acid in water (*v/v*) as phase A and 0.1% formic acid in methanol (*v/v*) as mobile phase

B, respectively. A 12-min gradient was set as follows: the initial gradient was 60% mobile phase A and 40% mobile phase B; 0-4 min, the mobile phase was from 60% to 55%, and mobile phase B was from 40% to 45%; 4-7 min, mobile phase A and B remained at 55% and 45%, respectively; 7-9.8 min, the mobile phase was from 55% to 5%, and mobile phase B was from 45% to 95%; 9.8-11 min, mobile phase A and B remained at 5% and 95%, respectively; the post time was 1 min. The compounds were detected by MRM mode (the precursor and fragment ions of each compound can be seen in **Table S11**) *via* electron spray ion source (ESI). The de-clustering potential, entrance potential, and collision outlet potential were 50.0, 10.0, and 8.0 eV, respectively. The positive ion mode was used with a dry temperature of 500 °C and dry gas of 40 L/min. The 3, 3', 5-triiodine-L-thyroxine-$^{13}C_6$ isotope was used as the internal standard.

Before the detection of real samples, standard curves with detection range and lower limit of quantification of the ten substances were created. As shown in **Table S12**, the linearity of the standard curves was excellent with the range higher than three orders of magnitude. Moreover, the lower limit of quantification must be lower than the actual concentration of the substances in the samples.

**12. LC-MS/MS method validation**

The accuracy of the LC-MS/MS method was assessed in accordance with the U.S. food and drug administration (FDA) acceptance criteria for bio-analytical method validation guidance for industry. [7] Ten repetitions of low, medium, and high concentrations of each standard THM within the range of standard curve were detected on different days to evaluate the accuracy and inter-day precision, as per guidelines. The specific concentrations chosen for each THM are detailed in **Table S13**. Low, medium, and high concentration of T4 were chosen as 50, 100, and 150 ng/mL, respectively, to meet the ranges of their high serum concentrations. As shown in **Table S13**, inter-day precision analysis revealed excellent results for each standard THM, with most of the RSD values below 5% for concentrations under 1 pg/mL and even less than 2% for concentrations exceeding 1 ng/mL (**Table S13**). Moreover, repeated test of serum samples from DTC patients demonstrated good reproducibility for each THM, confirming the stability and accuracy of our LC-MS/MS method (**Table S14**)

### 13. Logistic regression model

The univariate logistic regression was performed to determine the association between individual THMs and DTC. After conducting collinearity analysis, the THMs that exhibited multicollinearity were removed and the remaining THMs were included in the multivariate logistic regression. The final equation comprised three THMs: $T_0$, 3-$T_1$AM, and 3-$T_1$. Therefore, a three-THM model of $T_0$, 3-$T_1$AM, and 3-$T_1$ was considered as the best-fitting parsimonious model, [8] yielding the following diagnostic signature:

**Factor (prediction) = -4.72 + 58.70 × $T_0$ + 39.45 × 3-$T_1$AM - 0.81 × 3-$T_1$.**

In the equation, -4.72 was the intercept; 58.70, 39.45, and -0.81 were the slopes (coefficients) for $T_0$, 3-$T_1$AM, and 3-$T_1$, respectively, in the best-fitting logistic-regression model. "$T_0$, 3-$T_1$AM, and 3-$T_1$" represent the actual concentrations (ng/mL) detected. "Factor" represents the collection of the three THMs. When the AUC of this logistic-regression model was maximized (*i.e.*, the value of sensitivity plus specificity was the greatest), the calculated cutoff value (Factor) was 0.346, with which the model had the best fitted sensitivity and specificity. A "Factor" > 0.346 was considered positive for DTC.

### 14. Immunohistochemical (IHC) staining for DIO3, ODC, and TG

Forty-seven DTC and 22 BTN tumors, along with matched adjacent normal tissues, were formalin-fixed and paraffin-embedded to produce tissue microarrays. The expression of deiodinase 3 (DIO3), ornithine decarboxylase (ODC), and thyroglobulin (TG) was assessed by IHC in these tissue microarrays. The procedures included deparaffinization, rehydration, antigen retrieval, endogenous peroxidase inactivation, and blocking of nonspecific reactions. Primary antibodies against DIO3, ODC, and TG (all purchased in Abcam, USA) were incubated overnight at a dilution of 1:100 for DIO3, 1:50 for ODC and 1:200 for TG at 4 ºC, followed by incubation with biotinylated secondary antibody (Roche, Germany). Slides were examined using an Olympus BX51 microscope equipped with an Olympus QColor 5 camera. Image capture was performed using the QCapturePro software. Immunoreactivity scores were based on the staining degree (color intensity) and staining extent (% of cells stained). Specifically, staining degree scores were assigned as follows: no staining = 0; light yellow = 1; brown = 2; and dark brown = 3. Staining extent scores were assigned as follows: 0-25% = 0 points; 26-50% = 1 point; 51-75% = 2 points; 76-100% = 3 points.

**15. Test of $T_0$, 3-$T_1$AM, 3-$T_1$, and T4 in DTC, BTN, and adjacent normal tissues**

Twenty-eight frozen DTC and twelve frozen BTN tumors, along with matched adjacent normal tissues, weighing approximately 100 mg each, were homogenized and subjected to THMs extraction using a 2 mL of acetonitrile solution. The precise tissue weights were provided in **Table S18**. The expression of $T_0$, 3-$T_1$AM, 3-$T_1$, and $T_4$ in tissues was analyzed by the same LC-MS/MS method employed for serum samples. As shown in **Fig. S5**, the concentration of $T_4$ in thyroid tissues aligned with previous literatures has no difference among DTC, BTN, and adjacent normal tissues. [9] However, the concentrations of $T_0$ and 3-$T_1$AM were significantly increased in DTC tissues, while those of 3-$T_1$ significantly were decreased in DTC and adjacent normal tissues, compared with BTN. This pattern mirrored that seen in serum samples.

The concentrations of $T_0$ and 3-$T_1$AM in DTC and adjacent normal tissues were about 10-1000 ng/g, which were three orders of magnitude higher than those found in serum (about 10-1000 pg/mL, seen in **Fig. S5** and **Table S18**), providing a plausible explanation for sufficient release of THMs from DTC to serum to effectively change the concentrations of THMs in the serum.

### 16. Urine iodine test

Urine iodine concentrations (UIC) were tested in a group of prospectively recruited subjects where urine specimens were also collected, including 150 DTC, 137 BTN, and 100 healthy volunteers (**HV**) as defined above. Urine iodine was analyzed by Urine Iodine Test Kit (Arsenic-cerium Catalytic Contact, Changsha Silky-Road Medical Technology Co., Ltd, China), coupled with automatic iodine analyzer (Changsha Silky-Road Medical Technology Co., Ltd, China). In brief, 5 mL of 24-hour collected urine sample was added to 55 mL of reaction solution containing $H_3AsO_3$ and $Ce^{4+}$. The iodine in urine can catalyze the reaction between $H_3AsO_3$ and $Ce^{4+}$ (the reaction equation was as follows), making $Ce^{4+}$ (yellow color) to $Ce^{3+}$ (colorless). The urine iodine concentration was calculated according to the time of the color change by the automatic iodine analyzer.

$$H_3AsO_3 + 2Ce^{4+} + H_2O \xrightarrow{I} H_3AsO_4 + 2Ce^{3+} + 2H^+$$

The UIC of subjects with DTC, BTN, and HV were 188.24 ± 21.37, 241.68±20.01, and 217.05±29.87, respectively. There was no difference in UIC among these three groups (**Fig. S9**).

We also performed additional analyses to address this issue more directly, including Sensitivity analysis of UIC for THM diagnosis, evaluation of THM diagnostic performance after individualized UIC correction, correlation analysis between UIC and thyroid function parameters $T_4$, $T_3$, and TSH, and analysis of the relationship between UIC and $T_0$, $3T_1AM$, and $3'T_1$.

Firstly, we assessed the sensitivity of our findings to potential unmeasured confounding through two complementary approaches. We compared unadjusted and urinary iodine-adjusted logistic regression models (since the UIC range varies from 10 to 2000, we performed a $\log_{10}$ transformation here). for each biomarker using E-value sensitivity analysis); stable effect estimates after covariate adjustment suggest that the observed associations are not driven by this key confounder. Then, we calculated E-values as a supplementary quantitative metric. The E-value represents the minimum strength of association, on the odds ratio scale, that an unmeasured confounder would need to have with both the exposure and the outcome to fully explain away the observed association, conditional on measured covariates [10]. For the PTC vs. BTN comparison, the UI-adjusted ORs for $T_0$ and $3T_1AM$ were 2.23 (95% CI: 1.30–3.81) and 2.71 (95% CI: 1.35–5.42), yielding E-values of 3.88 and 4.86 for the point estimates and 1.93 and 2.04 for the confidence interval lower bounds, respectively. These E-values indicate that an unmeasured confounder would need to be associated with both the biomarker and case status by an OR of at least ~2-fold to nullify the observed effects. While we report E-values for transparency, our primary sensitivity analysis relies on the consistency of estimates across nested multivariable models. (**Fig. S10A**).

Furthermore, we adjusted the individualized UIC by dividing the THM values by their corresponding $\log_{10}$(UIC) values and performed between-group analysis on the resulting data. After adjusting for UIC, $T_0$ and $3T_1AM$ remained significantly higher in the PTC group compared to the BTN group, while $3'T_1$ changed from being lower than the BTN group to showing no significant difference (**Fig. S10B**). This suggests that UIC values may not affect the diagnostic utility of $T_0$ and $3T_1AM$.

Furthermore, we performed correlation analyses between UIC and $FT_4$, $FT_3$, TSH as well as THMs. No significant correlations were observed between UIC and $FT_4$ or $FT_3$. A weak negative correlation was detected between UIC and TSH, yet the corresponding P value was greater than 0.05. Meanwhile, UIC showed no statistically significant correlation with THMs (**Fig. S10C**).

### 17: Prevalence-adjusted PPV and NPV for THM Testing

As malignancy-enriched cohorts could potentially affect the observed PPV and NPV, we calculated prevalence-adjusted predictive values using cohort-specific sensitivity and specificity. Specifically, we recalculated the predictive values by applying cohort-specific sensitivity and specificity to assumed disease prevalence using following formulas:

**Adjusted PPV = Se×P / [Se×P + (1 - Sp) ×(1 - P)]**

**Adjusted NPV = Sp×(1 - P) / [(1 - Se) ×P + Sp×(1 - P)]**

P denotes assumed disease prevalence; Se denotes sensitivity; Sp denotes specificity. And the recalculated values can be seen in **Table S20**.

Using the general-population incidence of thyroid cancer (49 per 100,000), the adjusted NPV was approximately 99.996% across cohorts, whereas the adjusted PPV varied substantially because PPV is highly prevalence- and specificity-dependent. Because THM testing is intended for patients with thyroid nodules rather than population-wide screening, we also estimated adjusted predictive values using a thyroid-nodule malignancy prevalence of 5%-15%.

### 18: Analysis of the correlation relationship between the expression of thyroid hormone metabolizing enzymes in DTC and serum $FT_4$, TSH, and $T_0$.

We examined the relationship between the thyroid hormone-metabolizing enzyme (deiodinases) expression obtained from immunohistochemistry and corresponding patients' thyroid function (including $FT_4$ levels and TSH) and $T_0$. The results are shown in **Fig. S12**. In DTC patients, deiodinase activities showed a trend of positive correlation with TSH and negative correlation with $FT_4$, although neither correlation reached statistical significance, consistent with our baseline data. However, deiodinase activity showed a significant positive correlation with $T_0$ in all cases of DTC, whereas in BTN patients there was no significant association between deiodinase activity and $T_0$. This explains why $T_0$ levels were higher in thyroid cancer patients compared to healthy individuals and BTN patients.

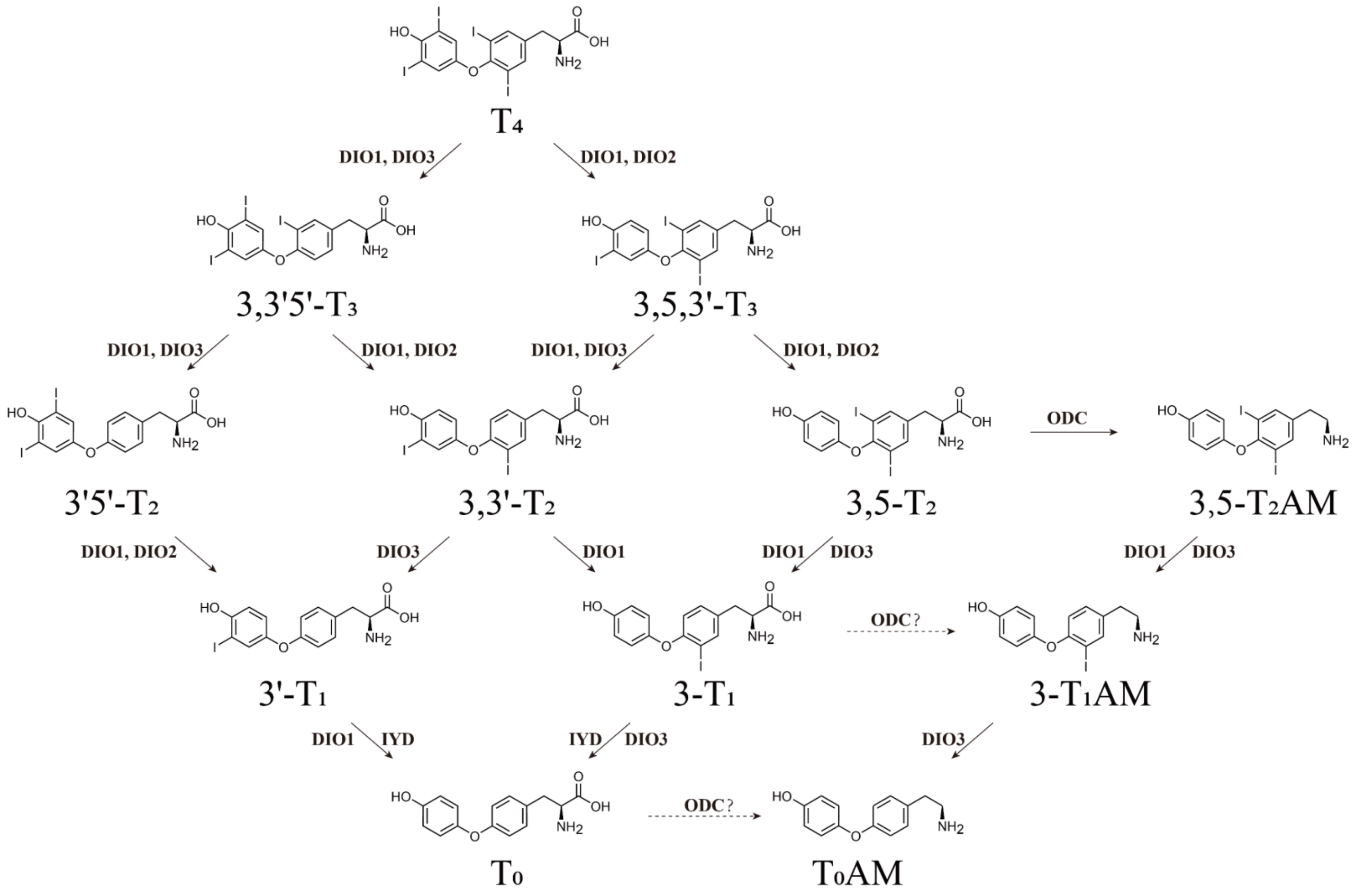

**Figure S1.** Thyroid hormone metabolites in their metabolic pathway and the deiodinases. [11,12]

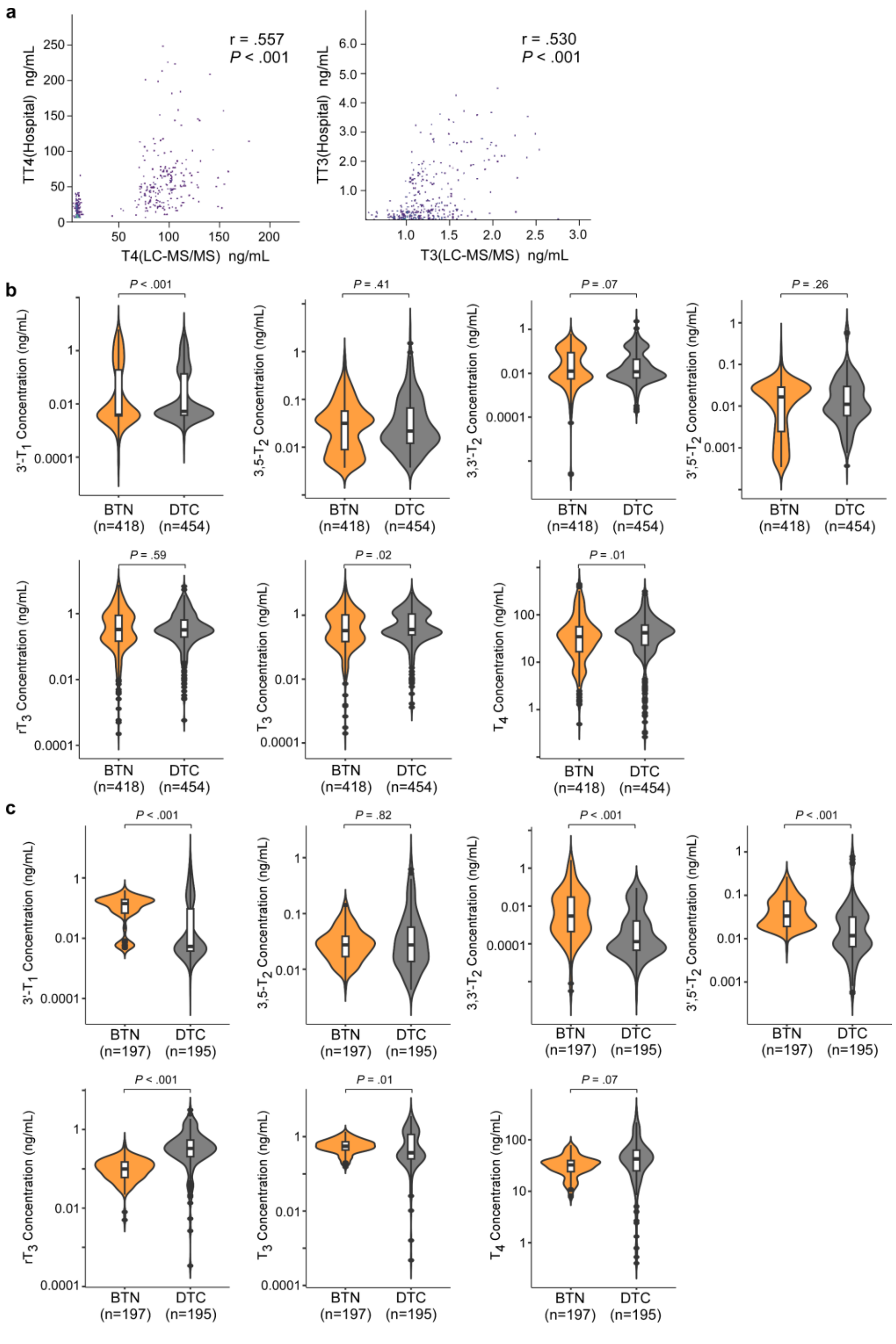

**Figure S2. Differences in serum $T_4$, $T_3$, $rT_3$ and other THMs between DTC and BTN.** (a) The correlation of $T_4$ and $T_3$ test results between the clinical method and the LC-MS method; (b-c) Comparison of thyroid hormones and their metabolite levels between DTC and BTN groups in training group (b) and validation group (c). DTC, Differentiated thyroid cancer; BTN, benign thyroid nodule.

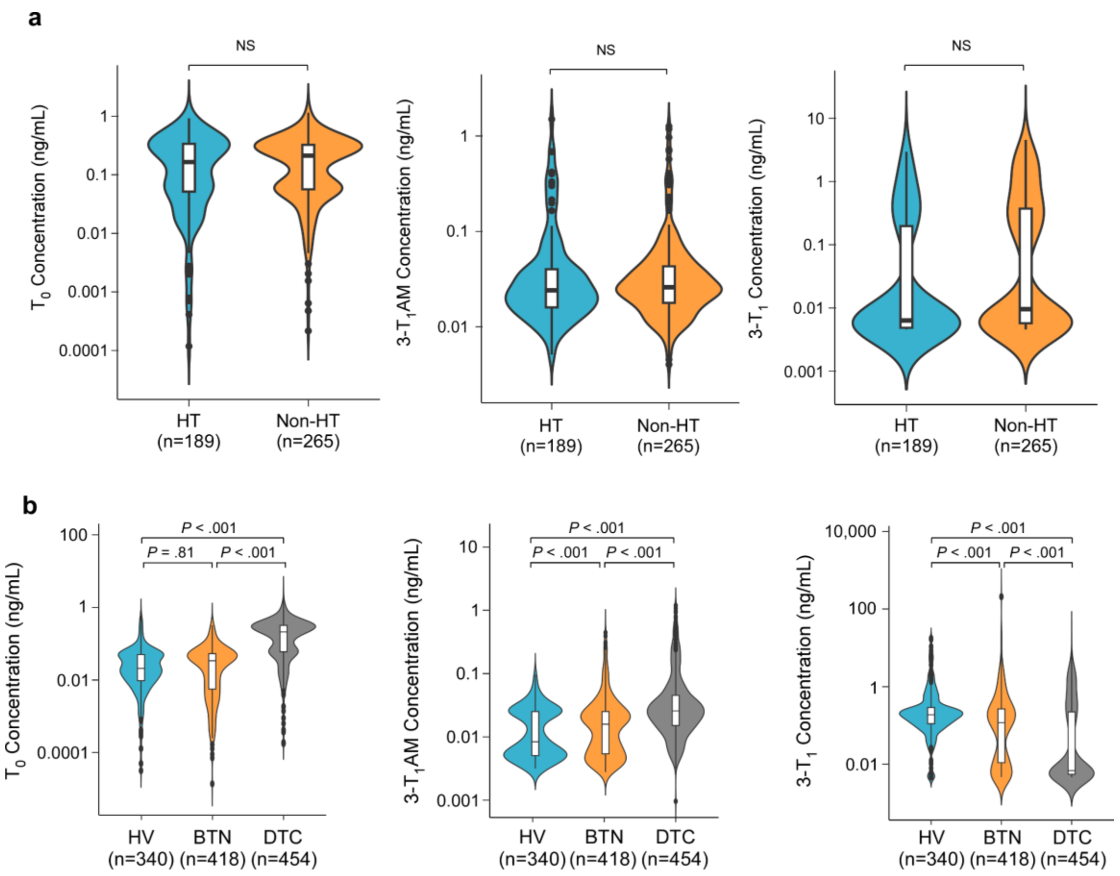

**Figure S3. Serum THM test in subjects with Hashimoto's thyroiditis (HT) and in healthy volunteers (HV).** (a) Comparison of the levels of $T_0$, 3-$T_1$AM, and 3-$T_1$ between individuals with HT and those without HT in DTC patients; (b) Comparison of $T_0$, 3-$T_1$AM, and 3-$T_1$ among the DTC, BTN and HV groups. HT, Hashimoto's thyroiditis; HV, healthy volunteers; DTC, differentiated thyroid cancer; BTN, benign thyroid nodule.

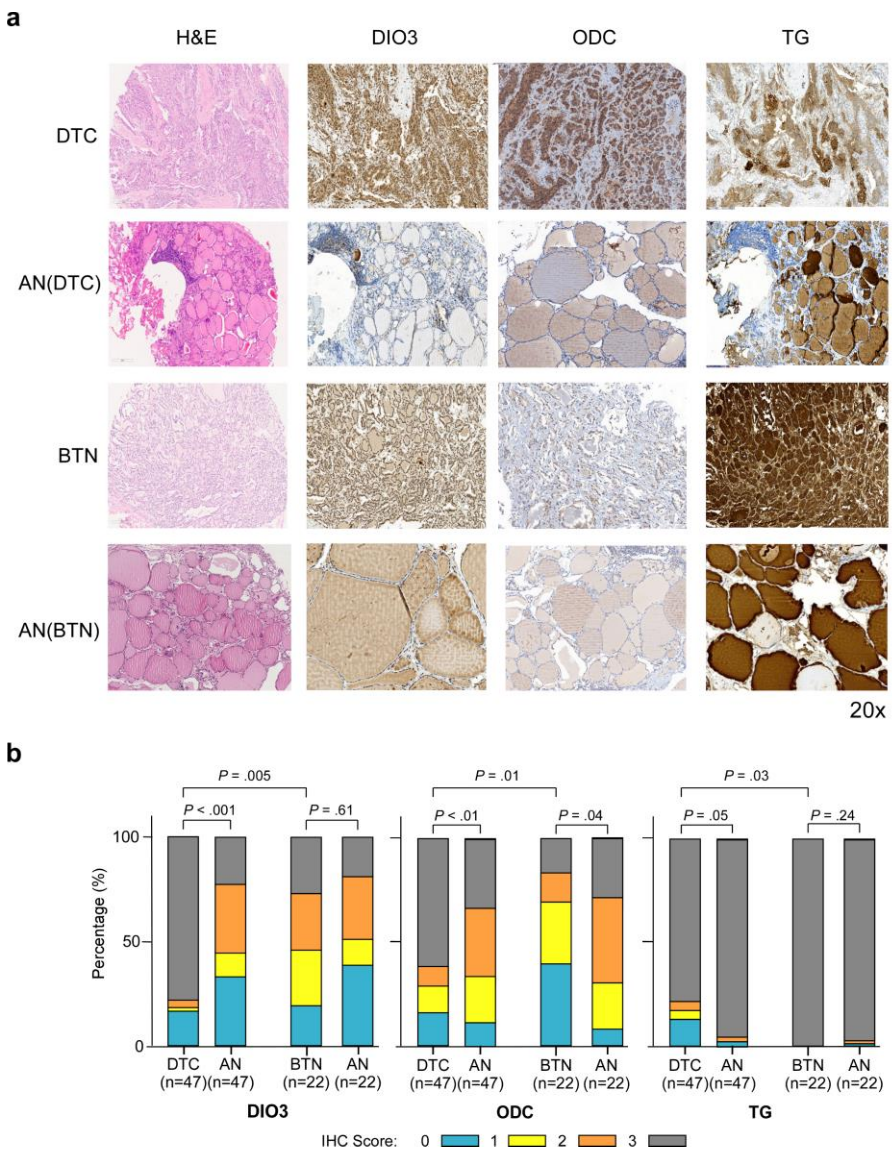

**Figure S4. Tumor microarray immunohistostaining of thyroid hormone-metabolizing enzymes and TG.** (A) The immunohistochemical analysis results for DIO3, ODC, and TG using tissue microarrays, consisting of 47 DTC tissues, 47 matched adjacent normal tissues, and 22 BTN tissues. (B) The IHC score of DIO3, ODC, and TG in adjacent normal, DTC, and BTN tissues. The average gray value (staining degree/intensity) and percentage of positive cells (staining extent) were used as IHC measurement scores by Image. Four ratings were given: High positive (3), Positive (2), Low Positive (1), and Negative (0). DTC, differentiated thyroid cancer; BTN, benign thyroid nodule; AN, adjacent normal tissues; DIO3, deiodinases 3; ODC, ornithine decarboxylase; TG, thyroglobulin; IHC, immunohistochemistry.

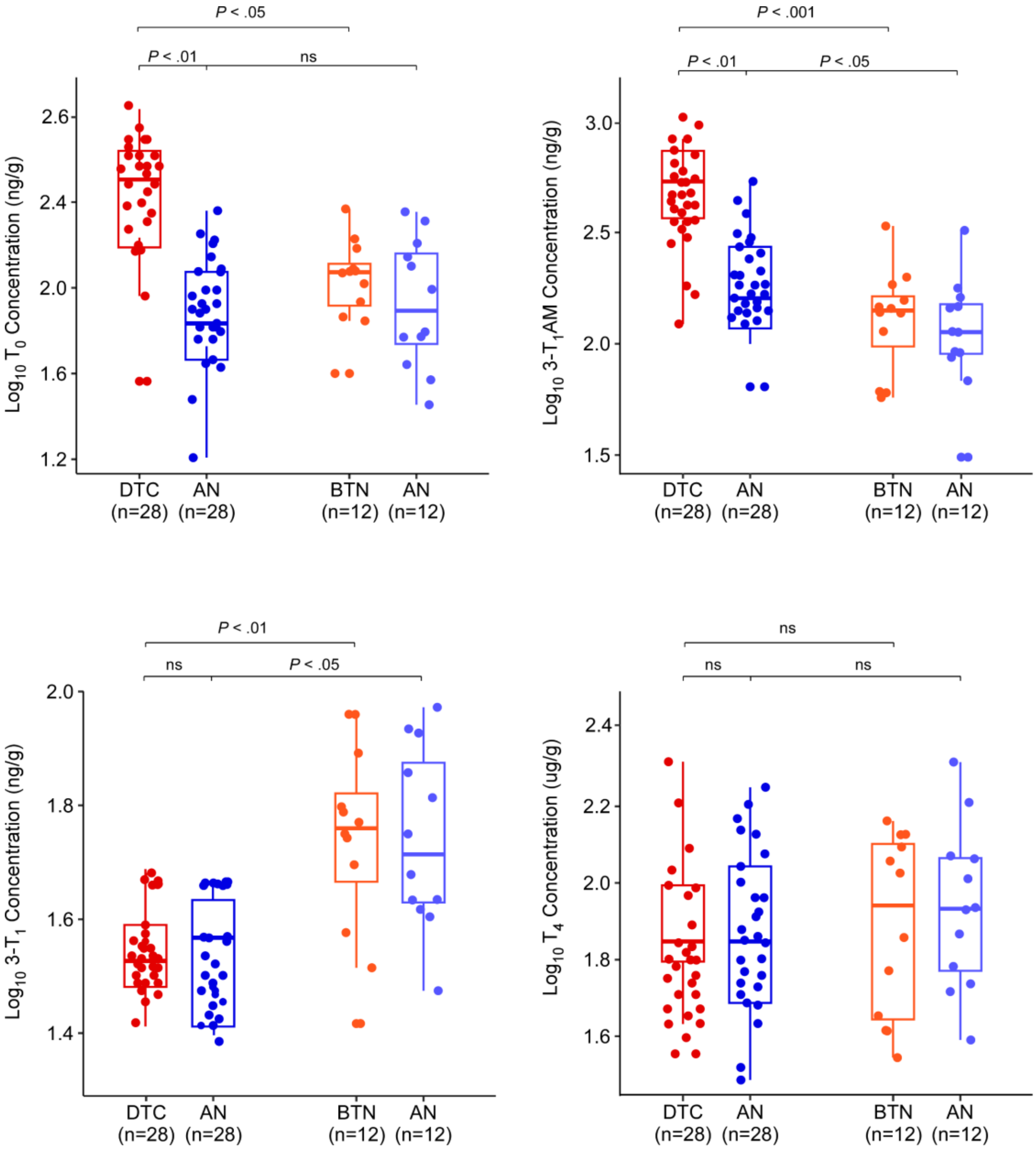

**Figure S5. Test of $T_0$, 3-$T_1$AM, 3-$T_1$, and $T_4$ in tumors of DTC, BTN, and adjacent normal tissues.** $T_0$, 3-$T_1$AM, 3-$T_1$, and $T_4$ were analyzed in twelve DTC, BTN, and adjacent normal tissues. The results showed the statistical differences in $T_0$, 3-$T_1$AM, and 3-$T_1$ between the DTC and BTN groups; the trend was as same as in serum. The results showed no difference in $T_4$ among the four groups. AN, Adjacent normal tissues; BTN, benign thyroid nodules; DTC, differentiated thyroid cancer.

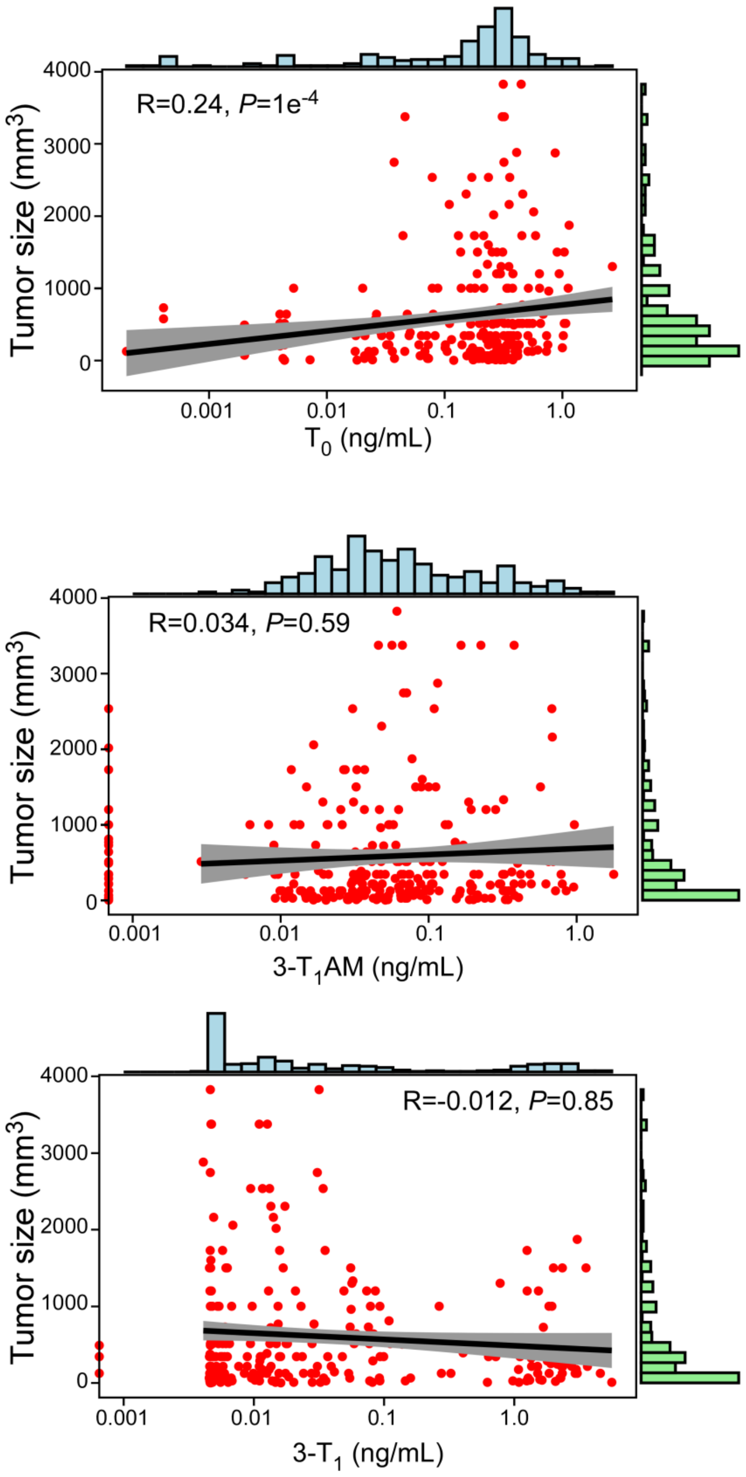

**Figure S6. Correlation between serum $T_0$, 3-$T_1$AM or 3-$T_1$ (ng/ml) and the tumor size of differentiated thyroid cancer (DTC) ($mm^3$).** The tumor size is calculated by length×width×width.

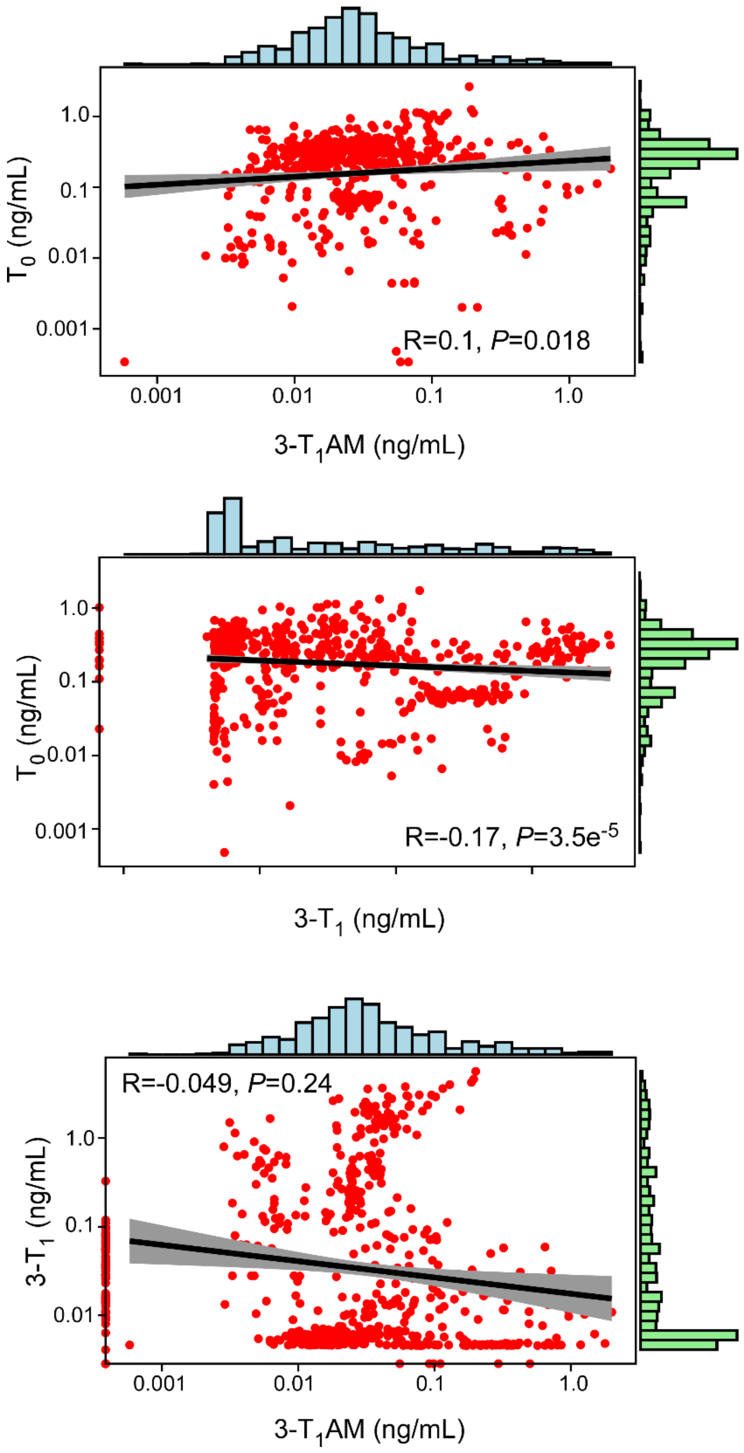

**Figure S7. The relationship among serum $T_0$, 3-$T_1$AM, and 3-$T_1$ in patients with differentiated thyroid cancer (DTC).**

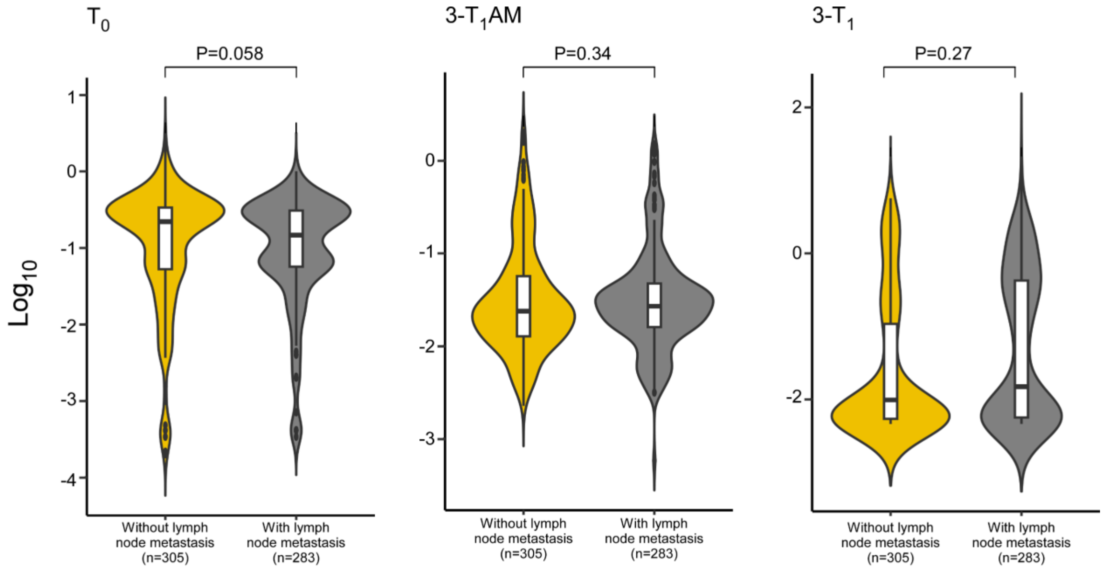

**Figure S8. Comparison of the serum levels of $T_0$, 3-$T_1$AM, and 3-$T_1$ between DTC subjects with and without lymph node metastasis.** DTC, differentiated thyroid cancer.

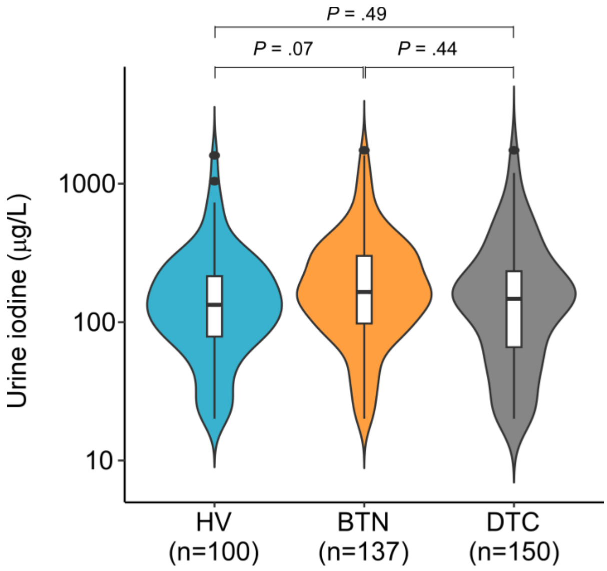

**Figure S9. Urine iodine test.** Median urine iodine concentrations (UIC) were analyzed in the prospectively recruited subjects with available urine specimens, including 100 HV, 137 BTN, and 150 DTC. The median UIC of subjects with DTC, BTN, and HV were 188.24±21.37, 241.68±20.01, and 217.05±29.87, respectively. As shown by the indicated *p* values, there was no statistical difference among the three groups. HV, healthy volunteers; BTN, benign thyroid nodules; DTC, differentiated thyroid cancer.

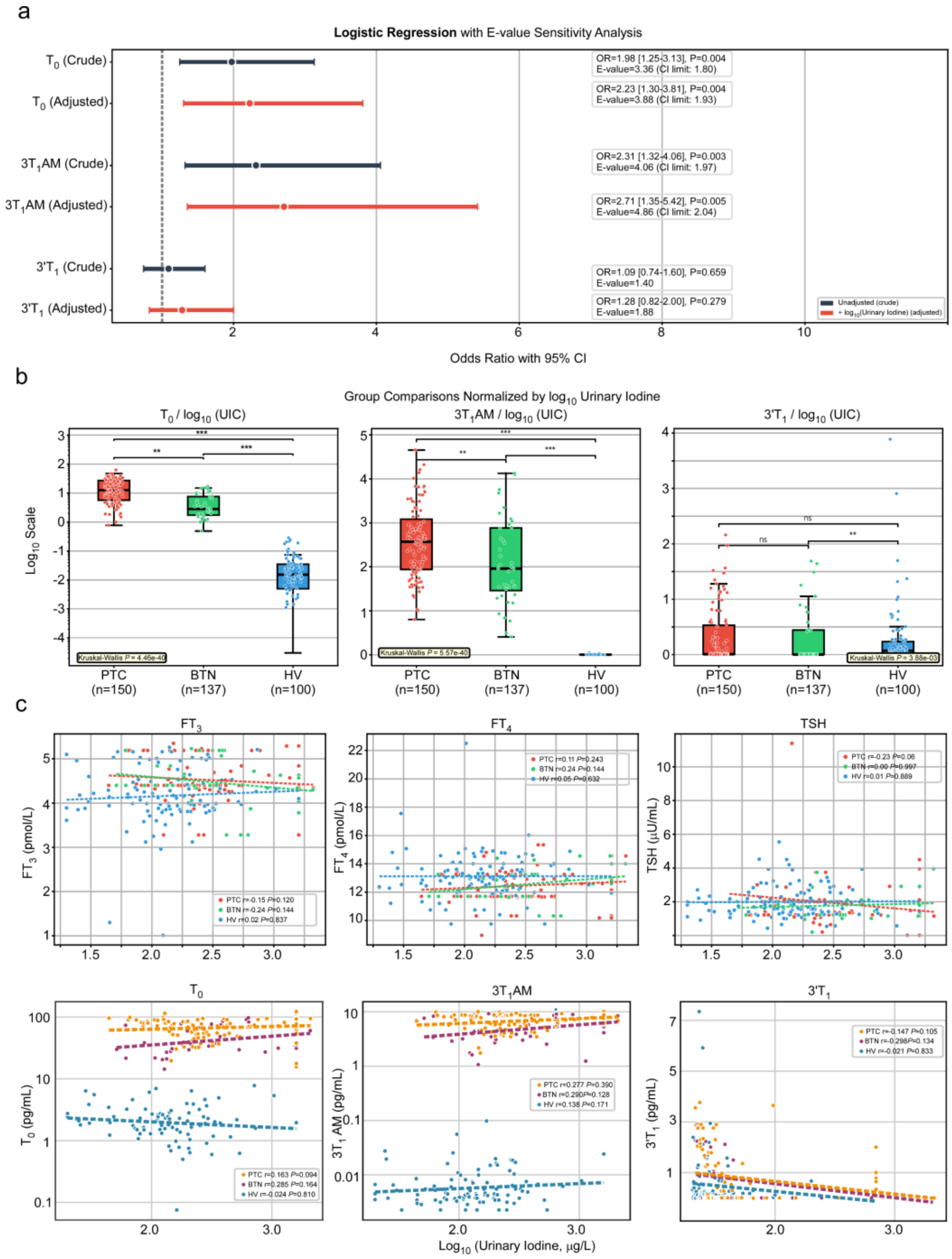

**Figure S10 Analysis of the influence of UIC on THMs**

(A) Crude and $\log_{10}$(urinary iodine)-adjusted ORs comparison for each biomarker using E-value sensitivity analysis; (B) THMs comparison between PTC, BTN and HV by UIC-adjusted; (C) The correlation analyses between UIC and $FT_4$, $FT_3$, TSH as well as THMs.

HV, healthy volunteers; BTN, benign thyroid nodules; PTC, papillary thyroid cancer; THMs, thyroid hormone metabolites

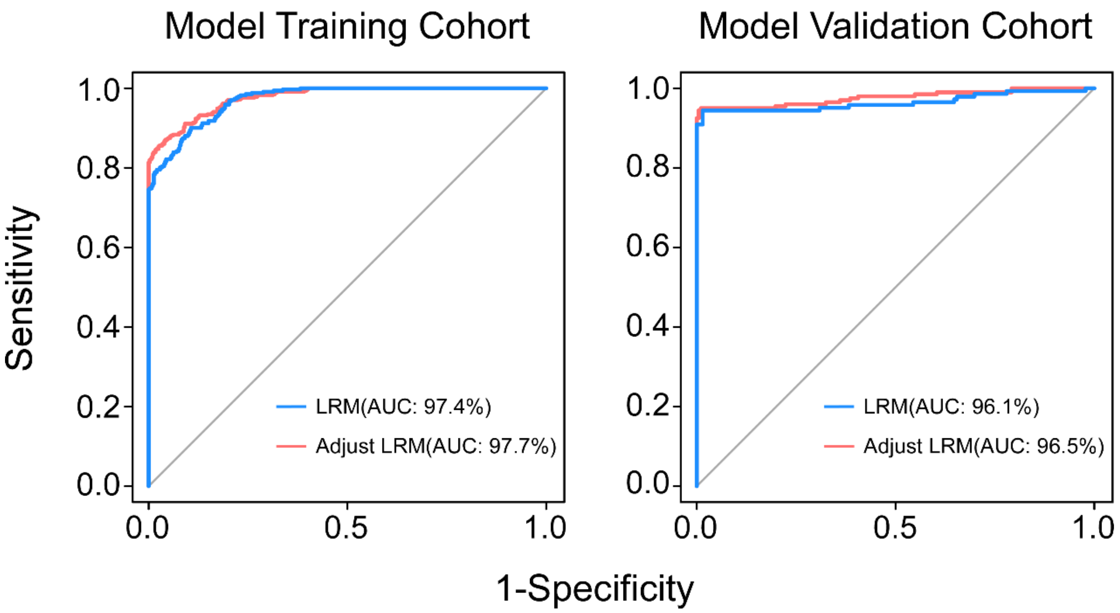

**Figure S11. Logistic Regression Model Calibration.** The calibration of logistic regression model by free thyroxine (FT4).

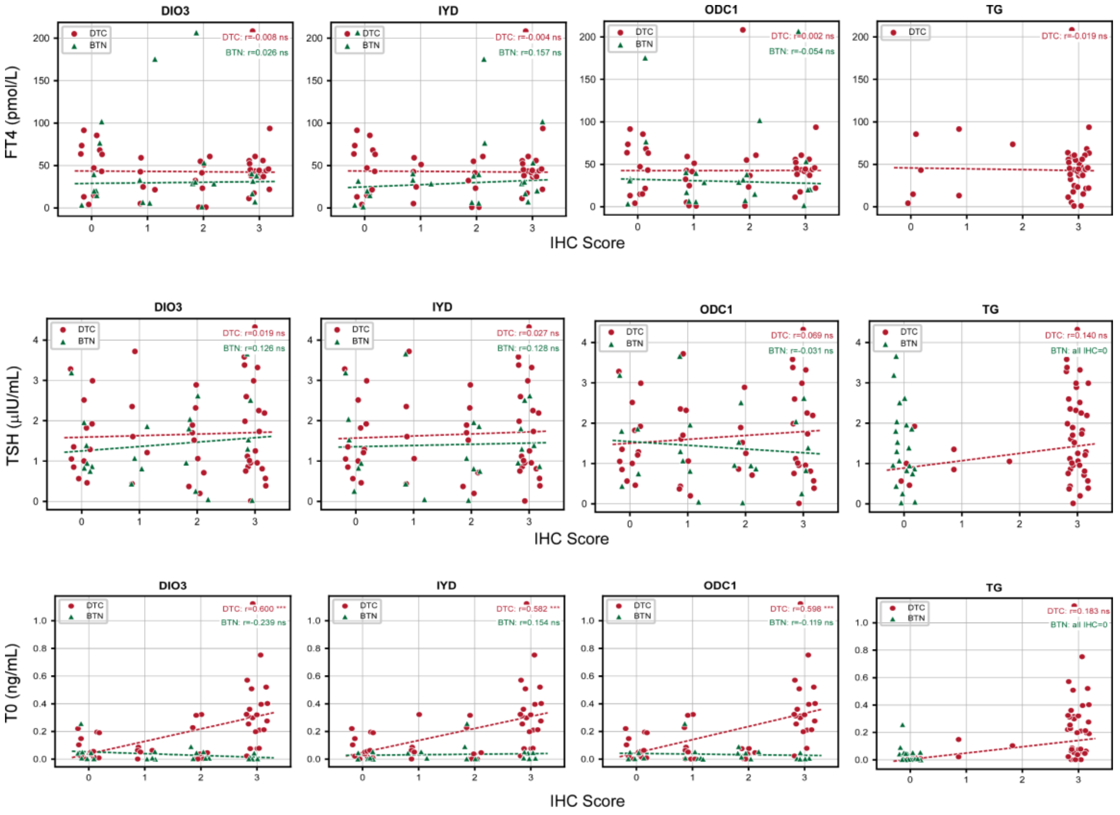

**Figure S12. The correlation expression of metabolizing enzymes in DTC with FT$_4$, TSH, and T$_0$.**

**Supplementary Tables**

**Supplementary Table S1.** Specific number of subjects recruited from each medical center

| Model | Tumor | Total | Xuzhou | Shanghai | Anhui | Liyang | Jinan |
|---|---|---|---|---|---|---|---|
| Training | DTC | 454 | 121 | 139 | 95 | 73 | 26 |
| | BTN | 418 | 219 | 16 | 32 | 21 | 130 |
| Validation | DTC | 195 | 37 | 53 | 51 | 35 | 19 |
| | BTN | 197 | 126 | - | 31 | - | 40 |
| Second validation | DTC | 300 | 300 | | | | |
| | BTN | 99 | 99 | | | | |

Xuzhou=Xuzhou Central Hospital
Shanghai=Fudan university Shanghai cancer center
Anhui=The first affiliated hospital of Anhui medical university
Liyang=Liyang People's Hospital
Jinan=Jinan Central Hospital

**Supplementary Table S2.** Independent histopathological review results of the model training cohort (FFPE specimens) by three senior thyroid pathologists

| **Case ID** | **Histology** | | | **Final Pathology Diagnosis** |
|---|---|---|---|---|
| | **Dr. Wu** | **Dr. Zhu** | **Dr. Zhang** | |
| AH0278 | PTC | PTC | PTC | PTC |
| AH0201 | PTC | PTC | PTC | PTC |
| SH0066 | PTC | TFND | PTC | PTC |
| AH0290 | PTC | PTC | PTC | PTC |
| AH0106 | FTA | PTC | PTC | PTC |
| AH0324 | PTC | PTC | FTC | PTC |
| AH0137 | PTC | PTC | PTC | PTC |
| AH0219 | PTC | PTC | PTC | PTC |
| AH0276 | PTC | PTC | PTC | PTC |
| AH0223 | PTC | PTC | PTC | PTC |
| AH0257 | PTC | PTC | PTC | PTC |
| AH0109 | PTC | FTC | PTC | PTC |
| AH0336 | PTC | PTC | PTC | PTC |
| AH0206 | PTC | PTC | FTA | PTC |
| AH0326 | PTC | PTC | PTC | PTC |

| | | | | |
|---|---|---|---|---|
| AH0100 | PTC | PTC | PTC | PTC |
| AH0331 | PTC | PTC | PTC | PTC |
| AH0320 | PTC | PTC | PTC | PTC |
| AH0261 | PTC | PTC | PTC | PTC |
| AH0310 | PTC | FTC | PTC | PTC |
| AH0262 | PTC | PTC | PTC | PTC |
| AH0202 | PTC | PTC | PTC | PTC |
| AH0285 | PTC | PTC | PTC | PTC |
| AH0252 | TFND | PTC | PTC | PTC |
| AH0022 | PTC | PTC | PTC | PTC |
| AH0319 | PTC | PTC | PTC | PTC |
| AH0213 | PTC | PTC | PTC | PTC |
| AH0272 | PTC | PTC | PTC | PTC |
| AH0322 | PTC | PTC | FTC | PTC |
| AH0259 | PTC | PTC | PTC | PTC |
| AH0195 | PTC | TFND | PTC | PTC |
| AH0155 | PTC | PTC | PTC | PTC |
| AH0260 | PTC | FTA | PTC | PTC |
| AH0108 | PTC | PTC | PTC | PTC |
| AH0173 | PTC | PTC | PTC | PTC |
| AH0315 | PTC | PTC | PTC | PTC |
| AH0171 | PTC | PTC | PTC | PTC |
| AH0283 | PTC | FTA | PTC | PTC |
| AH0308 | PTC | PTC | PTC | PTC |
| AH0332 | PTC | PTC | PTC | PTC |
| AH0178 | PTC | PTC | PTC | PTC |
| SH0093 | PTC | PTC | PTC | PTC |
| AH0174 | PTC | PTC | PTC | PTC |
| AH0134 | TFND | PTC | PTC | PTC |
| AH0318 | PTC | PTC | PTC | PTC |
| AH0175 | PTC | PTC | PTC | PTC |
| AH0263 | PTC | PTC | PTC | PTC |
| AH0279 | PTC | PTC | PTC | PTC |
| SH0084 | PTC | PTC | FTC | PTC |
| AH0334 | PTC | PTC | PTC | PTC |
| AH0179 | PTC | PTC | PTC | PTC |
| SH0090 | PTC | PTC | PTC | PTC |
| AH0316 | PTC | PTC | PTC | PTC |
| SH0079 | PTC | PTC | PTC | PTC |
| AH0341 | PTC | FTC | PTC | PTC |
| AH0196 | PTC | PTC | PTC | PTC |
| AH0101 | PTC | PTC | PTC | PTC |
| SH0081 | PTC | PTC | PTC | PTC |
| SH0041 | PTC | PTC | PTC | PTC |
| AH0364 | PTC | PTC | PTC | PTC |
| AH0170 | TFND | PTC | PTC | PTC |

| | | | | |
|---|---|---|---|---|
| AH0211 | PTC | PTC | PTC | PTC |
| AH0107 | PTC | PTC | PTC | PTC |
| AH0003 | PTC | PTC | PTC | PTC |
| AH0185 | PTC | PTC | PTC | PTC |
| AH0312 | PTC | PTC | PTC | PTC |
| AH0265 | PTC | PTC | PTC | PTC |
| AH0267 | PTC | PTC | PTC | PTC |
| AH0131 | PTC | FTC | PTC | PTC |
| AH0325 | PTC | PTC | PTC | PTC |
| AH0323 | PTC | PTC | PTC | PTC |
| AH0177 | FTA | PTC | PTC | PTC |
| AH0327 | PTC | PTC | PTC | PTC |
| AH0287 | PTC | PTC | PTC | PTC |
| SH0085 | PTC | PTC | PTC | PTC |
| AH0266 | PTC | PTC | PTC | PTC |
| AH0306 | PTC | PTC | PTC | PTC |
| AH0360 | PTC | PTC | PTC | PTC |
| AH0025 | PTC | PTC | PTC | PTC |
| AH0329 | PTC | PTC | PTC | PTC |
| AH0024 | PTC | PTC | PTC | PTC |
| SH0045 | PTC | FTC | PTC | PTC |
| AH0339 | PTC | PTC | PTC | PTC |
| SH0031 | PTC | PTC | PTC | PTC |
| AH0208 | PTC | PTC | PTC | PTC |
| SH0044 | PTC | PTC | PTC | PTC |
| SH0097 | PTC | PTC | FTC | PTC |
| AH0268 | PTC | PTC | PTC | PTC |
| AH0215 | PTC | PTC | PTC | PTC |
| AH0239 | PTC | PTC | PTC | PTC |
| AH0162 | PTC | PTC | PTC | PTC |
| AH0164 | PTC | PTC | PTC | PTC |
| SH0062 | PTC | PTC | PTC | PTC |
| AH0229 | PTC | TFND | PTC | PTC |
| SH0089 | PTC | PTC | PTC | PTC |
| AH0037 | PTC | PTC | PTC | PTC |
| AH0247 | FTC | PTC | PTC | PTC |
| AH0105 | PTC | PTC | PTC | PTC |
| AH0158 | PTC | PTC | PTC | PTC |
| SH0059 | PTC | PTC | PTC | PTC |
| AH0029 | PTC | FTA | PTC | PTC |
| SH0091 | PTC | PTC | PTC | PTC |
| SH0082 | PTC | PTC | PTC | PTC |
| AH0193 | PTC | PTC | PTC | PTC |
| AH0231 | PTC | FTC | PTC | PTC |
| AH0036 | PTC | PTC | PTC | PTC |
| AH0112 | PTC | PTC | PTC | PTC |

| | | | | |
|---|---|---|---|---|
| AH0251 | PTC | PTC | PTC | PTC |
| AH0161 | PTC | PTC | PTC | PTC |
| AH0129 | PTC | PTC | PTC | PTC |
| SH0202 | PTC | PTC | PTC | PTC |
| SH0168 | PTC | TFND | PTC | PTC |
| SH0199 | PTC | PTC | PTC | PTC |
| SH0175 | PTC | PTC | PTC | PTC |
| SH0052 | PTC | PTC | PTC | PTC |
| SH0172 | PTC | PTC | PTC | PTC |
| SH0058 | TFND | PTC | PTC | PTC |
| SH0171 | PTC | PTC | PTC | PTC |
| AH0249 | PTC | PTC | TFND | PTC |
| SH0049 | PTC | PTC | PTC | PTC |
| AH0235 | PTC | PTC | PTC | PTC |
| SH0188 | PTC | PTC | PTC | PTC |
| AH0133 | PTC | PTC | PTC | PTC |
| AH0113 | PTC | PTC | PTC | PTC |
| AH0130 | PTC | PTC | PTC | PTC |
| AH0242 | PTC | PTC | FTC | PTC |
| SH0195 | PTC | PTC | PTC | PTC |
| SH0121 | PTC | PTC | PTC | PTC |
| SH0192 | PTC | PTC | PTC | PTC |
| SH0193 | PTC | PTC | PTC | PTC |
| AH0210 | PTC | PTC | PTC | PTC |
| AH0274 | PTC | TFND | PTC | PTC |
| SH0077 | PTC | PTC | PTC | PTC |
| SH0189 | PTC | PTC | PTC | PTC |
| AH0027 | TFND | PTC | PTC | PTC |
| SH0197 | PTC | PTC | FTC | PTC |
| JS0885 | PTC | PTC | PTC | PTC |
| SH0100 | PTC | PTC | PTC | PTC |
| AH0157 | PTC | FTC | PTC | PTC |
| SH0086 | PTC | PTC | PTC | PTC |
| AH0117 | PTC | PTC | PTC | PTC |
| SH0169 | PTC | PTC | PTC | PTC |
| SH0095 | PTC | PTC | FTA | PTC |
| AH0335 | PTC | PTC | PTC | PTC |
| AH0035 | PTC | PTC | PTC | PTC |
| AH0132 | PTC | PTC | PTC | PTC |
| SH0187 | PTC | PTC | PTC | PTC |
| AH0232 | FTC | PTC | PTC | PTC |
| AH0023 | PTC | PTC | PTC | PTC |
| AH0356 | PTC | PTC | PTC | PTC |
| AH0198 | PTC | PTC | PTC | PTC |
| AH0340 | PTC | PTC | PTC | PTC |
| SH0027 | PTC | PTC | PTC | PTC |

| | | | | |
|---|---|---|---|---|
| AH0209 | PTC | PTC | PTC | PTC |
| SH0057 | PTC | PTC | PTC | PTC |
| AH0338 | PTC | PTC | PTC | PTC |
| AH0004 | PTC | PTC | TFND | PTC |
| AH0228 | PTC | PTC | PTC | PTC |
| JS0877 | PTC | PTC | PTC | PTC |
| SH0024 | PTC | PTC | PTC | PTC |
| SH0092 | PTC | PTC | PTC | PTC |
| JS0874 | PTC | PTC | TFND | PTC |
| AH0030 | PTC | PTC | PTC | PTC |
| SH0096 | PTC | FTC | PTC | PTC |
| SH0060 | PTC | PTC | PTC | PTC |
| SH0038 | FTC | PTC | PTC | PTC |
| JS0862 | PTC+OCA | OCC | PTC+OCA | PTC+OCA |
| JS0859 | PTC | PTC | PTC | PTC |
| JS0882 | PTC+OCA | PTC | PTC+OCA | PTC+OCA |
| AH0240 | PTC | PTC | PTC | PTC |
| JS0858 | PTC | PTC | PTC | PTC |
| JS0863 | PTC | PTC | PTC | PTC |
| JS0865 | PTC | PTC | PTC | PTC |
| JS0860 | PTC | PTC | PTC | PTC |
| AH0026 | PTC | FTC | PTC | PTC |
| SH0117 | PTC | PTC | TFND | PTC |
| AH0214 | PTC | PTC | PTC | PTC |
| AH0226 | PTC | PTC | PTC | PTC |
| JS0857 | FTA | PTC | PTC | PTC |
| SH0035 | PTC | PTC | PTC | PTC |
| SH0144 | PTC | PTC | PTC | PTC |
| AH0358 | PTC | PTC | PTC | PTC |
| AH0220 | PTC | PTC | PTC | PTC |
| JS0876 | PTC | PTC | PTC | PTC |
| SH0178 | PTC | PTC | PTC | PTC |
| JS0861 | PTC | PTC | PTC | PTC |
| JS0851 | OCC | OCC | PTC | OCC |
| JS0852 | FTC | FTC | PTC | FTC |
| SH0185 | PTC | PTC | PTC | PTC |
| JS0888 | PTC | PTC | PTC | PTC |
| SH0204 | PTC | PTC | PTC | PTC |
| JS0890 | PTC | PTC | PTC | PTC |
| SH0176 | PTC | PTC | PTC | PTC |
| JS0872 | PTC | PTC | PTC | PTC |
| SH0194 | PTC | PTC | PTC | PTC |
| JS0871 | PTC | PTC | PTC | PTC |
| JS0847 | PTC | PTC | PTC | PTC |
| SH0040 | PTC | PTC | PTC | PTC |
| JS0846 | OCC | PTC | OCC | OCC |

| | | | | |
|---|---|---|---|---|
| JS0887 | PTC | PTC | PTC | PTC |
| JS0870 | PTC | PTC | PTC | PTC |
| JS0868 | PTC | PTC | PTC | PTC |
| JS0867 | PTC | PTC | PTC | PTC |
| JS0869 | PTC | PTC | PTC | PTC |
| SH0029 | PTC | PTC | PTC | PTC |
| SH0070 | PTC | PTC | PTC | PTC |
| SH0020 | PTC | TFND | PTC | PTC |
| AH0127 | PTC | PTC | FTC | PTC |
| SH0145 | PTC | PTC | PTC | PTC |
| AH0245 | PTC | PTC | PTC | PTC |
| AH0138 | TFND | PTC | PTC | PTC |
| SH0164 | PTC | PTC | PTC | PTC |
| SH0098 | PTC | PTC | PTC | PTC |
| AH0303 | PTC | PTC | PTC | PTC |
| AH0207 | PTC | PTC | PTC | PTC |
| AH0227 | PTC | PTC | FTA | PTC |
| SH0063 | PTC | PTC | PTC | PTC |
| SH0074 | PTC | PTC | PTC | PTC |
| SH0138 | PTC | PTC | PTC | PTC |
| SH0075 | PTC | PTC | PTC | PTC |
| SH0120 | PTC | FTC | PTC | PTC |
| SH0042 | PTC | PTC | PTC | PTC |
| SH0072 | PTC | PTC | PTC | PTC |
| SH0107 | PTC | PTC | PTC | PTC |
| JS0873 | PTC | PTC | PTC | PTC |
| AH0230 | FTC | PTC | PTC | PTC |
| AH0028 | PTC | PTC | PTC | PTC |
| SH0083 | PTC | PTC | PTC | PTC |
| SH0130 | PTC | PTC | PTC | PTC |
| SH0073 | PTC | PTC | PTC | PTC |
| SH0123 | PTC | PTC | PTC | PTC |
| SH0101 | PTC | PTC | PTC | PTC |
| JS0800 | PTC | PTC | PTC | PTC |
| SH0039 | PTC | PTC | PTC | PTC |
| SH0150 | PTC | PTC | PTC | PTC |
| SH0078 | PTC | PTC | PTC | PTC |
| AH0186 | PTC | PTC | TFND | PTC |
| SH0152 | PTC | PTC | PTC | PTC |
| SH0143 | PTC | PTC | PTC | PTC |
| AH0313 | PTC | PTC | PTC | PTC |
| SH0076 | PTC | PTC | PTC | PTC |
| AH0110 | PTC | PTC | FTA | PTC |
| AH0121 | PTC | PTC | PTC | PTC |
| SH0003 | PTC | PTC | PTC | PTC |
| AH0225 | PTC | PTC | PTC | PTC |

| | | | | |
|---|---|---|---|---|
| SH0112 | PTC | PTC | PTC | PTC |
| SH0113 | PTC | PTC | PTC | PTC |
| AH0104 | PTC | PTC | PTC | PTC |
| SH0025 | PTC | PTC | PTC | PTC |
| SH0153 | PTC | PTC | PTC | PTC |
| SH0016 | PTC | PTC | PTC | PTC |
| SH0051 | PTC | PTC | PTC | PTC |
| SH0094 | PTC | PTC | PTC | PTC |
| AH0140 | PTC | PTC | PTC | PTC |
| SH0142 | PTC | FTC | PTC | PTC |
| AH0244 | PTC | PTC | PTC | PTC |
| SH0103 | PTC | PTC | PTC | PTC |
| SH0122 | PTC | PTC | PTC | PTC |
| SH0102 | PTC | PTC | PTC | PTC |
| AH0218 | PTC | PTC | PTC | PTC |
| SH0013 | PTC | PTC | PTC | PTC |
| SH0056 | PTC | PTC | PTC | PTC |
| SH0116 | PTC | PTC | PTC | PTC |
| JS0699 | FTC | FTA | FTA | FTA |
| AH0281 | PTC | PTC | PTC | PTC |
| SH0139 | PTC | PTC | PTC | PTC |
| SH0050 | PTC | PTC | PTC | PTC |
| SH0165 | PTC | PTC | PTC | PTC |
| SH0055 | PTC | PTC | PTC | PTC |
| SH0119 | PTC | PTC | PTC | PTC |
| SH0065 | PTC | PTC | PTC | PTC |
| AH0187 | PTC | PTC | PTC | PTC |
| SH0001 | PTC | PTC | PTC | PTC |
| SH0018 | PTC | PTC | PTC | PTC |
| AH0114 | PTC | PTC | PTC | PTC |
| SH0088 | PTC | PTC | PTC | PTC |
| SH0155 | PTC | PTC | PTC | PTC |
| SH0140 | PTC | PTC | PTC | PTC |
| SH0109 | PTC | PTC | PTC | PTC |
| AH0248 | PTC | PTC | PTC | PTC |
| SH0128 | PTC | PTC | PTC | PTC |
| JS0696 | FTA | FTA | PTC | FTA |
| AH0122 | PTC | PTC | PTC | PTC |
| AH0243 | FTA | PTC | PTC | PTC |
| AH0191 | PTC | PTC | PTC | PTC |
| SH0008 | PTC | PTC | PTC | PTC |
| JS0793 | OCC | PTC | OCC | OCC |
| SH0047 | PTC | PTC | PTC | PTC |
| SH0156 | PTC | PTC | PTC | PTC |
| JS0799 | PTC | PTC | PTC | PTC |
| SH0137 | PTC | PTC | PTC | PTC |

| | | | | |
|---|---|---|---|---|
| SH0032 | PTC | PTC | TFND | PTC |
| SH0162 | PTC | PTC | PTC | PTC |
| SH0054 | PTC | FTC | PTC | PTC |
| SH0068 | PTC | PTC | PTC | PTC |
| SH0111 | FTC | PTC | PTC | PTC |
| SH0005 | PTC | PTC | PTC | PTC |
| SH0048 | PTC | PTC | PTC | PTC |
| SH0125 | PTC | PTC | PTC | PTC |
| SH0006 | PTC | PTC | PTC | PTC |
| JS0750 | PTC | PTC | PTC | PTC |
| AH0199 | PTC | PTC | PTC | PTC |
| SH0154 | PTC | PTC | PTC | PTC |
| AH0236 | PTC | PTC | PTC | PTC |
| SH0115 | PTC | PTC | PTC | PTC |
| SH0110 | PTC | FTA | PTC | PTC |
| SH0022 | PTC | PTC | PTC | PTC |
| SH0009 | FTC | PTC | PTC | PTC |
| JS0770 | TFND | TFND | PTC | TFND |
| SH0021 | PTC | PTC | PTC | PTC |
| JS0801 | PTC | PTC | PTC | PTC |
| JS0749 | PTC | PTC | PTC | PTC |
| AH0126 | FTC | PTC | PTC | PTC |
| JS0789 | PTC | PTC | PTC | PTC |
| AH0116 | PTC | PTC | PTC | PTC |
| AH0205 | PTC | PTC | PTC | PTC |
| JS0796 | PTC | PTC | PTC | PTC |
| JS0812 | PTC | PTC | PTC | PTC |
| JS0803 | PTC | PTC | PTC | PTC |
| JS0798 | FTA | PTC | PTC | PTC |
| JS0828 | FTA | PTC | FTA | FTA |
| JS0788 | PTC | PTC | PTC | PTC |
| SH0106 | PTC | PTC | PTC | PTC |
| JS0813 | PTC | PTC | PTC | PTC |
| JS0760 | PTC | FTA | FTA | FTA |
| JS0787 | PTC | PTC | PTC | PTC |
| JS0802 | PTC | PTC | PTC | PTC |
| JS0706 | FTA | FTA | PTC | FTA |
| AH0033 | PTC | PTC | FTC | PTC |
| JS0811 | PTC | PTC | PTC | PTC |
| JS0767 | FTA | PTC | FTA | FTA |
| JS0804 | PTC | PTC | PTC | PTC |
| JS0810 | PTC | PTC | PTC | PTC |
| JS0809 | PTC | PTC | PTC | PTC |
| JS0758 | PTC | FTA | FTA | FTA |
| SH0067 | FTC | FTC | TFND | FTC |
| AH0317 | FTC | FTC | FTC | FTC |

| | | | | |
|---|---|---|---|---|
| JS0794 | HT | FTC | FTC | FTC |
| AH0047 | TFND | FTA | TFND | TFND |
| AH0153 | TFND | FTA | FTA | FTA |
| AH0292 | TFND | TFND | TFND | TFND |
| AH0048 | TFND | TFND | TFND | TFND |
| AH0295 | FTA | FTA | TFND | FTA |
| AH0046 | FTA | TFND | TFND | TFND |
| AH0079 | TFND | TFND | FTA | TFND |
| AH0141 | TFND | FTA | TFND | TFND |
| AH0350 | FTA | FTA | FTA | FTA |
| AH0353 | HT | HT | TFND | HT |
| AH0144 | FTA | TFND | TFND | TFND |
| AH0351 | FTA | FTA | HT | FTA |
| AH0093 | FTA | TFND | TFND | TFND |
| AH0078 | TFND | TFND | TFND | TFND |
| AH0352 | FTA | FTA | FTA | FTA |
| AH0075 | TFND | TFND | TFND | TFND |
| AH0015 | TFND | FTA | TFND | TFND |
| AH0052 | TFND | HT | HT | HT |
| AH0084 | TFND | TFND | TFND | TFND |
| AH0142 | TFND | TFND | HT | TFND |
| SH0208 | TFND | FTA | TFND | TFND |
| SH0207 | TFND | FTA | TFND | TFND |
| AH0151 | HT | TFND | TFND | TFND |
| AH0092 | TFND | FTA | TFND | TFND |
| AH0008 | TFND | FTA | FTA | FTA |
| AH0343 | TFND | TFND | TFND | TFND |
| AH0333 | FTA | FTA | FTA | FTA |
| AH0010 | TFND | TFND | TFND | TFND |
| AH0149 | OCA | OCA | FTA | OCA |
| SH0213 | TFND | HT | TFND | TFND |
| SH0206 | HT | TFND | TFND | TFND |
| AH0145 | TFND | TFND | FTA | TFND |
| SH0215 | TFND | TFND | TFND | TFND |
| SH0214 | TFND | FTA | TFND | TFND |
| AH0055 | TFND | TFND | TFND | TFND |
| AH0056 | TFND | HT | TFND | TFND |
| AH0054 | TFND | TFND | TFND | TFND |
| AH0296 | TFND | FTA | FTA | FTA |
| AH0049 | TFND | FTA | TFND | TFND |
| AH0041 | TFND | FTA | TFND | TFND |
| AH0347 | HT | TFND | TFND | TFND |
| AH0057 | FTA | TFND | TFND | TFND |
| AH0058 | TFND | TFND | HT | TFND |
| AH0152 | TFND | FTA | TFND | TFND |
| AH0059 | TFND | TFND | TFND | TFND |

| | | | | |
|---|---|---|---|---|
| AH0297 | HT | FTA | FTA | FTA |
| AH0094 | FTA | TFND | TFND | TFND |
| AH0051 | TFND | FTA | TFND | TFND |
| JS2001 | FTC | FTC | PTC | FTC |
| JS2004 | PTC | FTA | PTC | PTC |
| JS2005 | PTC | HT | PTC | PTC |
| JS2008 | PTC | PTC | PTC | PTC |
| JS2010 | PTC | FTC | PTC | PTC |
| JS2013 | PTC | FTC | PTC | PTC |
| JS2014 | PTC | PTC | FTA | PTC |
| JS2016 | PTC | PTC | HT | PTC |
| JS2018 | PTC | PTC | PTC | PTC |
| JS2019 | PTC | PTC | PTC | PTC |
| JS2022 | PTC | FTC | PTC | PTC |
| JS2023 | PTC | FTC | PTC | PTC |
| JS2024 | FTC | FTC | PTC | FTC |
| JS2025 | PTC | PTC | HT | PTC |
| JS2026 | PTC | PTC | PTC | PTC |
| JS2027 | FTC | PTC | PTC | PTC |
| JS2028 | PTC | FTA | PTC | PTC |
| JS2029 | PTC | FTC | PTC | PTC |
| JS2030 | PTC | PTC | PTC | PTC |
| JS2031 | PTC | FTC | PTC | PTC |
| JS2032 | PTC | PTC | PTC | PTC |
| JS2033 | FTC | PTC | PTC | PTC |
| JS2036 | FTA | PTC | PTC | PTC |
| JS2037 | PTC | PTC | FTA | PTC |
| JS2039 | PTC | PTC | FTA | PTC |
| JS2040 | PTC | FTC | PTC | PTC |
| JS2041 | PTC | PTC | FTC | PTC |
| JS2042 | PTC | PTC | PTC | PTC |
| JS2043 | PTC | FTA | PTC | PTC |
| JS2044 | PTC | PTC | PTC | PTC |
| JS2046 | PTC | HT | PTC | PTC |
| JS2048 | PTC | PTC | FTC | PTC |
| JS2049 | FTC | FTC | FTC | FTC |
| JS2051 | PTC | PTC | PTC | PTC |
| JS2052 | PTC | PTC | HT | PTC |
| JS2053 | PTC | FTA | PTC | PTC |
| JS2054 | PTC | PTC | PTC | PTC |
| JS2056 | PTC | PTC | PTC | PTC |
| JS2057 | PTC | PTC | PTC | PTC |
| JS2058 | PTC | FTC | PTC | PTC |
| JS2061 | PTC | PTC | PTC | PTC |
| JS2062 | PTC | FTC | PTC | PTC |
| JS2063 | PTC | PTC | PTC | PTC |

| | | | | |
|---|---|---|---|---|
| JS2065 | PTC | PTC | PTC | PTC |
| JS2067 | PTC | PTC | PTC | PTC |
| JS2069 | PTC | PTC | PTC | PTC |
| JS2070 | PTC | FTC | PTC | PTC |
| JS2071 | FTC | FTC | PTC | FTC |
| JS2074 | PTC | FTC | PTC | PTC |
| JS2076 | PTC | PTC | HT | PTC |
| JS2077 | FTC | PTC | PTC | PTC |
| JS2078 | FTA | PTC | PTC | PTC |
| JS2083 | PTC | PTC | HT | PTC |
| JS2084 | PTC | PTC | PTC | PTC |
| JS2085 | PTC | FTC | PTC | PTC |
| JS2086 | PTC | PTC | PTC | PTC |
| JS2087 | PTC | PTC | PTC | PTC |
| JS2089 | PTC | FTC | PTC | PTC |
| JS2091 | FTC | FTC | FTC | FTC |
| JS2093 | PTC | FTC | FTC | FTC |
| JS2095 | PTC | FTA | PTC | PTC |
| JS2096 | PTC | PTC | PTC | PTC |
| JS2098 | PTC | PTC | PTC | PTC |
| JS2100 | PTC | PTC | PTC | PTC |
| JS2101 | PTC | FTC | PTC | PTC |
| JS2102 | PTC | PTC | PTC | PTC |
| JS2103 | PTC | FTA | PTC | PTC |
| JS2104 | PTC | PTC | PTC | PTC |
| JS2105 | PTC | PTC | PTC | PTC |
| JS2106 | PTC | PTC | HT | PTC |
| JS2107 | PTC | PTC | PTC | PTC |
| JS2108 | PTC | FTC | FTC | FTC |
| JS2110 | PTC | PTC | FTC | PTC |
| JS2112 | PTC | PTC | FTA | PTC |
| JS2114 | PTC | PTC | PTC | PTC |
| JS2115 | PTC | PTC | PTC | PTC |
| JS2117 | PTC | FTC | PTC | PTC |
| JS2119 | FTC | PTC | PTC | PTC |
| JS2121 | PTC | PTC | PTC | PTC |
| JS2123 | PTC | PTC | PTC | PTC |
| JS2125 | PTC | PTC | PTC | PTC |
| JS2127 | PTC | FTC | PTC | PTC |
| JS2128 | PTC | FTC | PTC | PTC |
| JS2129 | FTC | PTC | PTC | PTC |
| JS2130 | PTC | PTC | PTC | PTC |
| JS2132 | PTC | PTC | PTC | PTC |
| JS2133 | FTC | PTC | PTC | PTC |
| JS2134 | PTC | PTC | PTC | PTC |
| JS2135 | FTC | PTC | PTC | PTC |

| | | | | |
|---|---|---|---|---|
| JS2137 | FTC | PTC | PTC | PTC |
| JS2139 | PTC | PTC | FTC | PTC |
| JS2140 | PTC | PTC | PTC | PTC |
| JS2141 | PTC | PTC | PTC | PTC |
| JS2146 | FTC | PTC | FTC | FTC |
| JS2147 | PTC | FTC | PTC | PTC |
| JS2149 | HT | PTC | PTC | PTC |
| JS2151 | PTC | PTC | PTC | PTC |
| JS2154 | PTC | PTC | PTC | PTC |
| JS2158 | PTC | PTC | PTC | PTC |
| JS2161 | PTC | PTC | PTC | PTC |
| JS2162 | FTC | PTC | PTC | PTC |
| JS2164 | PTC | FTC | PTC | PTC |
| JS2167 | OCC | OCC | PTC | OCC |
| JS2171 | PTC | PTC | PTC | PTC |
| JS2175 | FTC | PTC | PTC | PTC |
| JS2180 | PTC | PTC | PTC | PTC |
| JS2181 | PTC | PTC | PTC | PTC |
| JS2182 | PTC | PTC | PTC | PTC |
| JS2184 | OCC | OCC | PTC | OCC |
| JS2185 | PTC | OCC | OCC | OCC |
| JS2187 | PTC | PTC | FTA | PTC |
| JS2188 | PTC | PTC | FTC | PTC |
| JS2191 | PTC | HT | PTC | PTC |
| JS2192 | PTC | PTC | PTC | PTC |
| JS2194 | PTC | PTC | PTC | PTC |
| JS2195 | PTC | PTC | PTC | PTC |
| JS2197 | FTC | PTC | PTC | PTC |
| JS2198 | FTA | PTC | PTC | PTC |
| JS2199 | FTC | PTC | PTC | PTC |
| JS2201 | PTC | PTC | PTC | PTC |
| JS3003 | FTA | HT | FTA | FTA |
| JS3005 | FTA | TFND | FTA | FTA |
| JS3007 | FTA | TFND | TFND | TFND |
| JS3011 | FTA | TFND | TFND | TFND |
| JS3016 | FTA | FTA | HT | FTA |
| JS3027 | OCA | OCA | TFND | OCA |
| JS3029 | TFND | FTA | FTA | FTA |
| JS3032 | FTA | FTA | HT | FTA |
| JS3035 | TFND | TFND | HT | TFND |
| JS3040 | FTA | FTA | TFND | FTA |
| JS3044 | TFND | TFND | FTA | TFND |
| JS3047 | TFND | TFND | FTA | TFND |
| JS3049 | FTA | HT | FTA | FTA |
| JS3052 | FTA | HT | FTA | FTA |
| JS3055 | FTA | FTA | TFND | FTA |

| | | | | |
|---|---|---|---|---|
| JS3062 | TFND | TFND | FTA | TFND |
| JS3065 | TFND | HT | TFND | TFND |
| JS3070 | TFND | TFND | FTA | FTA |
| JS4001 | TFND | FTA | FTA | FTA |
| JS4002 | FTA | HT | FTA | FTA |
| JS4003 | TFND | HT | TFND | TFND |
| JS4004 | FTA | FTA | TFND | FTA |
| JS4005 | TFND | TFND | FTA | TFND |
| JS4006 | FTA | FTA | TFND | FTA |
| JS4007 | TFND | FTA | TFND | TFND |
| JS4008 | FTA | TFND | FTA | FTA |
| JS4009 | FTA | FTA | HT | FTA |
| JS4010 | TFND | TFND | FTA | TFND |
| JS4011 | TFND | FTA | TFND | TFND |
| JS4012 | FTA | TFND | TFND | TFND |
| JS4013 | FTA | TFND | TFND | TFND |
| JS4014 | TFND | FTA | FTA | FTA |
| JS4015 | FTA | HT | FTA | FTA |
| JS4016 | FTA | FTA | TFND | FTA |
| JS4017 | TFND | TFND | FTA | TFND |
| JS4018 | FTA | TFND | FTA | FTA |
| JS4019 | HT | TFND | TFND | TFND |
| JS4020 | FTA | HT | FTA | FTA |
| JS4021 | TFND | FTA | TFND | TFND |
| JS4022 | FTA | TFND | FTA | FTA |
| JS4023 | FTA | FTA | TFND | FTA |
| JS4024 | TFND | FTA | FTA | FTA |
| JS4025 | TFND | FTA | FTA | FTA |
| JS4026 | TFND | FTA | FTA | FTA |
| JS4027 | TFND | FTA | TFND | TFND |
| JS4028 | FTA | HT | FTA | FTA |
| JS4029 | TFND | TFND | FTA | TFND |
| JS4030 | FTA | FTA | TFND | FTA |
| JS4031 | FTA | FTA | TFND | FTA |
| JS4032 | FTA | TFND | FTA | FTA |
| JS4033 | FTA | TFND | TFND | TFND |
| JS4034 | HT | FTA | FTA | FTA |
| JS4035 | TFND | FTA | FTA | FTA |
| JS4036 | FTA | TFND | TFND | TFND |

TFND, Thyroid follicular nodular disease.
HT, Hashimoto's thyroiditis.
FTA, follicular thyroid adenoma.
FTC, follicular thyroid carcinoma.
PTC, papillary thyroid carcinoma.
OCC, oncocytic thyroid carcinoma
OCA, oncocytic thyroid adenoma

**Supplementary Table S3.** Independent histopathological review results of the model validation cohort (FFPE specimens) by three senior thyroid pathologists

| Case ID | Histology | | | Final Pathology Diagnosis |
|---|---|---|---|---|
| | Dr. Wu | Dr. Zhu | Dr. Zhang | |
| AH0005 | FTC | PTC | PTC | PTC |
| AH0007 | PTC | PTC | PTC | PTC |
| AH0009 | PTC | PTC | PTC | PTC |
| AH0011 | PTC | PTC | PTC | PTC |
| AH0031 | PTC | PTC | PTC | PTC |
| AH0032 | PTC | PTC | PTC | PTC |
| AH0034 | PTC | PTC | PTC | PTC |
| AH0038 | PTC | PTC | PTC | PTC |
| AH0098 | PTC | PTC | PTC | PTC |
| AH0099 | FTA | PTC | PTC | PTC |
| AH0102 | PTC | PTC | PTC | PTC |
| AH0111 | PTC | PTC | PTC | PTC |
| AH0115 | PTC | PTC | PTC | PTC |
| AH0119 | PTC | PTC | PTC | PTC |
| AH0124 | PTC | PTC | PTC | PTC |
| AH0125 | PTC | PTC | PTC | PTC |
| AH0128 | PTC | PTC | PTC | PTC |
| AH0136 | FTA | PTC | PTC | PTC |
| AH0139 | TFND | PTC | PTC | PTC |
| AH0156 | PTC | PTC | PTC | PTC |
| AH0163 | PTC | PTC | PTC | PTC |
| AH0169 | PTC | PTC | PTC | PTC |
| AH0172 | PTC | PTC | PTC | PTC |
| AH0180 | PTC | FTC | PTC | PTC |
| AH0182 | PTC | PTC | PTC | PTC |
| AH0188 | PTC | PTC | PTC | PTC |
| AH0197 | PTC | PTC | PTC | PTC |
| AH0200 | PTC | PTC | PTC | PTC |
| AH0204 | PTC | PTC | PTC | PTC |
| AH0212 | PTC | PTC | PTC | PTC |
| AH0216 | PTC | PTC | PTC | PTC |
| AH0217 | PTC | PTC | PTC | PTC |
| AH0221 | PTC | PTC | PTC | PTC |
| AH0222 | PTC | PTC | PTC | PTC |
| AH0224 | PTC | TFND | PTC | PTC |
| AH0233 | PTC | PTC | PTC | PTC |
| AH0234 | PTC | PTC | PTC | PTC |

| | | | | |
|---|---|---|---|---|
| AH0237 | PTC | PTC | PTC | PTC |
| AH0238 | PTC | PTC | PTC | PTC |
| AH0241 | PTC | PTC | PTC | PTC |
| AH0250 | PTC | PTC | PTC | PTC |
| AH0253 | PTC | PTC | PTC | PTC |
| AH0256 | FTC | FTC | TFND | FTC |
| AH0258 | PTC | PTC | PTC | PTC |
| AH0264 | FTA | PTC | PTC | PTC |
| AH0270 | PTC | PTC | PTC | PTC |
| AH0275 | PTC | PTC | PTC | PTC |
| AH0282 | PTC | PTC | PTC | PTC |
| AH0284 | PTC | PTC | PTC | PTC |
| AH0288 | PTC | PTC | PTC | PTC |
| AH0289 | PTC | PTC | PTC | PTC |
| AH0291 | PTC | PTC | PTC | PTC |
| AH0300 | PTC | PTC | PTC | PTC |
| AH0301 | PTC | FTA | PTC | PTC |
| AH0304 | PTC | PTC | PTC | PTC |
| AH0307 | PTC | FTC | PTC | PTC |
| AH0314 | PTC | PTC | PTC | PTC |
| AH0328 | PTC | PTC | PTC | PTC |
| AH0337 | FTA | PTC | PTC | PTC |
| AH0342 | PTC | PTC | PTC | PTC |
| AH0354 | PTC | PTC | PTC | PTC |
| AH0355 | PTC | PTC | FTC | PTC |
| AH0359 | PTC | PTC | PTC | PTC |
| AH0362 | PTC | PTC | PTC | PTC |
| AH0363 | PTC | PTC | PTC | PTC |
| AH0367 | FTA | TFND | TFND | TFND |
| AH0368 | FTA | TFND | TFND | TFND |
| AH0377 | TFND | TFND | TFND | TFND |
| AH0381 | PTC | PTC | FTC | PTC |
| AH0384 | FTA | TFND | TFND | TFND |
| AH0391 | FTA | TFND | TFND | TFND |
| AH0402 | FTA | TFND | TFND | TFND |
| AH0409 | TFND | TFND | TFND | TFND |
| AH0412 | FTA | TFND | TFND | TFND |
| AH0416 | TFND | TFND | TFND | TFND |
| AH0418 | TFND | TFND | HT | TFND |
| JS0718 | PTC | PTC | PTC | PTC |
| JS0720 | PTC | FTC | PTC | PTC |
| JS0723 | PTC | PTC | PTC | PTC |
| JS0731 | PTC | PTC | PTC | PTC |
| JS0746 | PTC | PTC | PTC | PTC |
| JS0751 | PTC | PTC | PTC | PTC |
| JS0752 | PTC | PTC | PTC | PTC |

| | | | | |
|---|---|---|---|---|
| JS0763 | PTC | FTC | PTC | PTC |
| JS0769 | PTC | PTC | PTC | PTC |
| JS0771 | PTC | PTC | PTC | PTC |
| JS0786 | PTC | PTC | PTC | PTC |
| JS0790 | PTC | PTC | PTC | PTC |
| JS0791 | FTC | PTC+OCA | PTC+OCA | PTC+OCA |
| JS0792 | PTC | PTC | PTC | PTC |
| JS0795 | FTC | FTC | HT | FTC |
| JS0797 | PTC | PTC | PTC | PTC |
| JS0805 | PTC | PTC | PTC | PTC |
| JS0806 | FTC | FTC | PTC | FTC |
| JS0807 | PTC | PTC | PTC | PTC |
| JS0808 | PTC | PTC | PTC | PTC |
| JS0814 | FTA | PTC | PTC | PTC |
| JS0825 | PTC | PTC | PTC | PTC |
| JS0844 | PTC | PTC | PTC | PTC |
| JS0848 | PTC | PTC | PTC | PTC |
| JS0875 | PTC | PTC | PTC | PTC |
| JS0878 | PTC | PTC | PTC | PTC |
| JS0991 | FTA | TFND | TFND | TFND |
| JS0992 | FTA | TFND | TFND | TFND |
| JS0993 | FTA | TFND | TFND | TFND |
| JS0994 | TFND | TFND | TFND | TFND |
| JS0995 | FTA | TFND | TFND | TFND |
| JS0996 | TFND | TFND | TFND | TFND |
| JS0997 | FTA | TFND | TFND | TFND |
| JS0998 | FTA | TFND | TFND | TFND |
| JS0999 | TFND | HT | TFND | TFND |
| JS1000 | TFND | TFND | TFND | TFND |
| JS1001 | FTA | FTA | TFND | FTA |
| JS1002 | TFND | TFND | TFND | TFND |
| JS1003 | TFND | TFND | TFND | TFND |
| JS1004 | FTA | FTA | FTA | FTA |
| JS1005 | TFND | TFND | TFND | TFND |
| JS1006 | FTA | FTA | FTA | FTA |
| JS1007 | TFND | TFND | TFND | TFND |
| JS1008 | TFND | TFND | TFND | TFND |
| JS1009 | FTA | TFND | TFND | TFND |
| JS1010 | TFND | TFND | TFND | TFND |
| JS1011 | TFND | TFND | TFND | TFND |
| JS1012 | FTA | TFND | TFND | TFND |
| JS1013 | TFND | TFND | TFND | TFND |
| JS1014 | FTA | TFND | TFND | TFND |
| JS1015 | FTA | TFND | TFND | TFND |
| JS1016 | FTA | TFND | TFND | TFND |
| JS1017 | TFND | TFND | TFND | TFND |

| | | | | |
|---|---|---|---|---|
| JS1018 | TFND | TFND | TFND | TFND |
| JS1019 | FTA | TFND | TFND | TFND |
| JS1020 | TFND | TFND | TFND | TFND |
| JS1021 | FTA | TFND | TFND | TFND |
| JS1022 | FTA | TFND | TFND | TFND |
| JS1023 | HT | TFND | TFND | TFND |
| JS1024 | FTA | TFND | TFND | TFND |
| JS1025 | TFND | TFND | FTA | TFND |
| JS1026 | TFND | FTA | FTA | FTA |
| JS1027 | TFND | FTA | TFND | TFND |
| JS1028 | TFND | TFND | HT | TFND |
| JS1029 | TFND | TFND | TFND | TFND |
| JS1030 | TFND | TFND | TFND | TFND |
| JS1031 | TFND | FTA | TFND | TFND |
| JS1032 | TFND | TFND | TFND | TFND |
| JS1033 | FTA | FTA | FTA | FTA |
| JS1034 | TFND | TFND | FTA | TFND |
| JS1035 | FTA | TFND | TFND | TFND |
| JS1036 | TFND | TFND | TFND | TFND |
| JS1037 | TFND | TFND | TFND | TFND |
| JS1038 | TFND | TFND | FTA | TFND |
| JS1039 | TFND | TFND | FTA | TFND |
| JS1040 | FTA | TFND | TFND | TFND |
| JS1041 | TFND | TFND | FTA | TFND |
| JS1042 | HT | HT | TFND | HT |
| JS1043 | TFND | TFND | TFND | TFND |
| JS1044 | TFND | TFND | TFND | TFND |
| JS1045 | TFND | TFND | FTA | TFND |
| JS1046 | TFND | FTA | TFND | TFND |
| JS1047 | TFND | TFND | FTA | TFND |
| JS1048 | TFND | TFND | TFND | TFND |
| JS1049 | FTA | TFND | FTA | FTA |
| JS1050 | TFND | TFND | TFND | TFND |
| JS1051 | TFND | TFND | TFND | TFND |
| JS1052 | FTA | FTA | FTA | FTA |
| JS1053 | TFND | TFND | FTA | TFND |
| JS1054 | TFND | TFND | TFND | TFND |
| JS1055 | TFND | TFND | TFND | TFND |
| JS1056 | TFND | TFND | TFND | TFND |
| JS1057 | FTA | FTA | FTA | FTA |
| JS1058 | TFND | TFND | TFND | TFND |
| JS1059 | TFND | FTA | TFND | TFND |
| JS1060 | TFND | TFND | TFND | TFND |
| JS1061 | TFND | TFND | TFND | TFND |
| JS1062 | TFND | TFND | TFND | TFND |
| JS1063 | TFND | TFND | TFND | TFND |

| | | | | |
|---|---|---|---|---|
| JS1064 | FTA | FTA | FTA | FTA |
| JS1065 | TFND | FTA | TFND | TFND |
| JS1066 | FTA | TFND | TFND | TFND |
| JS1067 | TFND | TFND | TFND | TFND |
| JS1068 | FTA | TFND | TFND | TFND |
| JS1069 | TFND | FTA | TFND | TFND |
| JS1070 | TFND | TFND | TFND | TFND |
| JS1071 | TFND | FTA | TFND | TFND |
| JS1072 | FTA | TFND | TFND | TFND |
| JS1073 | FTA | TFND | TFND | TFND |
| JS1074 | FTA | FTA | TFND | FTA |
| JS1075 | TFND | TFND | TFND | TFND |
| JS1076 | FTA | FTA | FTA | FTA |
| JS1077 | TFND | TFND | TFND | TFND |
| JS1078 | TFND | TFND | TFND | TFND |
| JS1079 | FTA | FTA | FTA | FTA |
| JS1080 | TFND | TFND | TFND | TFND |
| JS1081 | TFND | FTA | TFND | TFND |
| JS1082 | TFND | TFND | TFND | TFND |
| JS1083 | TFND | TFND | TFND | TFND |
| JS1084 | FTA | FTA | FTA | FTA |
| JS1085 | TFND | TFND | TFND | TFND |
| JS1086 | FTA | FTA | FTA | FTA |
| JS1087 | TFND | TFND | TFND | TFND |
| JS1088 | FTA | TFND | TFND | TFND |
| JS1089 | FTA | TFND | TFND | TFND |
| JS1090 | TFND | TFND | TFND | TFND |
| JS1091 | TFND | TFND | TFND | TFND |
| JS1092 | FTA | TFND | TFND | TFND |
| JS1093 | FTA | TFND | TFND | TFND |
| JS1094 | TFND | TFND | TFND | TFND |
| JS1095 | TFND | TFND | TFND | TFND |
| JS1096 | FTA | FTA | FTA | FTA |
| JS1097 | FTA | TFND | TFND | TFND |
| JS1098 | TFND | TFND | TFND | TFND |
| JS1099 | TFND | FTA | TFND | TFND |
| JS1100 | TFND | TFND | TFND | TFND |
| JS1101 | TFND | TFND | TFND | TFND |
| JS1102 | TFND | TFND | TFND | TFND |
| JS1103 | FTA | TFND | TFND | TFND |
| JS1104 | FTA | TFND | TFND | TFND |
| JS1105 | FTA | TFND | TFND | TFND |
| JS1106 | TFND | TFND | TFND | TFND |
| JS1107 | FTA | FTA | FTA | FTA |
| JS1108 | TFND | FTA | TFND | TFND |
| JS1109 | TFND | TFND | TFND | TFND |

| | | | | |
|---|---|---|---|---|
| JS1110 | TFND | TFND | TFND | TFND |
| JS1111 | FTA | FTA | FTA | FTA |
| JS1112 | TFND | TFND | TFND | TFND |
| JS1113 | FTA | FTA | FTA | FTA |
| JS1114 | FTA | TFND | TFND | TFND |
| JS1115 | TFND | TFND | TFND | TFND |
| JS1116 | FTA | TFND | TFND | TFND |
| JS1117 | TFND | TFND | TFND | TFND |
| SH0007 | PTC | PTC | PTC | PTC |
| SH0014 | PTC | PTC | PTC | PTC |
| SH0017 | PTC | FTC | PTC | PTC |
| SH0028 | PTC | PTC | PTC | PTC |
| SH0030 | PTC | PTC | PTC | PTC |
| SH0034 | PTC | PTC | PTC | PTC |
| SH0036 | PTC | PTC | PTC | PTC |
| SH0037 | PTC | PTC | PTC | PTC |
| SH0043 | PTC | PTC | PTC | PTC |
| SH0046 | PTC | PTC | PTC | PTC |
| SH0053 | FTA | PTC | PTC | PTC |
| SH0061 | PTC | PTC | PTC | PTC |
| SH0064 | PTC | PTC | PTC | PTC |
| SH0069 | PTC | PTC | PTC | PTC |
| SH0071 | PTC | FTC | PTC | PTC |
| SH0080 | PTC | PTC | PTC | PTC |
| SH0087 | PTC | PTC | PTC | PTC |
| SH0099 | PTC | PTC | PTC | PTC |
| SH0104 | PTC | PTC | PTC | PTC |
| SH0105 | PTC | PTC | PTC | PTC |
| SH0108 | PTC | PTC | PTC | PTC |
| SH0114 | PTC | PTC | PTC | PTC |
| SH0124 | PTC | PTC | PTC | PTC |
| SH0126 | PTC | FTA | PTC | PTC |
| SH0127 | PTC | FTA | PTC | PTC |
| SH0129 | PTC | PTC | PTC | PTC |
| SH0131 | PTC | PTC | PTC | PTC |
| SH0132 | PTC | FTC | PTC | PTC |
| SH0133 | PTC | PTC | PTC | PTC |
| SH0134 | PTC | PTC | PTC | PTC |
| SH0135 | PTC | PTC | PTC | PTC |
| SH0136 | PTC | PTC | PTC | PTC |
| SH0141 | PTC | PTC | PTC | PTC |
| SH0146 | PTC | PTC | PTC | PTC |
| SH0149 | PTC | PTC | PTC | PTC |
| SH0151 | PTC | PTC | PTC | PTC |
| SH0157 | PTC | PTC | PTC | PTC |
| SH0158 | PTC | PTC | TFND | PTC |

| | | | | |
|---|---|---|---|---|
| SH0159 | PTC | PTC | PTC | PTC |
| SH0161 | PTC | PTC | PTC | PTC |
| SH0166 | PTC | PTC | TFND | PTC |
| SH0170 | PTC | PTC | PTC | PTC |
| SH0173 | PTC | PTC | PTC | PTC |
| SH0174 | PTC | PTC | PTC | PTC |
| SH0177 | PTC | PTC | PTC | PTC |
| SH0180 | FTA | PTC | PTC | PTC |
| SH0181 | PTC | PTC | PTC | PTC |
| SH0183 | PTC | PTC | PTC | PTC |
| SH0186 | PTC | PTC | PTC | PTC |
| SH0190 | PTC | PTC | PTC | PTC |
| SH0196 | FTC | PTC | PTC | PTC |
| SH0198 | PTC | PTC | PTC | PTC |
| SH0212 | PTC | PTC | TFND | PTC |
| JS2002 | PTC | FTC | PTC | PTC |
| JS2003 | FTC | PTC | FTC | FTC |
| JS2006 | PTC | FTA | PTC | PTC |
| JS2007 | PTC | PTC | FTC | PTC |
| JS2009 | PTC | PTC | PTC | PTC |
| JS2011 | PTC+OCA | FTC | PTC+OCA | PTC+OCA |
| JS2012 | PTC | FTC | PTC | PTC |
| JS2015 | PTC | PTC | PTC | PTC |
| JS2017 | FTC | PTC | PTC | PTC |
| JS2020 | FTA | PTC | PTC | PTC |
| JS2021 | PTC | PTC | PTC | PTC |
| JS2034 | PTC | FTC | PTC | PTC |
| JS2035 | PTC | PTC | PTC | PTC |
| JS2038 | PTC | PTC | PTC | PTC |
| JS2045 | FTC | PTC | PTC | PTC |
| JS2047 | PTC | PTC | PTC | PTC |
| JS2050 | FTA | PTC | PTC | PTC |
| JS2055 | PTC | PTC | PTC | PTC |
| JS2059 | PTC | PTC | PTC | PTC |
| JS2060 | OCC | PTC | OCC | OCC |
| JS2064 | PTC | PTC | PTC | PTC |
| JS2066 | PTC | PTC | PTC | PTC |
| JS2068 | PTC | PTC | PTC | PTC |
| JS2072 | FTC | PTC | PTC | PTC |
| JS2073 | PTC | FTA | PTC | PTC |
| JS2079 | PTC | PTC | PTC | PTC |
| JS2080 | PTC | PTC | FTA | PTC |
| JS2081 | PTC | PTC | PTC | PTC |
| JS2082 | PTC | PTC | FTC | PTC |
| JS2088 | PTC | PTC | TFND | PTC |
| JS2097 | OCC | OCC | FTA | OCC |

| | | | | |
|---|---|---|---|---|
| JS2109 | PTC | PTC | PTC | PTC |
| JS2116 | PTC | PTC | PTC | PTC |
| JS2120 | PTC | PTC | PTC | PTC |
| JS2122 | PTC | TFND | PTC | PTC |
| JS2124 | PTC | PTC | PTC | PTC |
| JS2126 | PTC | FTC | PTC | PTC |
| JS2138 | FTC | PTC | PTC | PTC |
| JS2142 | PTC | FTA | PTC | PTC |
| JS2144 | PTC | PTC | PTC | PTC |
| JS2145 | PTC | FTC | PTC | PTC |
| JS2148 | PTC | PTC | PTC | PTC |
| JS2152 | PTC | PTC | PTC | PTC |
| JS2159 | PTC | PTC | TFND | PTC |
| JS2186 | PTC | PTC | PTC | PTC |
| JS2189 | FTC | PTC | PTC | PTC |
| JS2190 | PTC | PTC | PTC | PTC |
| JS2193 | PTC | FTC | PTC | PTC |
| JS2196 | PTC | PTC | PTC | PTC |
| JS2200 | PTC | PTC | PTC | PTC |
| JS3001 | FTA | FTA | TFND | FTA |
| JS3002 | FTA | FTA | TFND | FTA |
| JS3004 | FTA | TFND | TFND | TFND |
| JS3006 | FTA | FTA | HT | FTA |
| JS3008 | TFND | FTA | TFND | TFND |
| JS3009 | FTA | FTA | TFND | FTA |
| JS3010 | FTA | TFND | TFND | TFND |
| JS3012 | FTA | TFND | TFND | TFND |
| JS3013 | TFND | HT | TFND | TFND |
| JS3014 | FTA | FTA | TFND | FTA |
| JS3015 | FTA | FTA | FTA | FTA |
| JS3017 | FTA | FTA | TFND | FTA |
| JS3018 | FTA | TFND | TFND | TFND |
| JS3019 | FTA | FTA | TFND | FTA |
| JS3020 | TFND | TFND | TFND | TFND |
| JS3021 | FTA | TFND | TFND | TFND |
| JS3022 | FTA | FTA | TFND | FTA |
| JS3023 | TFND | TFND | TFND | TFND |
| JS3024 | TFND | HT | TFND | TFND |
| JS3025 | TFND | TFND | FTA | TFND |
| JS3026 | FTA | TFND | FTA | FTA |
| JS3028 | FTA | TFND | FTA | FTA |
| JS3030 | FTA | FTA | FTA | FTA |
| JS3031 | FTA | FTA | TFND | FTA |
| JS3033 | TFND | FTA | TFND | TFND |
| JS3034 | FTA | FTA | FTA | FTA |
| JS3036 | TFND | FTA | TFND | TFND |

| | | | | |
|---|---|---|---|---|
| JS3037 | FTA | FTA | TFND | FTA |
| JS3038 | FTA | TFND | TFND | TFND |
| JS3039 | HT | TFND | TFND | TFND |
| JS3041 | FTA | FTA | TFND | FTA |
| JS3042 | TFND | FTA | FTA | FTA |
| JS3043 | TFND | FTA | TFND | TFND |
| JS3045 | FTA | TFND | FTA | FTA |
| JS3046 | FTA | TFND | TFND | TFND |
| JS3048 | FTA | FTA | TFND | FTA |
| JS3050 | TFND | FTA | FTA | FTA |
| JS3051 | FTA | FTA | FTA | FTA |
| JS3053 | TFND | TFND | TFND | TFND |
| JS3054 | FTA | FTA | TFND | FTA |
| JS3056 | FTA | TFND | FTA | FTA |
| JS3057 | TFND | FTA | TFND | TFND |
| JS3058 | TFND | FTA | TFND | TFND |
| JS3059 | FTA | FTA | TFND | FTA |
| JS3060 | FTA | TFND | TFND | TFND |
| JS3061 | FTA | HT | FTA | FTA |
| JS3063 | FTA | TFND | TFND | TFND |
| JS3064 | TFND | FTA | FTA | FTA |
| JS3066 | HT | FTA | FTA | FTA |
| JS3067 | FTA | TFND | TFND | TFND |
| JS3068 | FTA | TFND | TFND | TFND |
| JS3069 | TFND | FTA | TFND | TFND |
| JS3071 | FTA | TFND | FTA | FTA |
| JS3072 | FTA | FTA | TFND | FTA |
| JS3073 | FTA | TFND | FTA | FTA |
| JS3074 | TFND | FTA | TFND | TFND |
| JS3075 | FTA | TFND | FTA | FTA |
| JS3076 | TFND | TFND | HT | TFND |
| JS3077 | FTA | TFND | FTA | FTA |
| JS3078 | TFND | FTA | TFND | TFND |

TFND, Thyroid follicular nodular disease.
HT, Hashimoto's thyroiditis.
FTA, follicular thyroid adenoma.
FTC, follicular thyroid carcinoma.
PTC, papillary thyroid carcinoma.
OCC, oncocytic thyroid cancer
OCA, oncocytic thyroid adenoma

**Supplementary Table S4.** Independent histopathological review results of the second validation cohort (FFPE specimens) by three senior thyroid pathologists

| Case ID | Histology | | | Final Pathology Diagnosis |
|---|---|---|---|---|
| | Dr. Wu | Dr. Zhu | Dr. Zhang | |
| JS6718 | FTC | FTC | PTC | FTC |
| JS6845 | FTC | PTC | FTC | FTC |
| JS7393 | FTC | FTC | PTC | FTC |
| JS6889 | OCC | OCC | OCC | OCC |
| JS7333 | PTC | PTC | PTC | PTC |
| JS7341 | PTC | PTC | PTC | PTC |
| JS7360 | PTC | PTC | PTC | PTC |
| JS7198 | PTC | PTC | PTC | PTC |
| JS7340 | FTC | PTC | PTC | PTC |
| JS7344 | PTC | PTC | PTC | PTC |
| JS6749 | PTC | PTC | PTC | PTC |
| JS7102 | PTC | PTC | PTC | PTC |
| JS7380 | FTC | PTC | FTC | PTC |
| JS7335 | PTC | PTC | PTC | PTC |
| JS7106 | PTC | PTC | PTC | PTC |
| JS7257 | PTC | PTC | PTC | PTC |
| JS7332 | PTC | PTC | PTC | PTC |
| JS6714 | PTC | PTC | PTC | PTC |
| JS7465 | PTC | PTC | PTC | PTC |
| JS7031 | PTC | PTC | PTC | PTC |
| JS7372 | PTC | PTC | PTC | PTC |
| JS7347 | PTC | PTC | PTC | PTC |
| JS7343 | PTC | PTC | PTC | PTC |
| JS7427 | FTC | PTC | PTC | PTC |
| JS7058 | PTC | PTC | PTC | PTC |
| JS7354 | PTC | PTC | PTC | PTC |
| JS7256 | PTC | PTC | PTC | PTC |
| JS7339 | PTC | FTA | PTC | PTC |
| JS7385 | PTC | PTC | PTC | PTC |
| JS6896 | PTC | PTC | PTC | PTC |
| JS7035 | PTC | PTC | PTC | PTC |
| JS7361 | PTC | FTC | PTC | PTC |
| JS6793 | PTC | PTC | PTC | PTC |
| JS6696 | PTC | PTC | PTC | PTC |
| JS7437 | PTC | PTC | PTC | PTC |
| JS7008 | PTC | PTC | PTC | PTC |
| JS7060 | PTC | PTC | PTC | PTC |
| JS7228 | PTC | FTC | PTC | PTC |
| JS6794 | PTC | PTC | PTC | PTC |
| JS6707 | PTC | PTC | PTC | PTC |
| JS7412 | FTC | PTC | PTC | PTC |

| | | | | |
|---|---|---|---|---|
| JS7388 | PTC | PTC | PTC | PTC |
| JS7287 | PTC | PTC | PTC | PTC |
| JS7290 | PTC | PTC | PTC | PTC |
| JS6725 | PTC | PTC | PTC | PTC |
| JS6859 | PTC | PTC | PTC | PTC |
| JS6702 | PTC | PTC | PTC | PTC |
| JS6704 | PTC | PTC | PTC | PTC |
| JS7196 | PTC | PTC | PTC | PTC |
| JS6864 | PTC | PTC | PTC | PTC |
| JS7357 | PTC | PTC | PTC | PTC |
| JS7055 | PTC | PTC | PTC | PTC |
| JS6856 | PTC | PTC | PTC | PTC |
| JS6709 | PTC | PTC | PTC | PTC |
| JS7186 | PTC | PTC | PTC | PTC |
| JS6703 | PTC | PTC | PTC | PTC |
| JS6720 | PTC | PTC | PTC | PTC |
| JS6772 | PTC | PTC | PTC | PTC |
| JS6706 | PTC | PTC | PTC | PTC |
| JS7202 | PTC | PTC | PTC | PTC |
| JS7337 | PTC | PTC | PTC | PTC |
| JS6719 | PTC | PTC | PTC | PTC |
| JS7326 | PTC | PTC | PTC | PTC |
| JS6729 | PTC | PTC | PTC | PTC |
| JS6721 | PTC | PTC | PTC | PTC |
| JS7325 | PTC | PTC | PTC | PTC |
| JS6726 | PTC | PTC | PTC | PTC |
| JS7330 | PTC | PTC | PTC | PTC |
| JS6803 | PTC | PTC | PTC | PTC |
| JS7036 | PTC | PTC | PTC | PTC |
| JS7323 | PTC | PTC | PTC | PTC |
| JS7336 | PTC | PTC | PTC | PTC |
| JS7334 | PTC | PTC | PTC | PTC |
| JS6727 | PTC | PTC | PTC | PTC |
| JS6776 | PTC | PTC | PTC | PTC |
| JS7359 | PTC | PTC | PTC | PTC |
| JS6732 | PTC | PTC | PTC | PTC |
| JS6766 | PTC | PTC | PTC | PTC |
| JS7399 | PTC | PTC | PTC | PTC |
| JS6731 | PTC | PTC | PTC | PTC |
| JS6765 | PTC | PTC | PTC | PTC |
| JS6746 | PTC | PTC | PTC | PTC |
| JS6739 | PTC | PTC | PTC | PTC |
| JS6753 | PTC | PTC | PTC | PTC |
| JS7400 | PTC | PTC | PTC | PTC |
| JS6800 | PTC | PTC | PTC | PTC |
| JS7396 | PTC | PTC | PTC | PTC |

| | | | | |
|---|---|---|---|---|
| JS6773 | PTC | PTC | PTC | PTC |
| JS6779 | PTC | PTC | PTC | PTC |
| JS6758 | PTC | PTC | PTC | PTC |
| JS7387 | PTC | FTC | PTC | PTC |
| JS7066 | PTC | PTC | PTC | PTC |
| JS7028 | PTC | PTC | PTC | PTC |
| JS7379 | PTC | PTC | PTC | PTC |
| JS7391 | PTC | PTC | PTC | PTC |
| JS7371 | PTC | PTC | PTC | PTC |
| JS7370 | PTC | FTA | PTC | PTC |
| JS6874 | PTC | PTC | PTC | PTC |
| JS6770 | PTC | PTC | PTC | PTC |
| JS7462 | PTC | PTC | PTC | PTC |
| JS6785 | PTC | PTC | PTC | PTC |
| JS6796 | PTC | PTC | PTC | PTC |
| JS7390 | PTC | PTC | PTC | PTC |
| JS6862 | PTC | PTC | PTC | PTC |
| JS6762 | PTC | PTC | PTC | PTC |
| JS7098 | PTC | PTC | PTC | PTC |
| JS6812 | PTC | PTC | PTC | PTC |
| JS7038 | PTC | PTC | PTC | PTC |
| JS6810 | PTC | PTC | PTC | PTC |
| JS7295 | PTC | PTC | PTC | PTC |
| JS7402 | PTC | PTC | PTC | PTC |
| JS7005 | PTC | PTC | FTC | PTC |
| JS6821 | PTC | PTC | PTC | PTC |
| JS7409 | PTC | PTC | PTC | PTC |
| JS6870 | PTC | PTC | PTC | PTC |
| JS6830 | PTC | PTC | PTC | PTC |
| JS7419 | PTC | PTC | PTC | PTC |
| JS7397 | PTC | PTC | PTC | PTC |
| JS6835 | PTC | PTC | PTC | PTC |
| JS7431 | PTC | PTC | PTC | PTC |
| JS7413 | PTC | PTC | PTC | PTC |
| JS7439 | PTC | PTC | PTC | PTC |
| JS7414 | PTC | PTC | PTC | PTC |
| JS7020 | PTC | PTC | PTC | PTC |
| JS6807 | PTC | PTC | PTC | PTC |
| JS7423 | PTC | PTC | PTC | PTC |
| JS7416 | PTC | PTC | PTC | PTC |
| JS6841 | FTC | PTC | PTC | PTC |
| JS7424 | PTC | PTC | PTC | PTC |
| JS6829 | PTC | PTC | PTC | PTC |
| JS6831 | PTC | PTC | PTC | PTC |
| JS6861 | PTC | PTC | PTC | PTC |
| JS7438 | PTC | PTC | PTC | PTC |

| | | | | |
|---|---|---|---|---|
| JS7443 | PTC | PTC | PTC | PTC |
| JS6846 | PTC | PTC | PTC | PTC |
| JS6865 | PTC | PTC | PTC | PTC |
| JS7433 | PTC | PTC | PTC | PTC |
| JS7442 | PTC | PTC | PTC | PTC |
| JS6887 | PTC | FTC | PTC | PTC |
| JS7173 | PTC | PTC | PTC | PTC |
| JS7444 | PTC | PTC | PTC | PTC |
| JS6872 | PTC | PTC | PTC | PTC |
| JS6882 | PTC | PTC | PTC | PTC |
| JS7455 | PTC | PTC | PTC | PTC |
| JS7441 | PTC | PTC | PTC | PTC |
| JS7440 | PTC | PTC | PTC | PTC |
| JS6885 | PTC | PTC | PTC | PTC |
| JS6873 | PTC | PTC | PTC | PTC |
| JS7014 | PTC | PTC | PTC | PTC |
| JS6857 | PTC | PTC | PTC | PTC |
| JS7459 | PTC | PTC | PTC | PTC |
| JS7428 | PTC | PTC | PTC | PTC |
| JS6843 | PTC | FTC | PTC | PTC |
| JS7006 | PTC | PTC | PTC | PTC |
| JS6888 | PTC | PTC | PTC | PTC |
| JS7457 | PTC | PTC | PTC | PTC |
| JS6879 | PTC | PTC | PTC | PTC |
| JS7160 | PTC | PTC | PTC | PTC |
| JS7075 | PTC | PTC | PTC | PTC |
| JS7458 | PTC | PTC | PTC | PTC |
| JS6895 | PTC | PTC | FTC | PTC |
| JS7461 | PTC | PTC | PTC | PTC |
| JS7467 | PTC | PTC | PTC | PTC |
| JS7013 | PTC | PTC | PTC | PTC |
| JS7456 | PTC | PTC | PTC | PTC |
| JS7466 | PTC | PTC | PTC | PTC |
| JS7451 | PTC | PTC | PTC | PTC |
| JS7023 | PTC | PTC | PTC | PTC |
| JS7100 | PTC | PTC | PTC | PTC |
| JS7464 | PTC | PTC | PTC | PTC |
| JS7003 | PTC | PTC | PTC | PTC |
| JS7092 | PTC | PTC | PTC | PTC |
| JS6880 | PTC | PTC | FTA | PTC |
| JS7135 | PTC | PTC | PTC | PTC |
| JS7163 | PTC | PTC | PTC | PTC |
| JS6893 | PTC | PTC | PTC | PTC |
| JS7112 | PTC | PTC | PTC | PTC |
| JS6824 | PTC | PTC | PTC | PTC |
| JS7304 | FTC | PTC | PTC | PTC |

| | | | | |
|---|---|---|---|---|
| JS7000 | PTC | PTC | PTC | PTC |
| JS7001 | PTC | PTC | PTC | PTC |
| JS7128 | PTC | PTC | PTC | PTC |
| JS7171 | PTC | PTC | PTC | PTC |
| JS6838 | PTC | FTC | PTC | PTC |
| JS7039 | PTC | PTC | PTC | PTC |
| JS6790 | PTC | PTC | PTC | PTC |
| JS7021 | PTC | PTC | PTC | PTC |
| JS7108 | PTC | PTC | PTC | PTC |
| JS7011 | PTC | PTC | PTC | PTC |
| JS7082 | PTC | PTC | PTC | PTC |
| JS7017 | PTC | PTC | PTC | PTC |
| JS7015 | PTC | PTC | PTC | PTC |
| JS7084 | PTC | PTC | PTC | PTC |
| JS7012 | PTC | PTC | PTC | PTC |
| JS6877 | PTC | PTC | PTC | PTC |
| JS7080 | PTC | PTC | FTA | PTC |
| JS6849 | PTC | PTC | PTC | PTC |
| JS7276 | PTC | PTC | PTC | PTC |
| JS7032 | PTC | PTC | PTC | PTC |
| JS6822 | PTC | PTC | PTC | PTC |
| JS7029 | PTC | PTC | PTC | PTC |
| JS7096 | PTC | PTC | PTC | PTC |
| JS7142 | PTC | PTC | PTC | PTC |
| JS7030 | PTC | PTC | PTC | PTC |
| JS7166 | PTC | PTC | PTC | PTC |
| JS7141 | PTC | PTC | PTC | PTC |
| JS7043 | PTC | PTC | PTC | PTC |
| JS7051 | PTC | FTC | PTC | PTC |
| JS7057 | PTC | PTC | PTC | PTC |
| JS7168 | PTC | PTC | PTC | PTC |
| JS7049 | PTC | PTC | PTC | PTC |
| JS7047 | PTC | PTC | PTC | PTC |
| JS7059 | PTC | PTC | PTC | PTC |
| JS7054 | PTC | PTC | PTC | PTC |
| JS6715 | PTC | PTC | PTC | PTC |
| JS7056 | PTC | PTC | PTC | PTC |
| JS7045 | PTC | PTC | PTC | PTC |
| JS7064 | PTC | PTC | PTC | PTC |
| JS7076 | PTC | PTC | PTC | PTC |
| JS7063 | FTC | PTC | PTC | PTC |
| JS7078 | PTC | PTC | PTC | PTC |
| JS7072 | PTC | PTC | PTC | PTC |
| JS7105 | PTC | PTC | PTC | PTC |
| JS7191 | PTC | PTC | PTC | PTC |
| JS7081 | PTC | PTC | PTC | PTC |

| | | | | |
|---|---|---|---|---|
| JS7429 | PTC | PTC | PTC | PTC |
| JS7104 | FTC | PTC | PTC | PTC |
| JS7205 | PTC | PTC | PTC | PTC |
| JS7086 | PTC | PTC | PTC | PTC |
| JS7149 | PTC | PTC | PTC | PTC |
| JS7074 | PTC | PTC | PTC | PTC |
| JS7073 | PTC | PTC | PTC | PTC |
| JS7383 | PTC | PTC | PTC | PTC |
| JS7127 | PTC | PTC | PTC | PTC |
| JS7194 | PTC | PTC | PTC | PTC |
| JS7124 | PTC | PTC | PTC | PTC |
| JS7150 | PTC | PTC | PTC | PTC |
| JS7303 | PTC | PTC | PTC | PTC |
| JS7174 | PTC | PTC | PTC | PTC |
| JS7085 | PTC | PTC | PTC | PTC |
| JS7161 | PTC | PTC | PTC | PTC |
| JS7348 | PTC | FTC | PTC | PTC |
| JS7176 | PTC | PTC | PTC | PTC |
| JS7222 | PTC | PTC | PTC | PTC |
| JS7101 | PTC | PTC | PTC | PTC |
| JS7219 | PTC | PTC | PTC | PTC |
| JS7181 | PTC | PTC | PTC | PTC |
| JS7258 | PTC | PTC | PTC | PTC |
| JS7145 | PTC | PTC | PTC | PTC |
| JS7201 | PTC | PTC | PTC | PTC |
| JS7210 | FTA | PTC | PTC | PTC |
| JS7212 | PTC | PTC | PTC | PTC |
| JS7182 | PTC | PTC | PTC | PTC |
| JS7099 | PTC | PTC | PTC | PTC |
| JS7218 | PTC | PTC | PTC | PTC |
| JS7300 | PTC | PTC | PTC | PTC |
| JS7133 | PTC | PTC | PTC | PTC |
| JS7240 | PTC | PTC | PTC | PTC |
| JS7324 | PTC | PTC | PTC | PTC |
| JS7126 | PTC | PTC | FTC | PTC |
| JS7175 | PTC | PTC | PTC | PTC |
| JS7144 | PTC | PTC | PTC | PTC |
| JS7192 | PTC | PTC | PTC | PTC |
| JS7398 | PTC | PTC | PTC | PTC |
| JS7265 | PTC | FTC | PTC | PTC |
| JS7244 | PTC | PTC | PTC | PTC |
| JS7190 | PTC | PTC | PTC | PTC |
| JS7234 | PTC | PTC | PTC | PTC |
| JS7157 | PTC | PTC | PTC | PTC |
| JS7233 | PTC | PTC | PTC | PTC |
| JS7291 | PTC | PTC | PTC | PTC |

| | | | | |
|---|---|---|---|---|
| JS7356 | PTC | PTC | PTC | PTC |
| JS6866 | FTC | PTC | PTC | PTC |
| JS7236 | PTC | PTC | PTC | PTC |
| JS7260 | PTC | PTC | PTC | PTC |
| JS7329 | PTC | PTC | PTC | PTC |
| JS7262 | PTC | PTC | PTC | PTC |
| JS7268 | PTC | PTC | PTC | PTC |
| JS7285 | PTC | PTC | PTC | PTC |
| JS7273 | PTC | FTC | PTC | PTC |
| JS7283 | PTC | PTC | PTC | PTC |
| JS7050 | PTC | PTC | PTC | PTC |
| JS7288 | PTC | PTC | PTC | PTC |
| JS7292 | PTC | PTC | PTC | PTC |
| JS6850 | PTC | PTC | PTC | PTC |
| JS7280 | PTC | PTC | FTC | PTC |
| JS7286 | PTC | PTC | PTC | PTC |
| JS7284 | PTC | PTC | PTC | PTC |
| JS7318 | PTC | PTC | PTC | PTC |
| JS7293 | PTC | PTC | PTC | PTC |
| JS6797 | PTC | PTC | PTC | PTC |
| JS7382 | PTC | PTC | PTC | PTC |
| JS7121 | PTC | PTC | FTC | PTC |
| JS7314 | PTC | PTC | PTC | PTC |
| JS7316 | PTC | PTC | PTC | PTC |
| JS7107 | PTC | PTC | PTC | PTC |
| JS7179 | PTC | PTC | PTC | PTC |
| JS6734 | PTC | PTC | PTC | PTC |
| JS6808 | PTC | PTC | PTC | PTC |
| JS6802 | PTC | PTC | PTC | PTC |
| JS7230 | TFND | TFND | FTA | TFND |
| JS7071 | TFND | TFND | FTA | TFND |
| JS7193 | TFND | TFND | FTA | TFND |
| JS7252 | TFND | TFND | FTA | TFND |
| JS7432 | TFND | FTA | TFND | TFND |
| JS7069 | FTA | TFND | FTA | FTA |
| JS7245 | TFND | TFND | HT | TFND |
| JS7068 | TFND | TFND | FTA | TFND |
| JS7062 | TFND | TFND | FTA | TFND |
| JS7199 | TFND | FTA | TFND | TFND |
| JS7373 | FTA | TFND | TFND | TFND |
| JS7221 | HT | TFND | TFND | TFND |
| JS7046 | TFND | TFND | FTA | TFND |
| JS6828 | TFND | TFND | FTA | TFND |
| JS7177 | TFND | TFND | FTA | TFND |
| JS6858 | TFND | FTA | FTA | FTA |
| JS7041 | TFND | HT | TFND | TFND |

| | | | | |
|---|---|---|---|---|
| JS7195 | TFND | TFND | FTA | TFND |
| JS7067 | TFND | FTA | FTA | FTA |
| JS7362 | TFND | HT | TFND | TFND |
| JS6848 | TFND | FTA | TFND | TFND |
| JS6722 | TFND | TFND | TFND | TFND |
| JS6723 | TFND | HT | TFND | TFND |
| JS6799 | TFND | TFND | FTA | TFND |
| JS6742 | TFND | TFND | HT | TFND |
| JS6730 | HT | TFND | FTA | FTA |
| JS6713 | HT | TFND | FTA | FTA |
| JS6795 | FTA | FTA | FTA | FTA |
| JS6871 | FTA | TFND | FTA | FTA |
| JS7042 | HT | TFND | TFND | TFND |
| JS6712 | FTA | TFND | TFND | TFND |
| JS7040 | HT | TFND | TFND | TFND |
| JS7446 | TFND | TFND | HT | TFND |
| JS6701 | TFND | FTA | FTA | FTA |
| JS7211 | TFND | TFND | TFND | TFND |
| JS6842 | TFND | TFND | TFND | TFND |
| JS6875 | TFND | TFND | TFND | TFND |
| JS7033 | TFND | TFND | HT | TFND |
| JS6786 | TFND | FTA | TFND | TFND |
| JS6890 | TFND | FTA | FTA | FTA |
| JS6891 | TFND | FTA | FTA | FTA |
| JS7027 | TFND | TFND | TFND | TFND |
| JS6806 | TFND | TFND | FTA | TFND |
| JS7004 | TFND | TFND | HT | TFND |
| JS6740 | TFND | TFND | FTA | TFND |
| JS6804 | TFND | TFND | TFND | TFND |
| JS6741 | TFND | FTA | FTA | FTA |
| JS7165 | TFND | TFND | TFND | TFND |
| JS6833 | TFND | TFND | FTA | TFND |
| JS6811 | TFND | TFND | FTA | TFND |
| JS6697 | TFND | TFND | FTA | TFND |
| JS7425 | TFND | TFND | HT | TFND |
| JS7002 | TFND | TFND | FTA | TFND |
| JS7147 | TFND | TFND | TFND | TFND |
| JS7239 | FTA | FTA | TFND | FTA |
| JS7386 | FTA | FTA | TFND | FTA |
| JS6886 | FTA | FTA | TFND | FTA |
| JS7140 | TFND | FTA | FTA | FTA |
| JS6832 | FTA | FTA | FTA | FTA |
| JS7034 | TFND | TFND | TFND | TFND |
| JS7206 | TFND | TFND | HT | TFND |
| JS7024 | TFND | FTA | FTA | FTA |
| JS6894 | TFND | TFND | TFND | TFND |

| | | | | |
|---|---|---|---|---|
| JS7453 | FTA | FTA | HT | FTA |
| JS7103 | TFND | TFND | TFND | TFND |
| JS7255 | TFND | FTA | FTA | FTA |
| JS7282 | TFND | FTA | FTA | FTA |
| JS7200 | TFND | TFND | TFND | TFND |
| JS7129 | TFND | TFND | TFND | TFND |
| JS6744 | FTA | FTA | FTA | FTA |
| JS7213 | TFND | TFND | FTA | TFND |
| JS7153 | TFND | TFND | TFND | TFND |
| JS7154 | FTA | FTA | FTA | FTA |
| JS6855 | TFND | TFND | FTA | TFND |
| JS7411 | TFND | TFND | TFND | TFND |
| JS7355 | TFND | FTA | FTA | FTA |
| JS7421 | TFND | FTA | FTA | FTA |
| JS7130 | TFND | FTA | FTA | FTA |
| JS7238 | TFND | TFND | FTA | TFND |
| JS7272 | TFND | TFND | TFND | TFND |
| JS6809 | TFND | TFND | FTA | TFND |
| JS7172 | FTA | FTA | TFND | FTA |
| JS7317 | TFND | TFND | HT | TFND |
| JS7261 | FTA | FTA | TFND | FTA |
| JS7125 | TFND | TFND | TFND | TFND |
| JS7369 | TFND | FTA | FTA | FTA |
| JS7275 | TFND | FTA | FTA | FTA |
| JS6698 | FTA | FTA | FTA | FTA |
| JS6788 | FTA | FTA | FTA | FTA |
| JS7079 | TFND | FTA | FTA | FTA |
| JS7235 | FTA | FTA | FTA | FTA |
| JS7436 | FTA | FTA | FTA | FTA |
| JS7242 | TFND | TFND | TFND | TFND |
| JS7294 | TFND | TFND | FTA | TFND |
| JS7214 | TFND | TFND | FTA | TFND |
| JS6755 | FTA | FTA | FTA | FTA |
| JS7384 | TFND | TFND | TFND | TFND |
| JS7139 | FTA | FTA | FTA | FTA |
| JS6774 | TFND | TFND | FTA | TFND |

TFND, Thyroid follicular nodular disease.
HT, Hashimoto's thyroiditis.
FTA, follicular thyroid adenoma.
FTC, follicular thyroid carcinoma.
PTC, papillary thyroid carcinoma.
OCC, oncocytic thyroid cancer
OCA, oncocytic thyroid adenoma

**Supplementary Table S5.** Independent Bethesda cytological review results of FNAB thyroid specimens in the subjects with histopathologically confirmed diagnosis by three senior thyroid pathologists

| Case ID | Final Histology | Bethesda Cytological Categories | | | Final Bethesda Categories |
|---|---|---|---|---|---|
| | | Dr. Wu | Dr. Zhu | Dr. Wan | |
| AH0023 | PTC | PTC | SM | SM | SM |
| AH0057 | BTN | FN | FN | BTN | FN |
| AH0094 | BTN | FN | FN | BTN | FN |
| AH0106 | PTC | SM | AUS | SM | SM |
| AH0107 | PTC | PTC | SM | SM | SM |
| AH0111 | PTC | PTC | SM | SM | SM |
| AH0115 | PTC | SM | SM | SM | SM |
| AH0127 | PTC | PTC | PTC | SM | PTC |
| AH0134 | PTC | PTC | SM | SM | SM |
| AH0139 | PTC | SM | PTC | PTC | PTC |
| AH0156 | PTC | PTC | PTC | PTC | PTC |
| AH0164 | PTC | PTC | FN | PTC | PTC |
| AH0170 | PTC | AUS | AUS | FN | AUS |
| AH0174 | PTC | SM | SM | PTC | SM |
| AH0191 | PTC | AUS | FN | FN | FN |
| AH0197 | PTC | AUS | FN | FN | FN |
| AH0220 | PTC | SM | PTC | SM | SM |
| AH0222 | PTC | FN | BTN | FN | FN |
| AH0257 | PTC | FN | SM | SM | SM |
| AH0259 | PTC | SM | AUS | AUS | AUS |
| AH0261 | PTC | PTC | SM | SM | SM |
| AH0262 | PTC | SM | AUS | SM | SM |
| AH0264 | PTC | PTC | SM | SM | SM |
| AH0267 | PTC | PTC | PTC | PTC | PTC |
| AH0272 | PTC | PTC | SM | SM | SM |
| AH0275 | PTC | SM | SM | SM | SM |
| AH0279 | PTC | AUS | AUS | FN | AUS |
| AH0290 | PTC | PTC | PTC | AUS | PTC |
| AH0291 | PTC | SM | SM | AUS | SM |
| AH0300 | PTC | SM | SM | SM | SM |
| AH0312 | PTC | SM | SM | SM | SM |
| AH0313 | PTC | AUS | FN | FN | FN |
| AH0322 | PTC | AUS | AUS | FN | AUS |
| AH0325 | PTC | PTC | SM | SM | SM |
| AH0327 | PTC | PTC | SM | SM | SM |
| AH0331 | PTC | SM | AUS | SM | SM |
| AH0334 | PTC | SM | SM | SM | SM |
| AH0335 | PTC | AUS | AUS | FN | AUS |
| AH0336 | PTC | PTC | PTC | AUS | PTC |

| | | | | | |
|---|---|---|---|---|---|
| AH0337 | PTC | PTC | PTC | AUS | PTC |
| AH0340 | PTC | PTC | SM | PTC | PTC |
| AH0342 | PTC | AUS | BTN | BTN | BTN |
| AH0358 | PTC | PTC | PTC | PTC | PTC |
| AH0391 | BTN | AUS | BTN | BTN | BTN |
| JS0696 | BTN | AUS | FN | FN | FN |
| JS0699 | BTN | AUS | FN | FN | FN |
| JS0706 | BTN | AUS | FN | AUS | AUS |
| JS0718 | PTC | AUS | FN | FN | FN |
| JS0720 | PTC | AUS | AUS | FN | AUS |
| JS0723 | PTC | AUS | FN | AUS | AUS |
| JS0731 | PTC | PTC | SM | SM | SM |
| JS0737 | PTC | PTC | PTC | PTC | PTC |
| JS0746 | PTC | AUS | FN | FN | FN |
| JS0749 | PTC | PTC | SM | SM | SM |
| JS0750 | PTC | AUS | AUS | FN | AUS |
| JS0751 | PTC | SM | SM | SM | SM |
| JS0752 | PTC | PTC | PTC | SM | PTC |
| JS0758 | BTN | PTC | FN | PTC | PTC |
| JS0760 | BTN | PTC | SM | SM | SM |
| JS0763 | PTC | AUS | FN | AUS | AUS |
| JS0767 | BTN | AUS | PTC | PTC | PTC |
| JS0769 | PTC | PTC | SM | SM | SM |
| JS0770 | BTN | FN | AUS | FN | FN |
| JS0771 | PTC | AUS | FN | FN | FN |
| JS0786 | PTC | PTC | PTC | PTC | PTC |
| JS0787 | PTC | SM | SM | SM | SM |
| JS0788 | PTC | PTC | SM | SM | SM |
| JS0789 | PTC | SM | SM | SM | SM |
| JS0790 | PTC | AUS | FN | FN | FN |
| JS0791 | PTC | PTC | PTC | SM | PTC |
| JS0792 | PTC | PTC | PTC | PTC | PTC |
| JS0793 | PTC | PTC | PTC | PTC | PTC |
| JS0794 | FTC | FTC | FTC | FN | FTC |
| JS0795 | FTC | FTC | FN | FTC | FTC |
| JS0796 | PTC | PTC | PTC | AUS | PTC |
| JS0797 | PTC | AUS | AUS | FN | AUS |
| JS0798 | PTC | PTC | PTC | PTC | PTC |
| JS0799 | PTC | PTC | PTC | PTC | PTC |
| JS0800 | PTC | AUS | FN | AUS | AUS |
| JS0801 | PTC | PTC | PTC | SM | PTC |
| JS0802 | PTC | PTC | PTC | PTC | PTC |
| JS0803 | PTC | PTC | SM | SM | SM |
| JS0804 | PTC | AUS | FN | AUS | AUS |

| | | | | | |
|---|---|---|---|---|---|
| JS0805 | PTC | FN | FN | FN | FN |
| JS0806 | PTC | PTC | PTC | PTC | PTC |
| JS0807 | PTC | PTC | PTC | PTC | PTC |
| JS0808 | PTC | PTC | SM | SM | SM |
| JS0809 | PTC | BTN | FN | BTN | BTN |
| JS0810 | PTC | PTC | PTC | PTC | PTC |
| JS0811 | PTC | AUS | FN | FN | FN |
| JS0812 | PTC | BTN | BTN | AUS | BTN |
| JS0813 | PTC | PTC | PTC | PTC | PTC |
| JS0814 | PTC | FN | FN | FN | FN |
| JS0825 | PTC | PTC | SM | SM | SM |
| JS0828 | BTN | FN | PTC | PTC | PTC |
| JS0844 | PTC | PTC | PTC | FTC | PTC |
| JS0846 | PTC | PTC | PTC | AUS | PTC |
| JS0847 | PTC | SM | SM | SM | SM |
| JS0848 | PTC | PTC | PTC | PTC | PTC |
| JS0851 | PTC | AUS | AUS | FN | AUS |
| JS0852 | PTC | FN | AUS | FN | FN |
| JS0857 | PTC | PTC | PTC | PTC | PTC |
| JS0858 | PTC | FTC | PTC | PTC | PTC |
| JS0859 | PTC | SM | SM | PTC | SM |
| JS0860 | PTC | PTC | PTC | PTC | PTC |
| JS0861 | PTC | PTC | PTC | SM | PTC |
| JS0862 | PTC | SM | SM | SM | SM |
| JS0863 | PTC | AUS | FN | AUS | AUS |
| JS0865 | PTC | FN | FN | FN | FN |
| JS0867 | PTC | PTC | PTC | PTC | PTC |
| JS0868 | PTC | FTC | PTC | PTC | PTC |
| JS0869 | PTC | AUS | FN | AUS | AUS |
| JS0870 | PTC | PTC | PTC | SM | PTC |
| JS0871 | PTC | AUS | FN | FN | FN |
| JS0872 | PTC | PTC | PTC | PTC | PTC |
| JS0873 | PTC | BTN | AUS | BTN | BTN |
| JS0874 | PTC | BTN | BTN | FTC | BTN |
| JS0875 | PTC | SM | PTC | PTC | PTC |
| JS0876 | PTC | BTN | BTN | FN | BTN |
| JS0877 | PTC | AUS | FN | FN | FN |
| JS0878 | PTC | PTC | PTC | FN | PTC |
| JS0882 | PTC | FTC | PTC | PTC | PTC |
| JS0885 | PTC | PTC | PTC | PTC | PTC |
| JS0887 | PTC | PTC | PTC | FN | PTC |
| JS0888 | PTC | PTC | PTC | SM | PTC |
| JS0890 | PTC | PTC | SM | SM | SM |
| JS0991 | BTN | AUS | BTN | BTN | BTN |

| | | | | | |
|---|---|---|---|---|---|
| JS0992 | BTN | AUS | BTN | BTN | BTN |
| JS0993 | BTN | BTN | BTN | BTN | BTN |
| JS0994 | BTN | PTC | SM | SM | SM |
| JS0995 | BTN | BTN | FN | BTN | BTN |
| JS0996 | BTN | BTN | AUS | BTN | BTN |
| JS0997 | BTN | BTN | AUS | BTN | BTN |
| JS0998 | BTN | BTN | BTN | BTN | BTN |
| JS0999 | BTN | AUS | BTN | BTN | BTN |
| JS1000 | BTN | BTN | BTN | BTN | BTN |
| JS1001 | BTN | PTC | PTC | SM | PTC |
| JS1002 | BTN | AUS | FN | AUS | AUS |
| JS1003 | BTN | BTN | BTN | BTN | BTN |
| JS1004 | BTN | BTN | BTN | AUS | BTN |
| JS1005 | BTN | PTC | SM | SM | SM |
| JS1006 | BTN | AUS | AUS | AUS | AUS |
| JS1007 | BTN | BTN | AUS | BTN | BTN |
| JS1008 | BTN | BTN | AUS | BTN | BTN |
| JS1009 | BTN | BTN | BTN | BTN | BTN |
| JS1010 | BTN | BTN | BTN | FN | BTN |
| JS1011 | BTN | BTN | BTN | BTN | BTN |
| JS1012 | BTN | AUS | AUS | FN | AUS |
| JS1013 | BTN | AUS | BTN | BTN | BTN |
| JS1014 | BTN | BTN | BTN | BTN | BTN |
| JS1015 | BTN | AUS | BTN | BTN | BTN |
| JS1016 | BTN | BTN | BTN | BTN | BTN |
| JS1017 | BTN | BTN | FN | BTN | BTN |
| JS1018 | BTN | BTN | BTN | BTN | BTN |
| JS1020 | BTN | BTN | BTN | BTN | BTN |
| JS1021 | BTN | AUS | FN | FN | FN |
| JS1022 | BTN | BTN | BTN | FN | BTN |
| JS1023 | BTN | AUS | AUS | FN | AUS |
| JS1024 | BTN | PTC | SM | SM | SM |
| JS1025 | BTN | AUS | AUS | FN | AUS |
| JS1026 | BTN | AUS | BTN | BTN | BTN |
| JS1027 | BTN | BTN | FN | BTN | BTN |
| JS1028 | BTN | BTN | FN | BTN | BTN |
| JS1029 | BTN | FN | BTN | BTN | BTN |
| JS1030 | BTN | AUS | AUS | AUS | AUS |
| JS1031 | BTN | AUS | BTN | BTN | BTN |
| JS1032 | BTN | BTN | BTN | BTN | BTN |
| JS1033 | BTN | FN | BTN | BTN | BTN |
| JS1034 | BTN | FN | BTN | BTN | BTN |
| JS1035 | BTN | AUS | AUS | FN | AUS |
| JS1036 | BTN | AUS | BTN | BTN | BTN |

| | | | | | |
|---|---|---|---|---|---|
| JS1037 | BTN | BTN | BTN | BTN | BTN |
| JS1038 | BTN | AUS | AUS | AUS | AUS |
| JS1039 | BTN | FN | BTN | BTN | BTN |
| JS1040 | BTN | BTN | BTN | BTN | BTN |
| JS1041 | BTN | BTN | BTN | FN | BTN |
| JS1042 | BTN | PTC | PTC | SM | PTC |
| JS1043 | BTN | BTN | BTN | BTN | BTN |
| JS1044 | BTN | BTN | BTN | FN | BTN |
| JS1045 | BTN | BTN | BTN | BTN | BTN |
| JS1046 | BTN | AUS | BTN | BTN | BTN |
| JS1047 | BTN | BTN | BTN | BTN | BTN |
| JS1048 | BTN | AUS | BTN | BTN | BTN |
| JS1049 | BTN | AUS | BTN | BTN | BTN |
| JS1050 | BTN | BTN | BTN | BTN | BTN |
| JS1051 | BTN | BTN | BTN | BTN | BTN |
| JS1052 | BTN | BTN | SM | BTN | BTN |
| JS1053 | BTN | AUS | BTN | BTN | BTN |
| JS1054 | BTN | AUS | AUS | FN | AUS |
| JS1055 | BTN | BTN | AUS | BTN | BTN |
| JS1056 | BTN | BTN | BTN | BTN | BTN |
| JS1057 | BTN | AUS | FN | AUS | AUS |
| JS1058 | BTN | BTN | BTN | BTN | BTN |
| JS1059 | BTN | AUS | BTN | BTN | BTN |
| JS1060 | BTN | BTN | BTN | BTN | BTN |
| JS1061 | BTN | FN | BTN | BTN | BTN |
| JS1062 | BTN | BTN | BTN | BTN | BTN |
| JS1063 | BTN | AUS | BTN | BTN | BTN |
| JS1064 | BTN | BTN | BTN | BTN | BTN |
| JS1065 | BTN | BTN | BTN | FN | BTN |
| JS1066 | BTN | BTN | BTN | BTN | BTN |
| JS1067 | BTN | BTN | BTN | FN | BTN |
| JS1068 | BTN | AUS | AUS | FN | AUS |
| JS1069 | BTN | BTN | BTN | FN | BTN |
| JS1070 | BTN | BTN | BTN | BTN | BTN |
| JS1071 | BTN | BTN | BTN | BTN | BTN |
| JS1072 | BTN | AUS | AUS | FN | AUS |
| JS1073 | BTN | BTN | BTN | BTN | BTN |
| JS1074 | BTN | AUS | AUS | FN | AUS |
| JS1075 | BTN | BTN | BTN | BTN | BTN |
| JS1076 | BTN | BTN | BTN | BTN | BTN |
| JS1077 | BTN | BTN | BTN | BTN | BTN |
| JS1078 | BTN | AUS | AUS | FN | AUS |
| JS1079 | BTN | BTN | BTN | BTN | BTN |
| JS1080 | BTN | AUS | BTN | BTN | BTN |

| | | | | | |
|---|---|---|---|---|---|
| JS1081 | BTN | BTN | BTN | BTN | BTN |
| JS1082 | BTN | FN | FN | FN | FN |
| JS1083 | BTN | BTN | BTN | BTN | BTN |
| JS1084 | BTN | FN | FN | FN | FN |
| JS1085 | BTN | PTC | SM | SM | SM |
| JS1086 | BTN | AUS | BTN | BTN | BTN |
| JS1087 | BTN | BTN | BTN | BTN | BTN |
| JS1088 | BTN | BTN | BTN | AUS | BTN |
| JS1089 | BTN | BTN | BTN | BTN | BTN |
| JS1090 | BTN | BTN | BTN | BTN | BTN |
| JS1091 | BTN | BTN | BTN | AUS | BTN |
| JS1092 | BTN | BTN | BTN | BTN | BTN |
| JS1093 | BTN | BTN | BTN | BTN | BTN |
| JS1094 | BTN | FN | BTN | BTN | BTN |
| JS1095 | BTN | BTN | BTN | BTN | BTN |
| JS1096 | BTN | BTN | BTN | BTN | BTN |
| JS1097 | BTN | BTN | BTN | BTN | BTN |
| JS1098 | BTN | AUS | FN | FN | FN |
| JS1099 | BTN | BTN | BTN | BTN | BTN |
| JS1100 | BTN | BTN | BTN | BTN | BTN |
| JS1101 | BTN | AUS | AUS | AUS | AUS |
| JS1102 | BTN | BTN | BTN | BTN | BTN |
| JS1103 | BTN | BTN | BTN | BTN | BTN |
| JS1104 | BTN | AUS | FN | FN | FN |
| JS1105 | BTN | BTN | BTN | BTN | BTN |
| JS1106 | BTN | BTN | BTN | BTN | BTN |
| JS1107 | BTN | BTN | BTN | BTN | BTN |
| JS1108 | BTN | SM | FN | FN | FN |
| JS1109 | BTN | BTN | BTN | BTN | BTN |
| JS1110 | BTN | FN | FN | FN | FN |
| JS1111 | BTN | AUS | AUS | AUS | AUS |
| JS1112 | BTN | FN | BTN | BTN | BTN |
| JS1113 | BTN | AUS | AUS | AUS | AUS |
| JS1114 | BTN | SM | SM | SM | SM |
| JS1115 | BTN | PTC | SM | SM | SM |
| JS1116 | BTN | BTN | BTN | BTN | BTN |
| JS1117 | BTN | PTC | SM | SM | SM |
| SH0001 | PTC | PTC | PTC | FTC | PTC |
| SH0005 | PTC | PTC | PTC | PTC | PTC |
| SH0006 | PTC | PTC | SM | SM | SM |
| SH0013 | PTC | PTC | PTC | PTC | PTC |
| SH0017 | PTC | PTC | PTC | FTC | PTC |
| SH0018 | PTC | PTC | PTC | PTC | PTC |
| SH0020 | PTC | PTC | PTC | PTC | PTC |

| | | | | | |
|---|---|---|---|---|---|
| SH0022 | PTC | PTC | PTC | AUS | PTC |
| SH0024 | PTC | PTC | PTC | PTC | PTC |
| SH0028 | PTC | AUS | FN | AUS | AUS |
| SH0030 | PTC | PTC | PTC | PTC | PTC |
| SH0032 | PTC | PTC | PTC | PTC | PTC |
| SH0035 | PTC | FTC | PTC | PTC | PTC |
| SH0036 | PTC | PTC | PTC | SM | PTC |
| SH0037 | PTC | PTC | PTC | PTC | PTC |
| SH0041 | PTC | PTC | PTC | SM | PTC |
| SH0043 | PTC | PTC | PTC | PTC | PTC |
| SH0044 | PTC | PTC | PTC | PTC | PTC |
| SH0045 | PTC | PTC | SM | PTC | PTC |
| SH0046 | PTC | PTC | PTC | PTC | PTC |
| SH0047 | PTC | PTC | SM | PTC | PTC |
| SH0049 | PTC | PTC | PTC | PTC | PTC |
| SH0052 | PTC | PTC | PTC | FTC | PTC |
| SH0053 | PTC | PTC | PTC | PTC | PTC |
| SH0054 | PTC | PTC | SM | PTC | PTC |
| SH0056 | PTC | PTC | PTC | PTC | PTC |
| SH0057 | PTC | AUS | AUS | AUS | AUS |
| SH0059 | PTC | PTC | PTC | PTC | PTC |
| SH0061 | PTC | SM | SM | SM | SM |
| SH0062 | PTC | AUS | AUS | AUS | AUS |
| SH0065 | PTC | PTC | PTC | PTC | PTC |
| SH0072 | PTC | PTC | PTC | PTC | PTC |
| SH0076 | PTC | SM | PTC | PTC | PTC |
| SH0078 | PTC | PTC | PTC | PTC | PTC |
| SH0079 | PTC | PTC | PTC | PTC | PTC |
| SH0080 | PTC | AUS | AUS | AUS | AUS |
| SH0085 | PTC | PTC | PTC | PTC | PTC |
| SH0087 | PTC | PTC | PTC | PTC | PTC |
| SH0089 | PTC | PTC | PTC | PTC | PTC |
| SH0090 | PTC | PTC | PTC | PTC | PTC |
| SH0092 | PTC | PTC | PTC | PTC | PTC |
| SH0093 | PTC | PTC | SM | SM | SM |
| SH0097 | PTC | PTC | PTC | FTC | PTC |
| SH0098 | PTC | PTC | PTC | PTC | PTC |
| SH0099 | PTC | SM | SM | AUS | SM |
| SH0101 | PTC | PTC | PTC | PTC | PTC |
| SH0103 | PTC | SM | SM | SM | SM |
| SH0104 | PTC | PTC | PTC | PTC | PTC |
| SH0105 | PTC | AUS | PTC | PTC | PTC |
| SH0106 | PTC | PTC | PTC | PTC | PTC |
| SH0108 | PTC | FN | SM | SM | SM |

| | | | | | |
|---|---|---|---|---|---|
| SH0109 | PTC | PTC | PTC | PTC | PTC |
| SH0110 | PTC | PTC | FTC | PTC | PTC |
| SH0111 | PTC | PTC | PTC | PTC | PTC |
| SH0113 | PTC | AUS | AUS | AUS | AUS |
| SH0114 | PTC | PTC | PTC | PTC | PTC |
| SH0116 | PTC | SM | PTC | PTC | PTC |
| SH0117 | PTC | PTC | PTC | PTC | PTC |
| SH0120 | PTC | SM | SM | SM | SM |
| SH0123 | PTC | PTC | SM | SM | SM |
| SH0124 | PTC | PTC | PTC | SM | PTC |
| SH0125 | PTC | PTC | PTC | PTC | PTC |
| SH0127 | PTC | PTC | PTC | PTC | PTC |
| SH0128 | PTC | SM | SM | FN | SM |
| SH0132 | PTC | PTC | PTC | PTC | PTC |
| SH0137 | PTC | AUS | FN | AUS | AUS |
| SH0139 | PTC | PTC | PTC | PTC | PTC |
| SH0143 | PTC | PTC | PTC | PTC | PTC |
| SH0144 | PTC | AUS | AUS | AUS | AUS |
| SH0145 | PTC | AUS | FN | AUS | AUS |
| SH0146 | PTC | PTC | PTC | PTC | PTC |
| SH0156 | PTC | PTC | PTC | SM | PTC |
| SH0157 | PTC | PTC | PTC | PTC | PTC |
| SH0159 | PTC | PTC | SM | SM | SM |
| SH0161 | PTC | SM | SM | SM | SM |
| JS0892 | BTN | FN | BTN | BTN | BTN |
| JS0895 | BTN | BTN | AUS | BTN | BTN |
| JS2001 | PTC | FN | SM | SM | SM |
| JS2004 | PTC | FN | SM | SM | SM |
| JS2005 | PTC | SM | PTC | PTC | PTC |
| JS2008 | PTC | AUS | FN | FN | FN |
| JS2010 | PTC | AUS | FN | FN | FN |
| JS2013 | PTC | AUS | SM | SM | SM |
| JS2014 | PTC | SM | PTC | PTC | PTC |
| JS2016 | PTC | AUS | PTC | PTC | PTC |
| JS2018 | PTC | FN | SM | SM | SM |
| JS2019 | PTC | FN | AUS | AUS | AUS |
| JS2022 | PTC | AUS | SM | SM | SM |
| JS2023 | PTC | FN | AUS | AUS | AUS |
| JS2024 | PTC | SM | PTC | PTC | PTC |
| JS2025 | PTC | FN | SM | SM | SM |
| JS2026 | PTC | AUS | SM | SM | SM |
| JS2027 | PTC | SM | PTC | PTC | PTC |
| JS2028 | PTC | SM | PTC | PTC | PTC |
| JS2029 | PTC | SM | PTC | PTC | PTC |
| JS2030 | PTC | AUS | FN | FN | FN |

| | | | | | |
|---|---|---|---|---|---|
| JS2031 | PTC | FN | SM | SM | SM |
| JS2032 | PTC | AUS | SM | SM | SM |
| JS2033 | PTC | FN | AUS | AUS | AUS |
| JS2036 | PTC | AUS | PTC | PTC | PTC |
| JS2037 | PTC | FN | SM | SM | SM |
| JS2039 | PTC | FN | AUS | AUS | AUS |
| JS2040 | PTC | FN | PTC | PTC | PTC |
| JS2041 | PTC | FN | PTC | PTC | PTC |
| JS2042 | PTC | FN | PTC | PTC | PTC |
| JS2043 | PTC | SM | AUS | AUS | AUS |
| JS2044 | PTC | AUS | BTN | BTN | BTN |
| JS2046 | PTC | FN | PTC | PTC | PTC |
| JS2048 | PTC | FN | PTC | PTC | PTC |
| JS2049 | PTC | AUS | BTN | BTN | BTN |
| JS2051 | PTC | FN | PTC | PTC | PTC |
| JS2052 | PTC | SM | AUS | AUS | AUS |
| JS2053 | PTC | SM | PTC | PTC | PTC |
| JS2054 | PTC | SM | PTC | PTC | PTC |
| JS2056 | PTC | SM | PTC | PTC | PTC |
| JS2057 | PTC | AUS | FN | FN | FN |
| JS2058 | PTC | AUS | BTN | BTN | BTN |
| JS2061 | PTC | FN | BTN | BTN | BTN |
| JS2062 | PTC | AUS | SM | SM | SM |
| JS2063 | PTC | PTC | AUS | PTC | PTC |
| JS2065 | PTC | AUS | FN | FN | FN |
| JS2067 | PTC | FN | SM | SM | SM |
| JS2069 | PTC | PTC | FN | PTC | PTC |
| JS2070 | PTC | AUS | FN | FN | FN |
| JS2071 | PTC | BTN | FN | FN | FN |
| JS2074 | PTC | BTN | FN | FN | FN |
| JS2076 | PTC | AUS | FN | FN | FN |
| JS2077 | PTC | FN | AUS | AUS | AUS |
| JS2078 | PTC | SM | FN | FN | FN |
| JS2083 | PTC | SM | AUS | AUS | AUS |
| JS2084 | PTC | SM | AUS | AUS | AUS |
| JS2085 | PTC | AUS | FN | FN | FN |
| JS2086 | PTC | BTN | FN | FN | FN |
| JS2087 | PTC | AUS | FN | FN | FN |
| JS2089 | PTC | BTN | FN | FN | FN |
| JS2091 | PTC | FN | AUS | FN | FN |
| JS2093 | PTC | AUS | SM | SM | SM |
| JS2095 | PTC | FN | BTN | FN | FN |
| JS2096 | PTC | AUS | SM | SM | SM |
| JS2098 | PTC | PTC | SM | PTC | PTC |
| JS2100 | PTC | FN | SM | FN | FN |
| JS2101 | PTC | AUS | SM | SM | SM |

| | | | | | |
|---|---|---|---|---|---|
| JS2102 | PTC | FN | AUS | FN | FN |
| JS2103 | PTC | FN | BTN | FN | FN |
| JS2104 | PTC | FN | AUS | FN | FN |
| JS2105 | PTC | FN | AUS | FN | FN |
| JS2106 | PTC | FN | AUS | AUS | AUS |
| JS2107 | PTC | FN | AUS | FN | FN |
| JS2108 | PTC | FN | AUS | FN | FN |
| JS2110 | PTC | FN | AUS | AUS | AUS |
| JS2112 | PTC | FN | AUS | AUS | AUS |
| JS2114 | PTC | FN | AUS | FN | FN |
| JS2115 | PTC | FN | SM | FN | FN |
| JS2117 | PTC | FN | SM | FN | FN |
| JS2119 | PTC | FN | AUS | AUS | AUS |
| JS2121 | PTC | FN | SM | FN | FN |
| JS2123 | PTC | FN | SM | FN | FN |
| JS2125 | PTC | FN | SM | FN | FN |
| JS2127 | PTC | AUS | FN | AUS | AUS |
| JS2128 | PTC | FN | SM | SM | SM |
| JS2129 | PTC | SM | FN | SM | SM |
| JS2130 | PTC | FN | BTN | FN | FN |
| JS2132 | PTC | FN | AUS | FN | FN |
| JS2133 | PTC | FN | AUS | FN | FN |
| JS2134 | PTC | FN | AUS | FN | FN |
| JS2135 | PTC | AUS | FN | AUS | AUS |
| JS2137 | PTC | SM | FN | SM | SM |
| JS2139 | PTC | AUS | FN | AUS | AUS |
| JS2140 | PTC | FN | AUS | FN | FN |
| JS2141 | PTC | AUS | BTN | AUS | AUS |
| JS2146 | PTC | SM | AUS | SM | SM |
| JS2147 | PTC | PTC | FN | PTC | PTC |
| JS2149 | PTC | SM | FN | SM | SM |
| JS2151 | PTC | SM | AUS | SM | SM |
| JS2154 | PTC | FN | FN | AUS | FN |
| JS2158 | PTC | SM | AUS | SM | SM |
| JS2161 | PTC | FN | FN | AUS | FN |
| JS2162 | PTC | SM | FN | SM | SM |
| JS2164 | PTC | AUS | BTN | AUS | AUS |
| JS2167 | PTC | PTC | SM | PTC | PTC |
| JS2171 | PTC | SM | AUS | SM | SM |
| JS2175 | PTC | SM | AUS | SM | SM |
| JS2180 | PTC | SM | FN | SM | SM |
| JS2181 | PTC | PTC | AUS | PTC | PTC |
| JS2182 | PTC | PTC | FN | PTC | PTC |
| JS2184 | PTC | FN | FN | AUS | FN |
| JS2185 | PTC | SM | FN | SM | SM |
| JS2187 | PTC | PTC | SM | PTC | PTC |

| | | | | | |
|---|---|---|---|---|---|
| JS2188 | PTC | PTC | FN | PTC | PTC |
| JS2191 | PTC | SM | AUS | SM | SM |
| JS2192 | PTC | SM | PTC | SM | SM |
| JS2194 | PTC | SM | PTC | SM | SM |
| JS2195 | PTC | SM | SM | PTC | SM |
| JS2197 | PTC | PTC | AUS | PTC | PTC |
| JS2198 | PTC | SM | SM | PTC | SM |
| JS2199 | PTC | FN | FN | AUS | FN |
| JS2201 | PTC | AUS | BTN | AUS | AUS |
| JS3003 | FTA | FN | BTN | BTN | BTN |
| JS3005 | FTA | US | BTN | BTN | BTN |
| JS3007 | TFND | FN | BTN | BTN | BTN |
| JS3011 | TFND | AUS | BTN | BTN | BTN |
| JS3016 | FTA | AUS | FN | AUS | AUS |
| JS3027 | TFND | FN | FN | AUS | FN |
| JS3029 | FTA | AUS | BTN | AUS | AUS |
| JS3032 | FTA | FN | FN | SM | FN |
| JS3035 | TFND | AUS | SM | AUS | AUS |
| JS3040 | FTA | FN | FN | SM | FN |
| JS3044 | TFND | AUS | FN | AUS | AUS |
| JS3047 | TFND | AUS | BTN | AUS | AUS |
| JS3049 | FTA | PTC | FN | PTC | PTC |
| JS3052 | FTA | AUS | FN | AUS | AUS |
| JS3055 | FTA | AUS | BTN | AUS | AUS |
| JS3062 | TFND | FN | FN | BTN | FN |
| JS3065 | TFND | AUS | SM | AUS | AUS |
| JS3070 | FTA | AUS | AUS | FN | AUS |
| JS2002 | PTC | SM | SM | PTC | SM |
| JS2003 | PTC | AUS | AUS | BTN | AUS |
| JS2006 | PTC | PTC | SM | PTC | PTC |
| JS2007 | PTC | PTC | PTC | SM | PTC |
| JS2009 | PTC | PTC | PTC | FN | PTC |
| JS2011 | PTC | PTC | PTC | FN | PTC |
| JS2012 | PTC | PTC | PTC | FN | PTC |
| JS2015 | PTC | SM | SM | AUS | SM |
| JS2017 | PTC | PTC | PTC | FN | PTC |
| JS2020 | PTC | PTC | PTC | AUS | PTC |
| JS2021 | PTC | AUS | AUS | BTN | AUS |
| JS2034 | PTC | AUS | AUS | BTN | AUS |
| JS2035 | PTC | SM | SM | FN | SM |
| JS2038 | PTC | FN | FN | AUS | FN |
| JS2045 | PTC | SM | SM | PTC | SM |
| JS2047 | PTC | BTN | AUS | BTN | BTN |
| JS2050 | PTC | PTC | PTC | AUS | PTC |
| JS2055 | PTC | PTC | PTC | AUS | PTC |
| JS2059 | PTC | PTC | PTC | FN | PTC |

| | | | | | |
|---|---|---|---|---|---|
| JS2060 | PTC | BTN | FN | BTN | BTN |
| JS2064 | PTC | AUS | AUS | FN | AUS |
| JS2066 | PTC | AUS | AUS | FN | AUS |
| JS2068 | PTC | SM | SM | AUS | SM |
| JS2072 | PTC | SM | PTC | SM | SM |
| JS2073 | PTC | FN | FN | BTN | FN |
| JS2079 | PTC | PTC | PTC | FN | PTC |
| JS2080 | PTC | FN | FN | SM | FN |
| JS2081 | PTC | AUS | AUS | BTN | AUS |
| JS2082 | PTC | AUS | SM | SM | SM |
| JS2088 | PTC | FN | FN | AUS | FN |
| JS2097 | PTC | AUS | AUS | FN | AUS |
| JS2109 | PTC | FN | FN | SM | FN |
| JS2116 | PTC | FN | FN | BTN | FN |
| JS2120 | PTC | FN | FN | BTN | FN |
| JS2122 | PTC | AUS | AUS | BTN | AUS |
| JS2124 | PTC | FN | FN | BTN | FN |
| JS2126 | PTC | FN | FN | AUS | FN |
| JS2138 | PTC | FN | FN | AUS | FN |
| JS2142 | PTC | FN | FN | SM | FN |
| JS2144 | PTC | FN | FN | SM | FN |
| JS2145 | PTC | PTC | PTC | SM | PTC |
| JS2148 | PTC | PTC | PTC | SM | PTC |
| JS2152 | PTC | SM | PTC | SM | SM |
| JS2159 | PTC | FN | FN | AUS | FN |
| JS2186 | PTC | FN | FN | BTN | FN |
| JS2189 | PTC | AUS | AUS | FN | AUS |
| JS2190 | PTC | FN | AUS | FN | FN |
| JS2193 | PTC | FN | BTN | FN | FN |
| JS2196 | PTC | FN | SM | FN | FN |
| JS2200 | PTC | SM | PTC | SM | SM |
| JS3001 | FTA | AUS | AUS | FN | AUS |
| JS3002 | FTA | BTN | AUS | BTN | BTN |
| JS3004 | TFND | BTN | FN | BTN | BTN |
| JS3006 | FTA | FN | SM | SM | SM |
| JS3008 | TFND | BTN | FN | BTN | BTN |
| JS3009 | FTA | AUS | SM | SM | SM |
| JS3010 | TFND | SM | PTC | SM | SM |
| JS3012 | TFND | BTN | AUS | BTN | BTN |
| JS3013 | TFND | BTN | AUS | BTN | BTN |
| JS3014 | FTA | FN | SM | FN | FN |
| JS3015 | FTA | SM | AUS | SM | SM |
| JS3017 | FTA | BTN | BTN | AUS | BTN |
| JS3018 | TFND | AUS | AUS | FN | AUS |
| JS3019 | FTA | BTN | BTN | AUS | BTN |
| JS3020 | TFND | AUS | AUS | SM | AUS |

| | | | | | |
|---|---|---|---|---|---|
| JS3021 | TFND | AUS | AUS | SM | AUS |
| JS3022 | FTA | AUS | AUS | BTN | AUS |
| JS3023 | TFND | FN | BTN | FN | FN |
| JS3024 | TFND | FN | BTN | FN | FN |
| JS3025 | TFND | AUS | SM | SM | SM |
| JS3026 | FTA | AUS | AUS | FN | AUS |
| JS3028 | FTA | AUS | BTN | AUS | AUS |
| JS3030 | FTA | FN | AUS | FN | FN |
| JS3031 | FTA | AUS | BTN | AUS | AUS |
| JS3033 | TFND | AUS | FN | AUS | AUS |
| JS3034 | FTA | AUS | BTN | AUS | AUS |
| JS3036 | TFND | BTN | BTN | FN | BTN |
| JS3037 | FTA | FN | AUS | FN | FN |
| JS3038 | TFND | FN | AUS | FN | FN |
| JS3039 | TFND | BTN | FN | FN | FN |
| JS3041 | FTA | AUS | FN | AUS | AUS |
| JS3042 | FTA | AUS | BTN | AUS | AUS |
| JS3043 | TFND | SM | PTC | SM | SM |
| JS3045 | FTA | SM | PTC | SM | SM |
| JS3046 | TFND | BTN | FN | FN | FN |
| JS3048 | FTA | AUS | FN | FN | FN |
| JS3050 | FTA | PTC | PTC | FN | PTC |
| JS3051 | FTA | AUS | FN | FN | FN |
| JS3053 | TFND | AUS | SM | AUS | AUS |
| JS3054 | FTA | BTN | AUS | AUS | AUS |
| JS3056 | FTA | SM | FN | FN | FN |
| JS3057 | TFND | AUS | FN | FN | FN |
| JS3058 | TFND | FN | AUS | AUS | AUS |
| JS3059 | FTA | AUS | FN | FN | FN |
| JS3060 | TFND | AUS | FN | FN | FN |
| JS3061 | FTA | BTN | AUS | AUS | AUS |
| JS3063 | TFND | AUS | SM | AUS | AUS |
| JS3064 | FTA | AUS | FN | FN | FN |
| JS3066 | FTA | PTC | PTC | FN | PTC |
| JS3067 | TFND | AUS | SM | SM | SM |
| JS3068 | TFND | AUS | SM | SM | SM |
| JS3069 | TFND | PTC | PTC | AUS | PTC |
| JS3071 | FTA | PTC | PTC | SM | PTC |
| JS3072 | FTA | AUS | BTN | AUS | AUS |
| JS3073 | FTA | BTN | BTN | AUS | BTN |
| JS3074 | TFND | BTN | BTN | AUS | BTN |
| JS3075 | FTA | AUS | FN | AUS | AUS |
| JS3076 | TFND | AUS | SM | SM | SM |
| JS3077 | FTA | FN | AUS | AUS | AUS |
| JS3078 | TFND | BTN | BTN | AUS | BTN |
| JS6812 | PTC | PTC | PTC | PTC | PTC |

| | | | | | |
|---|---|---|---|---|---|
| JS7400 | PTC | PTC | PTC | PTC | PTC |
| JS7284 | PTC | PTC | PTC | PTC | PTC |
| JS7372 | PTC | PTC | PTC | SM | PTC |
| JS7160 | PTC | PTC | PTC | AUS | PTC |
| JS6731 | PTC | PTC | PTC | PTC | PTC |
| JS7063 | PTC | PTC | PTC | PTC | PTC |
| JS7031 | PTC | PTC | PTC | PTC | PTC |
| JS6885 | PTC | PTC | PTC | PTC | PTC |
| JS7107 | PTC | PTC | SM | PTC | PTC |
| JS6845 | FTC | PTC | PTC | FN | PTC |
| JS7144 | PTC | PTC | PTC | PTC | PTC |
| JS7175 | PTC | PTC | PTC | SM | PTC |
| JS7335 | PTC | PTC | PTC | PTC | PTC |
| JS7222 | PTC | PTC | PTC | PTC | PTC |
| JS6802 | PTC | PTC | PTC | PTC | PTC |
| JS7055 | PTC | PTC | PTC | PTC | PTC |
| JS6882 | PTC | PTC | PTC | PTC | PTC |
| JS7177 | TFND | PTC | PTC | PTC | PTC |
| JS7086 | PTC | PTC | PTC | PTC | PTC |
| JS7161 | PTC | PTC | PTC | SM | PTC |
| JS6862 | PTC | PTC | SM | PTC | PTC |
| JS7361 | PTC | PTC | PTC | PTC | PTC |
| JS7038 | PTC | PTC | PTC | PTC | PTC |
| JS7098 | PTC | PTC | PTC | PTC | PTC |
| JS6808 | PTC | PTC | PTC | PTC | PTC |
| JS7300 | PTC | PTC | PTC | PTC | PTC |
| JS6732 | PTC | PTC | PTC | PTC | PTC |
| JS7428 | PTC | PTC | PTC | PTC | PTC |
| JS7020 | PTC | PTC | PTC | PTC | PTC |
| JS7336 | PTC | PTC | PTC | PTC | PTC |
| JS7047 | PTC | PTC | PTC | SM | PTC |
| JS7075 | PTC | PTC | PTC | PTC | PTC |
| JS6720 | PTC | PTC | PTC | PTC | PTC |
| JS7465 | PTC | PTC | PTC | PTC | PTC |
| JS7234 | PTC | PTC | PTC | PTC | PTC |
| JS7029 | PTC | PTC | PTC | PTC | PTC |
| JS6877 | PTC | PTC | PTC | PTC | PTC |
| JS7329 | PTC | PTC | PTC | PTC | PTC |
| JS7045 | PTC | PTC | SM | PTC | PTC |
| JS7441 | PTC | PTC | PTC | PTC | PTC |
| JS6841 | PTC | PTC | PTC | PTC | PTC |
| JS6846 | PTC | PTC | PTC | PTC | PTC |
| JS6807 | PTC | PTC | SM | PTC | PTC |
| JS6866 | PTC | PTC | PTC | PTC | PTC |
| JS7314 | PTC | PTC | PTC | PTC | PTC |
| JS7347 | PTC | PTC | PTC | PTC | PTC |

| | | | | | |
|---|---|---|---|---|---|
| JS7339 | PTC | PTC | PTC | PTC | PTC |
| JS7181 | PTC | PTC | PTC | PTC | PTC |
| JS7440 | PTC | SM | PTC | PTC | PTC |
| JS7455 | PTC | PTC | PTC | PTC | PTC |
| JS7230 | TFND | SM | SM | SM | SM |
| JS7288 | PTC | SM | SM | PTC | SM |
| JS7256 | PTC | SM | SM | PTC | SM |
| JS6887 | PTC | SM | SM | PTC | SM |
| JS7186 | PTC | SM | SM | SM | SM |
| JS7199 | TFND | SM | SM | PTC | SM |
| JS6848 | TFND | SM | SM | SM | SM |
| JS7194 | PTC | SM | SM | SM | SM |
| JS7423 | PTC | SM | SM | PTC | SM |
| JS7424 | PTC | SM | SM | SM | SM |
| JS7179 | PTC | SM | SM | SM | SM |
| JS6895 | PTC | SM | SM | SM | SM |
| JS7324 | PTC | SM | SM | SM | SM |
| JS6803 | PTC | SM | SM | SM | SM |
| JS6891 | FTA | SM | SM | PTC | SM |
| JS7419 | PTC | SM | SM | SM | SM |
| JS7191 | PTC | SM | SM | SM | SM |
| JS7285 | PTC | SM | SM | SM | SM |
| JS7262 | PTC | SM | SM | SM | SM |
| JS7427 | PTC | SM | SM | SM | SM |
| JS7074 | PTC | SM | SM | PTC | SM |
| JS7219 | PTC | SM | SM | SM | SM |
| JS6729 | PTC | SM | SM | SM | SM |
| JS7124 | PTC | SM | SM | SM | SM |
| JS7286 | PTC | SM | SM | SM | SM |
| JS7126 | PTC | SM | SM | PTC | SM |
| JS6797 | PTC | SM | SM | SM | SM |
| JS7121 | PTC | SM | SM | SM | SM |
| JS6734 | PTC | SM | SM | PTC | SM |
| JS7238 | TFND | FN | FN | PTC | FN |
| JS7235 | FTA | FN | FN | SM | FN |
| JS6850 | PTC | FN | FN | PTC | FN |
| JS7145 | PTC | FN | FN | PTC | FN |
| JS7218 | PTC | FN | FN | FN | FN |
| JS7198 | PTC | FN | FN | PTC | FN |
| JS7357 | PTC | FN | FN | FN | FN |
| JS7174 | PTC | FN | FN | PTC | FN |
| JS7272 | TFND | AUS | AUS | AUS | AUS |
| JS7127 | PTC | AUS | AUS | AUS | AUS |
| JS6886 | FTA | AUS | AUS | AUS | AUS |
| JS7379 | PTC | AUS | AUS | AUS | AUS |
| JS7275 | FTA | AUS | BTN | AUS | AUS |

| | | | | | |
|---|---|---|---|---|---|
| JS6832 | FTA | AUS | BTN | AUS | AUS |
| JS7257 | PTC | AUS | AUS | AUS | AUS |
| JS6723 | TFND | AUS | AUS | AUS | AUS |
| JS7343 | PTC | AUS | AUS | AUS | AUS |
| JS7382 | PTC | AUS | AUS | AUS | AUS |
| JS7334 | PTC | AUS | AUS | AUS | AUS |
| JS7330 | PTC | AUS | AUS | AUS | AUS |
| JS6779 | PTC | AUS | AUS | AUS | AUS |
| JS6706 | PTC | AUS | AUS | AUS | AUS |
| JS7340 | PTC | AUS | AUS | AUS | AUS |
| JS7326 | PTC | AUS | AUS | AUS | AUS |
| JS7103 | TFND | AUS | BTN | AUS | AUS |
| JS7140 | FTA | AUS | BTN | AUS | AUS |
| JS7387 | PTC | AUS | AUS | AUS | AUS |
| JS7147 | TFND | BTN | BTN | BTN | BTN |
| JS7362 | TFND | BTN | BTN | BTN | BTN |
| JS7466 | PTC | AUS | BTN | BTN | BTN |
| JS7290 | PTC | BTN | BTN | BTN | BTN |
| JS7071 | TFND | BTN | AUS | BTN | BTN |
| JS7012 | PTC | BTN | BTN | BTN | BTN |
| JS6894 | TFND | BTN | BTN | BTN | BTN |
| JS7282 | FTA | BTN | BTN | BTN | BTN |
| JS7239 | FTA | BTN | BTN | AUS | BTN |
| JS7443 | PTC | BTN | BTN | BTN | BTN |
| JS7125 | TFND | BTN | BTN | BTN | BTN |
| JS7429 | PTC | BTN | BTN | BTN | BTN |
| JS7446 | TFND | BTN | BTN | BTN | BTN |
| JS7193 | TFND | BTN | BTN | BTN | BTN |
| JS6730 | FTA | BTN | BTN | BTN | BTN |

BTN, Benign thyroid nodules
AUS, Atypia of Undetermined Significance
FN, Follicular Neoplasm
SM, Suspicious for Malignancy
FTC, follicular thyroid carcinoma.
PTC, papillary thyroid carcinoma.

**Supplementary Table S6.** Independent Bethesda cytological review results of FNAB specimens in the model training cohort without histopathologically confirmed diagnosis by three senior thyroid pathologists

| Case ID | Final Histology | Bethesda Cytological Categories | | | Final Bethesda Categories |
|---|---|---|---|---|---|
| | | Dr. Wu | Dr. Zhu | Dr. Wan | |
| JS0001 | No surgery | BTN | AUS | BTN | BTN |
| JS0002 | No surgery | FN | AUS | AUS | AUS |
| JS0003 | No surgery | AUS | FN | FN | FN |
| JS0004 | No surgery | FN | AUS | FN | FN |
| JS0005 | No surgery | BTN | AUS | BTN | BTN |

| | | | | | |
|---|---|---|---|---|---|
| JS0006 | No surgery | FN | BTN | BTN | BTN |
| JS0007 | No surgery | FN | FN | AUS | FN |
| JS0008 | No surgery | BTN | BTN | AUS | BTN |
| JS0009 | No surgery | AUS | BTN | BTN | BTN |
| JS0010 | No surgery | BTN | FN | FN | FN |
| JS0011 | No surgery | BTN | FN | BTN | BTN |
| JS0012 | No surgery | BTN | BTN | BTN | BTN |
| JS0013 | No surgery | AUS | BTN | BTN | BTN |
| JS0014 | No surgery | AUS | BTN | BTN | BTN |
| JS0015 | No surgery | FN | AUS | AUS | AUS |
| JS0016 | No surgery | AUS | FN | AUS | AUS |
| JS0017 | No surgery | BTN | BTN | AUS | BTN |
| JS0018 | No surgery | AUS | FN | FN | FN |
| JS0019 | No surgery | FN | AUS | AUS | AUS |
| JS0020 | No surgery | AUS | BTN | BTN | BTN |
| JS0021 | No surgery | BTN | AUS | BTN | BTN |
| JS0022 | No surgery | FN | BTN | BTN | BTN |
| JS0023 | No surgery | AUS | BTN | BTN | BTN |
| JS0024 | No surgery | BTN | BTN | AUS | BTN |
| JS0025 | No surgery | BTN | BTN | FN | BTN |
| JS0026 | No surgery | AUS | AUS | FN | AUS |
| JS0027 | No surgery | BTN | FN | BTN | BTN |
| JS0028 | No surgery | AUS | BTN | BTN | BTN |
| JS0029 | No surgery | AUS | BTN | BTN | BTN |
| JS0030 | No surgery | FN | BTN | BTN | BTN |
| JS0031 | No surgery | BTN | BTN | BTN | BTN |
| JS0032 | No surgery | BTN | AUS | BTN | BTN |
| JS0033 | No surgery | BTN | AUS | BTN | BTN |
| JS0034 | No surgery | FN | AUS | FN | FN |
| JS0035 | No surgery | SM | PTC | SM | SM |
| JS0037 | No surgery | BTN | AUS | BTN | BTN |
| JS0038 | No surgery | FN | BTN | FN | FN |
| JS0040 | No surgery | BTN | BTN | AUS | BTN |
| JS0041 | No surgery | BTN | BTN | AUS | BTN |
| JS0042 | No surgery | AUS | BTN | BTN | BTN |
| JS0043 | No surgery | AUS | BTN | BTN | BTN |
| JS0044 | No surgery | BTN | BTN | BTN | BTN |
| JS0045 | No surgery | AUS | BTN | BTN | BTN |
| JS0046 | No surgery | BTN | AUS | BTN | BTN |
| JS0047 | No surgery | AUS | BTN | BTN | BTN |
| JS0048 | No surgery | AUS | AUS | FN | AUS |
| JS0049 | No surgery | AUS | BTN | BTN | BTN |
| JS0050 | No surgery | FN | BTN | BTN | BTN |
| JS0051 | No surgery | BTN | BTN | AUS | BTN |
| JS0052 | No surgery | BTN | BTN | AUS | BTN |
| JS0053 | No surgery | BTN | BTN | AUS | BTN |

| | | | | | |
|---|---|---|---|---|---|
| JS0054 | No surgery | AUS | FN | FN | FN |
| JS0055 | No surgery | FN | AUS | FN | FN |
| JS0056 | No surgery | BTN | AUS | BTN | BTN |
| JS0059 | No surgery | BTN | BTN | AUS | BTN |
| JS0060 | No surgery | BTN | AUS | BTN | BTN |
| JS0061 | No surgery | AUS | BTN | BTN | BTN |
| JS0062 | No surgery | AUS | BTN | BTN | BTN |
| JS0099 | No surgery | AUS | BTN | BTN | BTN |
| JS0121 | No surgery | AUS | BTN | BTN | BTN |
| JS0194 | No surgery | AUS | AUS | FN | AUS |
| JS0251 | No surgery | AUS | BTN | BTN | BTN |
| JS0252 | No surgery | FN | SM | SM | SM |
| JS0253 | No surgery | BTN | AUS | BTN | BTN |
| JS0284 | No surgery | BTN | AUS | BTN | BTN |
| JS0285 | No surgery | BTN | BTN | AUS | BTN |
| JS0286 | No surgery | BTN | BTN | AUS | BTN |
| JS0287 | No surgery | BTN | BTN | AUS | BTN |
| JS0288 | No surgery | BTN | AUS | BTN | BTN |
| JS0289 | No surgery | BTN | AUS | BTN | BTN |
| JS0290 | No surgery | BTN | AUS | BTN | BTN |
| JS0292 | No surgery | AUS | FN | FN | FN |
| JS0294 | No surgery | AUS | BTN | BTN | BTN |
| JS0295 | No surgery | AUS | BTN | BTN | BTN |
| JS0302 | No surgery | FN | AUS | FN | FN |
| JS0303 | No surgery | BTN | AUS | BTN | BTN |
| JS0343 | No surgery | BTN | BTN | AUS | BTN |
| JS0344 | No surgery | AUS | BTN | BTN | BTN |
| JS0348 | No surgery | BTN | BTN | AUS | BTN |
| JS0350 | No surgery | BTN | BTN | AUS | BTN |

BTN, Benign thyroid nodules
AUS, Atypia of Undetermined Significance
FN, Follicular Neoplasm
SM, Suspicious for Malignancy
FTC, follicular thyroid carcinoma.
PTC, papillary thyroid carcinoma.

**Supplementary Table S7.** Independent ACR TI-RADS review results of thyroid ultrasonographic imaging in the subjects with histopathologically confirmed diagnosis by three senior ultrasound radiologists

| Case ID | Final Histology | ACR TI-RADS Categories | | | Final ACR TI-RADS |
|---|---|---|---|---|---|
| | | Dr. Lou | Dr. Jin | Dr. Lyu | |
| AH0023 | PTC | 4 | 4 | 4 | 4 |
| AH0057 | BTN | 4 | 4 | 4 | 4 |
| AH0094 | BTN | 4 | 4 | 4 | 4 |
| AH0106 | PTC | 4 | 5 | 5 | 5 |
| AH0107 | PTC | 4 | 4 | 4 | 4 |
| AH0111 | PTC | 4 | 4 | 4 | 4 |
| AH0115 | PTC | 4 | 4 | 4 | 4 |
| AH0127 | PTC | 4 | 4 | 4 | 4 |
| AH0134 | PTC | 5 | 5 | 5 | 5 |
| AH0139 | PTC | 3 | 4 | 3 | 3 |
| AH0156 | PTC | 5 | 5 | 5 | 5 |
| AH0164 | PTC | 4 | 4 | 4 | 4 |
| AH0170 | PTC | 5 | 5 | 5 | 5 |
| AH0174 | PTC | 5 | 5 | 5 | 5 |
| AH0191 | PTC | 4 | 4 | 4 | 4 |
| AH0197 | PTC | 4 | 4 | 4 | 4 |
| AH0220 | PTC | 5 | 5 | 5 | 5 |
| AH0222 | PTC | 4 | 4 | 4 | 4 |
| AH0257 | PTC | 5 | 5 | 5 | 5 |
| AH0259 | PTC | 3 | 3 | 3 | 3 |
| AH0261 | PTC | 5 | 5 | 5 | 5 |
| AH0262 | PTC | 4 | 5 | 5 | 5 |
| AH0264 | PTC | 5 | 5 | 5 | 5 |
| AH0267 | PTC | 5 | 5 | 5 | 5 |
| AH0272 | PTC | 5 | 5 | 5 | 5 |
| AH0275 | PTC | 5 | 5 | 5 | 5 |
| AH0279 | PTC | 5 | 5 | 5 | 5 |
| AH0290 | PTC | 5 | 5 | 5 | 5 |
| AH0291 | PTC | 5 | 5 | 5 | 5 |
| AH0300 | PTC | 5 | 5 | 5 | 5 |
| AH0312 | PTC | 5 | 5 | 5 | 5 |
| AH0313 | PTC | 4 | 4 | 4 | 4 |
| AH0322 | PTC | 4 | 4 | 4 | 4 |
| AH0325 | PTC | 5 | 5 | 5 | 5 |
| AH0327 | PTC | 4 | 3 | 4 | 4 |
| AH0331 | PTC | 5 | 5 | 5 | 5 |
| AH0334 | PTC | 5 | 5 | 5 | 5 |
| AH0335 | PTC | 5 | 4 | 5 | 5 |
| AH0336 | PTC | 5 | 5 | 5 | 5 |
| AH0337 | PTC | 4 | 4 | 4 | 4 |

| | | | | | |
|---|---|---|---|---|---|
| AH0340 | PTC | 5 | 5 | 5 | 5 |
| AH0342 | PTC | 4 | 4 | 4 | 4 |
| AH0358 | PTC | 5 | 5 | 5 | 5 |
| AH0391 | BTN | 3 | 3 | 3 | 3 |
| JS0696 | BTN | 5 | 5 | 5 | 5 |
| JS0699 | BTN | 3 | 2 | 2 | 2 |
| JS0706 | BTN | 3 | 4 | 4 | 4 |
| JS0718 | PTC | 4 | 5 | 4 | 4 |
| JS0720 | PTC | 4 | 4 | 4 | 4 |
| JS0723 | PTC | 5 | 5 | 5 | 5 |
| JS0731 | PTC | 4 | 4 | 4 | 4 |
| JS0737 | PTC | 4 | 4 | 4 | 4 |
| JS0746 | PTC | 4 | 4 | 4 | 4 |
| JS0749 | PTC | 4 | 4 | 4 | 4 |
| JS0750 | PTC | 5 | 4 | 4 | 4 |
| JS0751 | PTC | 4 | 4 | 4 | 4 |
| JS0752 | PTC | 4 | 5 | 4 | 4 |
| JS0758 | BTN | 4 | 4 | 4 | 4 |
| JS0760 | BTN | 4 | 4 | 4 | 4 |
| JS0763 | PTC | 4 | 4 | 4 | 4 |
| JS0767 | BTN | 4 | 3 | 3 | 3 |
| JS0769 | PTC | 5 | 4 | 4 | 4 |
| JS0770 | BTN | 4 | 3 | 4 | 4 |
| JS0771 | PTC | 3 | 3 | 4 | 3 |
| JS0786 | PTC | 5 | 5 | 5 | 5 |
| JS0787 | PTC | 4 | 4 | 4 | 4 |
| JS0788 | PTC | 5 | 5 | 5 | 5 |
| JS0789 | PTC | 5 | 5 | 5 | 5 |
| JS0790 | PTC | 5 | 5 | 5 | 5 |
| JS0791 | PTC | 5 | 4 | 5 | 5 |
| JS0792 | PTC | 5 | 4 | 5 | 5 |
| JS0793 | PTC | 4 | 3 | 4 | 4 |
| JS0794 | FTC | 4 | 4 | 4 | 4 |
| JS0795 | FTC | 4 | 3 | 3 | 3 |
| JS0796 | PTC | 4 | 3 | 3 | 3 |
| JS0797 | PTC | 5 | 5 | 5 | 5 |
| JS0798 | PTC | 4 | 4 | 3 | 4 |
| JS0799 | PTC | 5 | 5 | 5 | 5 |
| JS0800 | PTC | 5 | 5 | 5 | 5 |
| JS0801 | PTC | 4 | 4 | 4 | 4 |
| JS0802 | PTC | 5 | 5 | 5 | 5 |
| JS0803 | PTC | 4 | 5 | 5 | 5 |
| JS0804 | PTC | 5 | 5 | 5 | 5 |
| JS0805 | PTC | 5 | 5 | 5 | 5 |
| JS0806 | PTC | 4 | 5 | 4 | 4 |
| JS0807 | PTC | 4 | 4 | 4 | 4 |

| | | | | | |
|---|---|---|---|---|---|
| JS0808 | PTC | 4 | 4 | 4 | 4 |
| JS0809 | PTC | 5 | 5 | 5 | 5 |
| JS0810 | PTC | 5 | 4 | 4 | 4 |
| JS0811 | PTC | 4 | 4 | 4 | 4 |
| JS0812 | PTC | 5 | 5 | 5 | 5 |
| JS0813 | PTC | 5 | 5 | 5 | 5 |
| JS0814 | PTC | 5 | 5 | 5 | 5 |
| JS0825 | PTC | 3 | 3 | 3 | 3 |
| JS0828 | BTN | 3 | 3 | 3 | 3 |
| JS0844 | PTC | 4 | 4 | 4 | 4 |
| JS0846 | PTC | 3 | 3 | 4 | 3 |
| JS0847 | PTC | 5 | 5 | 5 | 5 |
| JS0848 | PTC | 4 | 4 | 4 | 4 |
| JS0851 | PTC | 5 | 5 | 5 | 5 |
| JS0852 | PTC | 5 | 5 | 5 | 5 |
| JS0857 | PTC | 3 | 3 | 2 | 3 |
| JS0858 | PTC | 5 | 4 | 4 | 4 |
| JS0859 | PTC | 4 | 4 | 4 | 4 |
| JS0860 | PTC | 4 | 4 | 4 | 4 |
| JS0861 | PTC | 4 | 4 | 4 | 4 |
| JS0862 | PTC | 5 | 4 | 4 | 4 |
| JS0863 | PTC | 4 | 4 | 4 | 4 |
| JS0865 | PTC | 5 | 5 | 5 | 5 |
| JS0867 | PTC | 4 | 4 | 4 | 4 |
| JS0868 | PTC | 5 | 4 | 4 | 4 |
| JS0869 | PTC | 5 | 5 | 5 | 5 |
| JS0870 | PTC | 5 | 5 | 5 | 5 |
| JS0871 | PTC | 4 | 4 | 4 | 4 |
| JS0872 | PTC | 5 | 5 | 5 | 5 |
| JS0873 | PTC | 4 | 4 | 4 | 4 |
| JS0874 | PTC | 5 | 4 | 4 | 4 |
| JS0875 | PTC | 5 | 4 | 4 | 4 |
| JS0876 | PTC | 4 | 4 | 4 | 4 |
| JS0877 | PTC | 5 | 5 | 5 | 5 |
| JS0878 | PTC | 5 | 4 | 5 | 5 |
| JS0882 | PTC | 4 | 4 | 5 | 4 |
| JS0885 | PTC | 5 | 5 | 4 | 5 |
| JS0887 | PTC | 5 | 5 | 5 | 5 |
| JS0888 | PTC | 5 | 5 | 5 | 5 |
| JS0890 | PTC | 5 | 5 | 5 | 5 |
| JS0892 | BTN | 4 | 4 | 5 | 4 |
| JS0895 | BTN | 3 | 3 | 4 | 3 |
| JS0991 | BTN | 3 | 3 | 4 | 3 |
| JS0992 | BTN | 4 | 3 | 4 | 4 |
| JS0993 | BTN | 4 | 4 | 4 | 4 |
| JS0994 | BTN | 4 | 5 | 4 | 4 |

| | | | | | |
|---|---|---|---|---|---|
| JS0995 | BTN | 3 | 4 | 3 | 3 |
| JS0996 | BTN | 3 | 4 | 4 | 4 |
| JS0997 | BTN | 3 | 3 | 3 | 3 |
| JS0998 | BTN | 3 | 3 | 3 | 3 |
| JS0999 | BTN | 3 | 3 | 4 | 3 |
| JS1000 | BTN | 4 | 4 | 4 | 4 |
| JS1001 | BTN | 3 | 3 | 3 | 3 |
| JS1002 | BTN | 3 | 3 | 4 | 3 |
| JS1003 | BTN | 3 | 3 | 3 | 3 |
| JS1004 | BTN | 3 | 3 | 3 | 3 |
| JS1005 | BTN | 3 | 3 | 3 | 3 |
| JS1006 | BTN | 3 | 3 | 3 | 3 |
| JS1007 | BTN | 4 | 3 | 3 | 3 |
| JS1008 | BTN | 4 | 3 | 4 | 4 |
| JS1009 | BTN | 3 | 4 | 4 | 4 |
| JS1010 | BTN | 2 | 3 | 3 | 3 |
| JS1011 | BTN | 3 | 3 | 4 | 3 |
| JS1012 | BTN | 4 | 4 | 4 | 4 |
| JS1013 | BTN | 3 | 4 | 4 | 4 |
| JS1014 | BTN | 3 | 3 | 4 | 3 |
| JS1015 | BTN | 3 | 3 | 3 | 3 |
| JS1016 | BTN | 3 | 2 | 3 | 3 |
| JS1017 | BTN | 4 | 4 | 4 | 4 |
| JS1018 | BTN | 3 | 2 | 3 | 3 |
| JS1020 | BTN | 3 | 3 | 4 | 3 |
| JS1021 | BTN | 3 | 3 | 3 | 3 |
| JS1022 | BTN | 2 | 1 | 2 | 2 |
| JS1023 | BTN | 3 | 4 | 4 | 4 |
| JS1024 | BTN | 4 | 5 | 5 | 5 |
| JS1025 | BTN | 4 | 4 | 4 | 4 |
| JS1026 | BTN | 4 | 3 | 3 | 3 |
| JS1027 | BTN | 4 | 5 | 5 | 5 |
| JS1028 | BTN | 3 | 3 | 3 | 3 |
| JS1029 | BTN | 3 | 3 | 3 | 3 |
| JS1030 | BTN | 4 | 5 | 5 | 5 |
| JS1031 | BTN | 3 | 3 | 3 | 3 |
| JS1032 | BTN | 3 | 3 | 4 | 3 |
| JS1033 | BTN | 3 | 4 | 4 | 4 |
| JS1034 | BTN | 3 | 3 | 3 | 3 |
| JS1035 | BTN | 4 | 4 | 4 | 4 |
| JS1036 | BTN | 3 | 3 | 3 | 3 |
| JS1037 | BTN | 3 | 3 | 4 | 3 |
| JS1038 | BTN | 1 | 3 | 3 | 3 |
| JS1039 | BTN | 4 | 4 | 4 | 4 |
| JS1040 | BTN | 4 | 4 | 3 | 4 |
| JS1041 | BTN | 4 | 3 | 4 | 4 |

| | | | | | |
|---|---|---|---|---|---|
| JS1042 | BTN | 3 | 3 | 4 | 3 |
| JS1043 | BTN | 3 | 3 | 3 | 3 |
| JS1044 | BTN | 3 | 3 | 3 | 3 |
| JS1045 | BTN | 3 | 3 | 3 | 3 |
| JS1046 | BTN | 5 | 4 | 5 | 5 |
| JS1047 | BTN | 4 | 4 | 4 | 4 |
| JS1048 | BTN | 4 | 4 | 4 | 4 |
| JS1049 | BTN | 3 | 3 | 3 | 3 |
| JS1050 | BTN | 4 | 4 | 4 | 4 |
| JS1051 | BTN | 3 | 3 | 3 | 3 |
| JS1052 | BTN | 4 | 4 | 4 | 4 |
| JS1053 | BTN | 4 | 4 | 4 | 4 |
| JS1054 | BTN | 3 | 3 | 3 | 3 |
| JS1055 | BTN | 4 | 5 | 5 | 5 |
| JS1056 | BTN | 3 | 3 | 3 | 3 |
| JS1057 | BTN | 3 | 4 | 4 | 4 |
| JS1058 | BTN | 3 | 3 | 3 | 3 |
| JS1059 | BTN | 4 | 4 | 4 | 4 |
| JS1060 | BTN | 4 | 4 | 4 | 4 |
| JS1061 | BTN | 3 | 3 | 3 | 3 |
| JS1062 | BTN | 4 | 4 | 4 | 4 |
| JS1063 | BTN | 4 | 4 | 4 | 4 |
| JS1064 | BTN | 4 | 4 | 4 | 4 |
| JS1065 | BTN | 1 | 3 | 3 | 3 |
| JS1066 | BTN | 4 | 4 | 4 | 4 |
| JS1067 | BTN | 3 | 3 | 3 | 3 |
| JS1068 | BTN | 3 | 3 | 3 | 3 |
| JS1069 | BTN | 3 | 3 | 3 | 3 |
| JS1070 | BTN | 4 | 3 | 4 | 4 |
| JS1071 | BTN | 3 | 3 | 3 | 3 |
| JS1072 | BTN | 4 | 4 | 4 | 4 |
| JS1073 | BTN | 2 | 3 | 2 | 2 |
| JS1074 | BTN | 4 | 4 | 4 | 4 |
| JS1075 | BTN | 4 | 4 | 4 | 4 |
| JS1076 | BTN | 3 | 2 | 3 | 3 |
| JS1077 | BTN | 4 | 4 | 4 | 4 |
| JS1078 | BTN | 3 | 3 | 3 | 3 |
| JS1079 | BTN | 3 | 4 | 4 | 4 |
| JS1080 | BTN | 3 | 3 | 3 | 3 |
| JS1081 | BTN | 3 | 3 | 3 | 3 |
| JS1082 | BTN | 2 | 3 | 3 | 3 |
| JS1083 | BTN | 4 | 4 | 3 | 4 |
| JS1084 | BTN | 3 | 3 | 3 | 3 |
| JS1085 | BTN | 4 | 4 | 4 | 4 |
| JS1086 | BTN | 3 | 3 | 3 | 3 |
| JS1087 | BTN | 3 | 3 | 3 | 3 |

| | | | | | |
|---|---|---|---|---|---|
| JS1088 | BTN | 3 | 3 | 3 | 3 |
| JS1089 | BTN | 3 | 3 | 3 | 3 |
| JS1090 | BTN | 4 | 4 | 4 | 4 |
| JS1091 | BTN | 4 | 4 | 4 | 4 |
| JS1092 | BTN | 4 | 4 | 4 | 4 |
| JS1093 | BTN | 3 | 3 | 3 | 3 |
| JS1094 | BTN | 4 | 4 | 3 | 4 |
| JS1095 | BTN | 4 | 3 | 3 | 3 |
| JS1096 | BTN | 4 | 3 | 4 | 4 |
| JS1097 | BTN | 4 | 3 | 4 | 4 |
| JS1098 | BTN | 3 | 4 | 4 | 4 |
| JS1099 | BTN | 4 | 4 | 4 | 4 |
| JS1100 | BTN | 2 | 3 | 2 | 2 |
| JS1101 | BTN | 3 | 3 | 3 | 3 |
| JS1102 | BTN | 3 | 3 | 3 | 3 |
| JS1103 | BTN | 3 | 3 | 3 | 3 |
| JS1104 | BTN | 3 | 3 | 3 | 3 |
| JS1105 | BTN | 4 | 3 | 3 | 3 |
| JS1106 | BTN | 4 | 4 | 4 | 4 |
| JS1107 | BTN | 4 | 4 | 4 | 4 |
| JS1108 | BTN | 4 | 4 | 4 | 4 |
| JS1109 | BTN | 3 | 3 | 3 | 3 |
| JS1110 | BTN | 4 | 4 | 4 | 4 |
| JS1111 | BTN | 3 | 3 | 3 | 3 |
| JS1112 | BTN | 2 | 2 | 2 | 2 |
| JS1113 | BTN | 4 | 4 | 4 | 4 |
| JS1114 | BTN | 5 | 5 | 4 | 5 |
| JS1115 | BTN | 3 | 3 | 3 | 3 |
| JS1116 | BTN | 2 | 3 | 3 | 3 |
| JS1117 | BTN | 3 | 4 | 3 | 3 |
| SH0001 | PTC | 4 | 4 | 4 | 4 |
| SH0005 | PTC | 4 | 4 | 4 | 4 |
| SH0006 | PTC | 4 | 4 | 4 | 4 |
| SH0013 | PTC | 5 | 5 | 5 | 5 |
| SH0017 | PTC | 4 | 4 | 4 | 4 |
| SH0018 | PTC | 2 | 2 | 2 | 2 |
| SH0020 | PTC | 4 | 4 | 4 | 4 |
| SH0022 | PTC | 4 | 4 | 4 | 4 |
| SH0024 | PTC | 4 | 5 | 5 | 5 |
| SH0028 | PTC | 5 | 5 | 5 | 5 |
| SH0030 | PTC | 4 | 4 | 4 | 4 |
| SH0032 | PTC | 4 | 4 | 4 | 4 |
| SH0035 | PTC | 5 | 5 | 5 | 5 |
| SH0036 | PTC | 3 | 4 | 4 | 4 |
| SH0037 | PTC | 4 | 4 | 4 | 4 |
| SH0041 | PTC | 5 | 5 | 5 | 5 |

| | | | | | |
|---|---|---|---|---|---|
| SH0043 | PTC | 4 | 4 | 4 | 4 |
| SH0044 | PTC | 5 | 5 | 5 | 5 |
| SH0045 | PTC | 5 | 5 | 5 | 5 |
| SH0046 | PTC | 4 | 4 | 4 | 4 |
| SH0047 | PTC | 4 | 4 | 4 | 4 |
| SH0049 | PTC | 4 | 4 | 4 | 4 |
| SH0052 | PTC | 4 | 4 | 4 | 4 |
| SH0053 | PTC | 4 | 4 | 4 | 4 |
| SH0054 | PTC | 5 | 5 | 5 | 5 |
| SH0056 | PTC | 5 | 5 | 5 | 5 |
| SH0057 | PTC | 4 | 3 | 4 | 4 |
| SH0059 | PTC | 4 | 4 | 4 | 4 |
| SH0061 | PTC | 4 | 4 | 4 | 4 |
| SH0062 | PTC | 4 | 4 | 4 | 4 |
| SH0065 | PTC | 4 | 4 | 4 | 4 |
| SH0072 | PTC | 4 | 4 | 4 | 4 |
| SH0076 | PTC | 4 | 4 | 4 | 4 |
| SH0078 | PTC | 4 | 5 | 4 | 4 |
| SH0079 | PTC | 5 | 5 | 5 | 5 |
| SH0080 | PTC | 4 | 4 | 4 | 4 |
| SH0085 | PTC | 4 | 3 | 4 | 4 |
| SH0087 | PTC | 4 | 4 | 4 | 4 |
| SH0089 | PTC | 4 | 4 | 4 | 4 |
| SH0090 | PTC | 5 | 5 | 5 | 5 |
| SH0092 | PTC | 4 | 5 | 5 | 5 |
| SH0093 | PTC | 4 | 4 | 4 | 4 |
| SH0097 | PTC | 4 | 4 | 4 | 4 |
| SH0098 | PTC | 4 | 4 | 4 | 4 |
| SH0099 | PTC | 4 | 4 | 4 | 4 |
| SH0101 | PTC | 4 | 4 | 4 | 4 |
| SH0103 | PTC | 4 | 4 | 4 | 4 |
| SH0104 | PTC | 4 | 4 | 4 | 4 |
| SH0105 | PTC | 3 | 3 | 3 | 3 |
| SH0106 | PTC | 4 | 4 | 4 | 4 |
| SH0108 | PTC | 5 | 5 | 5 | 5 |
| SH0109 | PTC | 3 | 3 | 3 | 3 |
| SH0110 | PTC | 4 | 4 | 4 | 4 |
| SH0111 | PTC | 4 | 4 | 4 | 4 |
| SH0113 | PTC | 4 | 4 | 4 | 4 |
| SH0114 | PTC | 4 | 4 | 4 | 4 |
| SH0116 | PTC | 3 | 3 | 3 | 3 |
| SH0117 | PTC | 4 | 4 | 4 | 4 |
| SH0120 | PTC | 3 | 3 | 3 | 3 |
| SH0123 | PTC | 5 | 5 | 5 | 5 |
| SH0124 | PTC | 4 | 4 | 4 | 4 |
| SH0125 | PTC | 5 | 5 | 5 | 5 |

| | | | | | |
|---|---|---|---|---|---|
| SH0127 | PTC | 5 | 5 | 5 | 5 |
| SH0128 | PTC | 4 | 4 | 4 | 4 |
| SH0132 | PTC | 5 | 5 | 5 | 5 |
| SH0137 | PTC | 4 | 4 | 4 | 4 |
| SH0139 | PTC | 5 | 5 | 4 | 5 |
| SH0143 | PTC | 4 | 4 | 4 | 4 |
| SH0144 | PTC | 5 | 5 | 5 | 5 |
| SH0145 | PTC | 4 | 4 | 4 | 4 |
| SH0146 | PTC | 5 | 5 | 5 | 5 |
| SH0156 | PTC | 4 | 4 | 4 | 4 |
| SH0157 | PTC | 5 | 5 | 5 | 5 |
| SH0159 | PTC | 4 | 3 | 4 | 4 |
| SH0161 | PTC | 5 | 5 | 5 | 5 |
| JS2001 | PTC | 4 | 5 | 5 | 5 |
| JS2004 | PTC | 3 | 4 | 4 | 4 |
| JS2005 | PTC | 4 | 5 | 5 | 5 |
| JS2008 | PTC | 3 | 4 | 4 | 4 |
| JS2010 | PTC | 2 | 4 | 4 | 4 |
| JS2013 | PTC | 2 | 4 | 4 | 4 |
| JS2014 | PTC | 4 | 5 | 5 | 5 |
| JS2016 | PTC | 4 | 5 | 5 | 5 |
| JS2018 | PTC | 5 | 4 | 4 | 4 |
| JS2019 | PTC | 4 | 3 | 3 | 3 |
| JS2022 | PTC | 3 | 4 | 4 | 4 |
| JS2023 | PTC | 2 | 3 | 3 | 3 |
| JS2024 | PTC | 2 | 4 | 4 | 4 |
| JS2025 | PTC | 5 | 4 | 4 | 4 |
| JS2026 | PTC | 4 | 3 | 4 | 4 |
| JS2027 | PTC | 4 | 5 | 5 | 5 |
| JS2028 | PTC | 4 | 5 | 5 | 5 |
| JS2029 | PTC | 4 | 2 | 4 | 4 |
| JS2030 | PTC | 4 | 3 | 4 | 4 |
| JS2031 | PTC | 4 | 5 | 4 | 4 |
| JS2032 | PTC | 4 | 5 | 4 | 4 |
| JS2033 | PTC | 3 | 4 | 3 | 3 |
| JS2036 | PTC | 3 | 5 | 5 | 5 |
| JS2037 | PTC | 4 | 3 | 4 | 4 |
| JS2039 | PTC | 3 | 3 | 2 | 3 |
| JS2040 | PTC | 3 | 5 | 5 | 5 |
| JS2041 | PTC | 3 | 5 | 5 | 5 |
| JS2042 | PTC | 3 | 5 | 5 | 5 |
| JS2043 | PTC | 3 | 5 | 3 | 3 |
| JS2044 | PTC | 4 | 3 | 3 | 3 |
| JS2046 | PTC | 4 | 5 | 4 | 4 |
| JS2048 | PTC | 3 | 5 | 5 | 5 |
| JS2049 | PTC | 3 | 3 | 3 | 3 |

| | | | | | |
|---|---|---|---|---|---|
| JS2051 | PTC | 4 | 5 | 4 | 4 |
| JS2052 | PTC | 3 | 3 | 4 | 3 |
| JS2053 | PTC | 4 | 5 | 4 | 4 |
| JS2054 | PTC | 3 | 5 | 5 | 5 |
| JS2056 | PTC | 4 | 5 | 5 | 5 |
| JS2057 | PTC | 4 | 5 | 4 | 4 |
| JS2058 | PTC | 3 | 3 | 3 | 3 |
| JS2061 | PTC | 3 | 4 | 3 | 3 |
| JS2062 | PTC | 4 | 4 | 5 | 4 |
| JS2063 | PTC | 4 | 4 | 3 | 4 |
| JS2065 | PTC | 3 | 3 | 3 | 3 |
| JS2067 | PTC | 4 | 3 | 3 | 3 |
| JS2069 | PTC | 4 | 4 | 3 | 4 |
| JS2070 | PTC | 4 | 5 | 5 | 5 |
| JS2071 | PTC | 3 | 3 | 3 | 3 |
| JS2074 | PTC | 3 | 5 | 5 | 5 |
| JS2076 | PTC | 3 | 2 | 3 | 3 |
| JS2077 | PTC | 3 | 3 | 2 | 3 |
| JS2078 | PTC | 4 | 4 | 3 | 4 |
| JS2083 | PTC | 4 | 4 | 3 | 4 |
| JS2084 | PTC | 3 | 5 | 5 | 5 |
| JS2085 | PTC | 4 | 5 | 5 | 5 |
| JS2086 | PTC | 3 | 3 | 4 | 3 |
| JS2087 | PTC | 4 | 4 | 5 | 4 |
| JS2089 | PTC | 4 | 4 | 5 | 4 |
| JS2091 | PTC | 4 | 4 | 5 | 4 |
| JS2093 | PTC | 4 | 5 | 5 | 5 |
| JS2095 | PTC | 4 | 3 | 3 | 3 |
| JS2096 | PTC | 4 | 4 | 5 | 4 |
| JS2098 | PTC | 3 | 5 | 5 | 5 |
| JS2100 | PTC | 4 | 3 | 3 | 3 |
| JS2101 | PTC | 3 | 5 | 5 | 5 |
| JS2102 | PTC | 4 | 4 | 3 | 4 |
| JS2103 | PTC | 5 | 3 | 5 | 5 |
| JS2104 | PTC | 4 | 4 | 2 | 4 |
| JS2105 | PTC | 3 | 2 | 3 | 3 |
| JS2106 | PTC | 3 | 2 | 3 | 3 |
| JS2107 | PTC | 4 | 4 | 5 | 4 |
| JS2108 | PTC | 3 | 4 | 3 | 3 |
| JS2110 | PTC | 3 | 3 | 4 | 3 |
| JS2112 | PTC | 3 | 3 | 4 | 3 |
| JS2114 | PTC | 4 | 4 | 5 | 4 |
| JS2115 | PTC | 3 | 3 | 2 | 3 |
| JS2117 | PTC | 4 | 4 | 5 | 4 |
| JS2119 | PTC | 4 | 3 | 3 | 3 |
| JS2121 | PTC | 4 | 4 | 3 | 4 |

| | | | | | |
|---|---|---|---|---|---|
| JS2123 | PTC | 2 | 3 | 3 | 3 |
| JS2125 | PTC | 2 | 3 | 3 | 3 |
| JS2127 | PTC | 4 | 3 | 4 | 4 |
| JS2128 | PTC | 5 | 4 | 5 | 5 |
| JS2129 | PTC | 5 | 4 | 5 | 5 |
| JS2130 | PTC | 4 | 3 | 3 | 3 |
| JS2132 | PTC | 4 | 5 | 4 | 4 |
| JS2133 | PTC | 4 | 3 | 4 | 4 |
| JS2134 | PTC | 3 | 2 | 3 | 3 |
| JS2135 | PTC | 4 | 5 | 4 | 4 |
| JS2137 | PTC | 5 | 4 | 5 | 5 |
| JS2139 | PTC | 3 | 2 | 3 | 3 |
| JS2140 | PTC | 4 | 3 | 4 | 4 |
| JS2141 | PTC | 3 | 4 | 3 | 3 |
| JS2146 | PTC | 4 | 5 | 4 | 4 |
| JS2147 | PTC | 5 | 4 | 5 | 5 |
| JS2149 | PTC | 4 | 2 | 4 | 4 |
| JS2151 | PTC | 4 | 5 | 4 | 4 |
| JS2154 | PTC | 4 | 3 | 4 | 4 |
| JS2158 | PTC | 5 | 3 | 5 | 5 |
| JS2161 | PTC | 4 | 5 | 4 | 4 |
| JS2162 | PTC | 4 | 2 | 4 | 4 |
| JS2164 | PTC | 3 | 4 | 3 | 3 |
| JS2167 | PTC | 5 | 3 | 5 | 5 |
| JS2171 | PTC | 4 | 3 | 4 | 4 |
| JS2175 | PTC | 4 | 5 | 4 | 4 |
| JS2180 | PTC | 5 | 4 | 5 | 5 |
| JS2181 | PTC | 5 | 4 | 5 | 5 |
| JS2182 | PTC | 5 | 4 | 5 | 5 |
| JS2184 | PTC | 5 | 4 | 4 | 4 |
| JS2185 | PTC | 5 | 3 | 5 | 5 |
| JS2187 | PTC | 5 | 3 | 5 | 5 |
| JS2188 | PTC | 5 | 4 | 5 | 5 |
| JS2191 | PTC | 3 | 4 | 4 | 4 |
| JS2192 | PTC | 5 | 4 | 5 | 5 |
| JS2194 | PTC | 5 | 4 | 4 | 4 |
| JS2195 | PTC | 3 | 4 | 4 | 4 |
| JS2197 | PTC | 5 | 4 | 5 | 5 |
| JS2198 | PTC | 5 | 4 | 4 | 4 |
| JS2199 | PTC | 2 | 4 | 4 | 4 |
| JS2201 | PTC | 3 | 3 | 2 | 3 |
| JS3003 | FTA | 3 | 3 | 4 | 3 |
| JS3005 | FTA | 3 | 3 | 4 | 3 |
| JS3007 | TFND | 3 | 3 | 2 | 3 |
| JS3011 | TFND | 3 | 3 | 4 | 3 |
| JS3016 | FTA | 3 | 3 | 2 | 3 |

| | | | | | |
|---|---|---|---|---|---|
| JS3027 | TFND | 3 | 4 | 4 | 4 |
| JS3029 | FTA | 4 | 3 | 3 | 3 |
| JS3032 | FTA | 5 | 4 | 4 | 4 |
| JS3035 | TFND | 2 | 3 | 3 | 3 |
| JS3040 | FTA | 3 | 4 | 4 | 4 |
| JS3044 | TFND | 3 | 3 | 3 | 3 |
| JS3047 | TFND | 5 | 4 | 4 | 4 |
| JS3049 | FTA | 5 | 3 | 5 | 5 |
| JS3052 | FTA | 4 | 3 | 3 | 3 |
| JS3055 | FTA | 5 | 4 | 4 | 4 |
| JS3062 | TFND | 5 | 5 | 4 | 5 |
| JS3065 | TFND | 5 | 4 | 4 | 4 |
| JS3070 | FTA | 2 | 3 | 3 | 3 |
| JS2002 | PTC | 5 | 4 | 4 | 4 |
| JS2003 | PTC | 3 | 4 | 4 | 4 |
| JS2006 | PTC | 5 | 5 | 3 | 5 |
| JS2007 | PTC | 5 | 5 | 4 | 5 |
| JS2009 | PTC | 3 | 4 | 4 | 4 |
| JS2011 | PTC | 3 | 4 | 4 | 4 |
| JS2012 | PTC | 5 | 5 | 3 | 5 |
| JS2015 | PTC | 3 | 4 | 4 | 4 |
| JS2017 | PTC | 5 | 5 | 4 | 5 |
| JS2020 | PTC | 3 | 3 | 3 | 3 |
| JS2021 | PTC | 2 | 3 | 3 | 3 |
| JS2034 | PTC | 5 | 5 | 3 | 5 |
| JS2035 | PTC | 5 | 4 | 4 | 4 |
| JS2038 | PTC | 3 | 4 | 4 | 4 |
| JS2045 | PTC | 3 | 4 | 4 | 4 |
| JS2047 | PTC | 5 | 5 | 4 | 5 |
| JS2050 | PTC | 5 | 5 | 4 | 5 |
| JS2055 | PTC | 5 | 5 | 4 | 5 |
| JS2059 | PTC | 5 | 5 | 4 | 5 |
| JS2060 | PTC | 5 | 3 | 3 | 3 |
| JS2064 | PTC | 3 | 2 | 3 | 3 |
| JS2066 | PTC | 3 | 4 | 3 | 3 |
| JS2068 | PTC | 3 | 4 | 4 | 4 |
| JS2072 | PTC | 5 | 4 | 4 | 4 |
| JS2073 | PTC | 5 | 4 | 4 | 4 |
| JS2079 | PTC | 4 | 4 | 5 | 4 |
| JS2080 | PTC | 4 | 4 | 5 | 4 |
| JS2081 | PTC | 3 | 3 | 3 | 3 |
| JS2082 | PTC | 5 | 5 | 3 | 5 |
| JS2088 | PTC | 4 | 4 | 5 | 4 |
| JS2097 | PTC | 3 | 4 | 3 | 3 |
| JS2109 | PTC | 3 | 3 | 3 | 3 |
| JS2116 | PTC | 4 | 4 | 5 | 4 |

| | | | | | |
|---|---|---|---|---|---|
| JS2120 | PTC | 4 | 4 | 5 | 4 |
| JS2122 | PTC | 3 | 2 | 3 | 3 |
| JS2124 | PTC | 4 | 4 | 5 | 4 |
| JS2126 | PTC | 3 | 3 | 3 | 3 |
| JS2138 | PTC | 4 | 4 | 3 | 4 |
| JS2142 | PTC | 4 | 4 | 3 | 4 |
| JS2144 | PTC | 5 | 5 | 4 | 5 |
| JS2145 | PTC | 4 | 4 | 3 | 4 |
| JS2148 | PTC | 4 | 4 | 3 | 4 |
| JS2152 | PTC | 5 | 3 | 3 | 3 |
| JS2159 | PTC | 4 | 4 | 3 | 4 |
| JS2186 | PTC | 4 | 4 | 3 | 4 |
| JS2189 | PTC | 3 | 3 | 3 | 3 |
| JS2190 | PTC | 4 | 5 | 4 | 4 |
| JS2193 | PTC | 4 | 3 | 4 | 4 |
| JS2196 | PTC | 4 | 2 | 4 | 4 |
| JS2200 | PTC | 5 | 5 | 4 | 5 |
| JS3001 | FTA | 4 | 5 | 4 | 4 |
| JS3002 | FTA | 3 | 3 | 4 | 3 |
| JS3004 | TFND | 3 | 3 | 2 | 3 |
| JS3006 | FTA | 4 | 3 | 4 | 4 |
| JS3008 | TFND | 2 | 3 | 3 | 3 |
| JS3009 | FTA | 5 | 5 | 4 | 5 |
| JS3010 | TFND | 5 | 5 | 4 | 5 |
| JS3012 | TFND | 5 | 3 | 3 | 3 |
| JS3013 | TFND | 4 | 3 | 3 | 3 |
| JS3014 | FTA | 2 | 3 | 3 | 3 |
| JS3015 | FTA | 4 | 5 | 4 | 4 |
| JS3017 | FTA | 3 | 3 | 3 | 3 |
| JS3018 | TFND | 3 | 2 | 3 | 3 |
| JS3019 | FTA | 4 | 3 | 4 | 4 |
| JS3020 | TFND | 3 | 5 | 3 | 3 |
| JS3021 | TFND | 3 | 5 | 3 | 3 |
| JS3022 | FTA | 4 | 5 | 4 | 4 |
| JS3023 | TFND | 3 | 4 | 3 | 3 |
| JS3024 | TFND | 3 | 2 | 3 | 3 |
| JS3025 | TFND | 4 | 5 | 4 | 4 |
| JS3026 | FTA | 3 | 5 | 3 | 3 |
| JS3028 | FTA | 3 | 3 | 3 | 3 |
| JS3030 | FTA | 4 | 5 | 4 | 4 |
| JS3031 | FTA | 4 | 3 | 3 | 3 |
| JS3033 | TFND | 3 | 3 | 2 | 3 |
| JS3034 | FTA | 3 | 3 | 2 | 3 |
| JS3036 | TFND | 4 | 3 | 3 | 3 |
| JS3037 | FTA | 4 | 5 | 4 | 4 |
| JS3038 | TFND | 4 | 3 | 4 | 4 |

| | | | | | |
|---|---|---|---|---|---|
| JS3039 | TFND | 4 | 3 | 3 | 3 |
| JS3041 | FTA | 4 | 3 | 3 | 3 |
| JS3042 | FTA | 4 | 3 | 3 | 3 |
| JS3043 | TFND | 3 | 4 | 4 | 4 |
| JS3045 | FTA | 3 | 4 | 4 | 4 |
| JS3046 | TFND | 4 | 3 | 3 | 3 |
| JS3048 | FTA | 3 | 2 | 3 | 3 |
| JS3050 | FTA | 3 | 4 | 4 | 4 |
| JS3051 | FTA | 3 | 2 | 3 | 3 |
| JS3053 | TFND | 3 | 2 | 3 | 3 |
| JS3054 | FTA | 3 | 3 | 4 | 3 |
| JS3056 | FTA | 5 | 4 | 4 | 4 |
| JS3057 | TFND | 2 | 3 | 3 | 3 |
| JS3058 | TFND | 3 | 3 | 4 | 3 |
| JS3059 | FTA | 3 | 4 | 4 | 4 |
| JS3060 | TFND | 3 | 4 | 4 | 4 |
| JS3061 | FTA | 2 | 3 | 3 | 3 |
| JS3063 | TFND | 3 | 3 | 4 | 3 |
| JS3064 | FTA | 3 | 2 | 3 | 3 |
| JS3066 | FTA | 3 | 4 | 3 | 3 |
| JS3067 | TFND | 5 | 4 | 4 | 4 |
| JS3068 | TFND | 3 | 3 | 3 | 3 |
| JS3069 | TFND | 3 | 4 | 4 | 4 |
| JS3071 | FTA | 3 | 4 | 3 | 3 |
| JS3072 | FTA | 3 | 3 | 3 | 3 |
| JS3073 | FTA | 3 | 4 | 3 | 3 |
| JS3074 | TFND | 3 | 3 | 3 | 3 |
| JS3075 | FTA | 3 | 2 | 3 | 3 |
| JS3076 | TFND | 5 | 5 | 4 | 5 |
| JS3077 | FTA | 3 | 4 | 4 | 4 |
| JS3078 | TFND | 4 | 3 | 3 | 3 |
| JS6812 | PTC | 4 | 4 | 4 | 4 |
| JS7400 | PTC | 4 | 4 | 4 | 4 |
| JS7284 | PTC | 5 | 5 | 5 | 5 |
| JS7372 | PTC | 4 | 4 | 4 | 4 |
| JS7160 | PTC | 4 | 4 | 4 | 4 |
| JS6731 | PTC | 4 | 4 | 4 | 4 |
| JS7063 | PTC | 4 | 5 | 5 | 5 |
| JS7031 | PTC | 4 | 4 | 4 | 4 |
| JS6885 | PTC | 4 | 4 | 4 | 4 |
| JS7107 | PTC | 4 | 5 | 5 | 5 |
| JS6845 | FTC | 4 | 4 | 4 | 4 |
| JS7144 | PTC | 4 | 4 | 4 | 4 |
| JS7175 | PTC | 4 | 4 | 4 | 4 |
| JS7335 | PTC | 4 | 4 | 4 | 4 |
| JS7222 | PTC | 4 | 4 | 4 | 4 |

| | | | | | |
|---|---|---|---|---|---|
| JS6802 | PTC | 4 | 4 | 4 | 4 |
| JS7055 | PTC | 4 | 4 | 4 | 4 |
| JS6882 | PTC | 4 | 5 | 5 | 5 |
| JS7177 | TFND | 4 | 4 | 4 | 4 |
| JS7086 | PTC | 4 | 4 | 4 | 4 |
| JS7161 | PTC | 4 | 4 | 4 | 4 |
| JS6862 | PTC | 4 | 4 | 4 | 4 |
| JS7361 | PTC | 4 | 5 | 5 | 5 |
| JS7038 | PTC | 4 | 4 | 4 | 4 |
| JS7098 | PTC | 4 | 4 | 4 | 4 |
| JS6808 | PTC | 4 | 4 | 4 | 4 |
| JS7300 | PTC | 5 | 5 | 5 | 5 |
| JS6732 | PTC | 4 | 4 | 4 | 4 |
| JS7428 | PTC | 4 | 4 | 4 | 4 |
| JS7020 | PTC | 4 | 4 | 4 | 4 |
| JS7336 | PTC | 5 | 5 | 4 | 5 |
| JS7047 | PTC | 4 | 4 | 4 | 4 |
| JS7075 | PTC | 4 | 4 | 4 | 4 |
| JS6720 | PTC | 4 | 5 | 4 | 4 |
| JS7465 | PTC | 4 | 4 | 4 | 4 |
| JS7234 | PTC | 4 | 4 | 4 | 4 |
| JS7029 | PTC | 4 | 4 | 5 | 4 |
| JS6877 | PTC | 4 | 4 | 4 | 4 |
| JS7329 | PTC | 5 | 5 | 4 | 5 |
| JS7045 | PTC | 4 | 4 | 3 | 4 |
| JS7441 | PTC | 5 | 5 | 3 | 5 |
| JS6841 | PTC | 5 | 5 | 4 | 5 |
| JS6846 | PTC | 4 | 5 | 5 | 5 |
| JS6807 | PTC | 4 | 4 | 3 | 4 |
| JS6866 | PTC | 3 | 4 | 4 | 4 |
| JS7314 | PTC | 3 | 4 | 4 | 4 |
| JS7347 | PTC | 4 | 4 | 4 | 4 |
| JS7339 | PTC | 4 | 4 | 4 | 4 |
| JS7181 | PTC | 4 | 4 | 4 | 4 |
| JS7440 | PTC | 5 | 5 | 4 | 5 |
| JS7455 | PTC | 5 | 5 | 4 | 5 |
| JS7230 | TFND | 4 | 4 | 4 | 4 |
| JS7288 | PTC | 4 | 4 | 4 | 4 |
| JS7256 | PTC | 4 | 4 | 4 | 4 |
| JS6887 | PTC | 4 | 4 | 4 | 4 |
| JS7186 | PTC | 4 | 4 | 4 | 4 |
| JS7199 | TFND | 4 | 4 | 4 | 4 |
| JS6848 | TFND | 4 | 4 | 4 | 4 |
| JS7194 | PTC | 4 | 4 | 4 | 4 |
| JS7423 | PTC | 4 | 4 | 4 | 4 |
| JS7424 | PTC | 4 | 4 | 4 | 4 |

| | | | | | |
|---|---|---|---|---|---|
| JS7179 | PTC | 4 | 4 | 4 | 4 |
| JS6895 | PTC | 4 | 4 | 4 | 4 |
| JS7324 | PTC | 4 | 3 | 4 | 4 |
| JS6803 | PTC | 4 | 4 | 4 | 4 |
| JS6891 | FTA | 4 | 4 | 4 | 4 |
| JS7419 | PTC | 4 | 4 | 5 | 4 |
| JS7191 | PTC | 4 | 4 | 4 | 4 |
| JS7285 | PTC | 4 | 4 | 4 | 4 |
| JS7262 | PTC | 3 | 3 | 3 | 3 |
| JS7427 | PTC | 4 | 4 | 4 | 4 |
| JS7074 | PTC | 4 | 4 | 3 | 4 |
| JS7219 | PTC | 4 | 4 | 4 | 4 |
| JS6729 | PTC | 4 | 4 | 4 | 4 |
| JS7124 | PTC | 4 | 4 | 4 | 4 |
| JS7286 | PTC | 4 | 4 | 4 | 4 |
| JS7126 | PTC | 4 | 5 | 5 | 5 |
| JS6797 | PTC | 4 | 4 | 3 | 4 |
| JS7121 | PTC | 5 | 5 | 4 | 5 |
| JS6734 | PTC | 4 | 4 | 5 | 4 |
| JS7238 | TFND | 3 | 3 | 3 | 3 |
| JS7235 | FTA | 2 | 3 | 3 | 3 |
| JS6850 | PTC | 3 | 4 | 4 | 4 |
| JS7145 | PTC | 4 | 4 | 5 | 4 |
| JS7218 | PTC | 4 | 4 | 4 | 4 |
| JS7198 | PTC | 5 | 5 | 5 | 5 |
| JS7357 | PTC | 4 | 4 | 5 | 4 |
| JS7174 | PTC | 3 | 3 | 2 | 3 |
| JS7272 | TFND | 4 | 4 | 3 | 4 |
| JS7127 | PTC | 4 | 4 | 4 | 4 |
| JS6886 | FTA | 3 | 3 | 3 | 3 |
| JS7379 | PTC | 4 | 3 | 4 | 4 |
| JS7275 | FTA | 3 | 4 | 3 | 3 |
| JS6832 | FTA | 4 | 4 | 4 | 4 |
| JS7257 | PTC | 4 | 4 | 4 | 4 |
| JS6723 | TFND | 4 | 4 | 4 | 4 |
| JS7343 | PTC | 4 | 4 | 4 | 4 |
| JS7382 | PTC | 4 | 4 | 4 | 4 |
| JS7334 | PTC | 4 | 4 | 4 | 4 |
| JS7330 | PTC | 4 | 3 | 3 | 3 |
| JS6779 | PTC | 4 | 4 | 4 | 4 |
| JS6706 | PTC | 4 | 4 | 4 | 4 |
| JS7340 | PTC | 4 | 4 | 4 | 4 |
| JS7326 | PTC | 4 | 4 | 4 | 4 |
| JS7103 | TFND | 4 | 4 | 4 | 4 |
| JS7140 | FTA | 3 | 3 | 4 | 3 |
| JS7387 | PTC | 4 | 4 | 4 | 4 |

| | | | | | |
|---|---|---|---|---|---|
| JS7147 | TFND | 4 | 4 | 3 | 4 |
| JS7362 | TFND | 3 | 3 | 4 | 3 |
| JS7466 | PTC | 4 | 4 | 4 | 4 |
| JS7290 | PTC | 4 | 5 | 4 | 4 |
| JS7071 | TFND | 3 | 2 | 3 | 3 |
| JS7012 | PTC | 4 | 4 | 5 | 4 |
| JS6894 | TFND | 3 | 3 | 2 | 3 |
| JS7282 | FTA | 2 | 3 | 3 | 3 |
| JS7239 | FTA | 2 | 2 | 2 | 2 |
| JS7443 | PTC | 4 | 4 | 4 | 4 |
| JS7125 | TFND | 4 | 4 | 4 | 4 |
| JS7429 | PTC | 4 | 4 | 4 | 4 |
| JS7446 | TFND | 4 | 4 | 4 | 4 |
| JS7193 | TFND | 4 | 4 | 4 | 4 |
| JS6730 | FTA | 3 | 2 | 3 | 3 |
| JS6812 | PTC | 4 | 4 | 3 | 4 |
| JS7400 | PTC | 4 | 4 | 3 | 4 |
| JS7284 | PTC | 5 | 5 | 3 | 5 |

**Supplementary Table S8.** Independent ACR TI-RADS review results of thyroid ultrasonographic imaging in the model training group without histopathologically confirmed or FNAB diagnosis by three senior ultrasound radiologists

| Case ID | Final Histology | ACR TI-RADS Categories | | | Final ACR TI-RADS |
|---|---|---|---|---|---|
| | | Dr. Lou | Dr. Jin | Dr. Lyu | |
| AH0278 | PTC | 5 | 4 | 4 | 4 |
| AH0201 | PTC | 3 | 4 | 4 | 4 |
| AH0256 | FTC | 3 | 2 | 3 | 3 |
| SH0066 | PTC | 3 | 4 | 4 | 4 |
| AH0324 | PTC | 5 | 5 | 5 | 5 |
| AH0137 | PTC | 4 | 5 | 4 | 4 |
| AH0219 | PTC | 4 | 4 | 3 | 4 |
| AH0182 | PTC | 4 | 5 | 5 | 5 |
| AH0276 | PTC | 5 | 5 | 4 | 5 |
| AH0223 | PTC | 4 | 5 | 4 | 4 |
| AH0109 | PTC | 5 | 4 | 4 | 4 |
| AH0206 | PTC | 3 | 4 | 4 | 4 |
| AH0326 | PTC | 5 | 4 | 5 | 5 |
| AH0100 | PTC | 5 | 4 | 4 | 4 |
| AH0270 | PTC | 4 | 4 | 3 | 4 |
| AH0320 | PTC | 4 | 5 | 5 | 5 |
| AH0310 | PTC | 3 | 3 | 4 | 3 |
| AH0202 | PTC | 5 | 4 | 4 | 4 |
| AH0285 | PTC | 4 | 4 | 5 | 4 |

| | | | | | |
|---|---|---|---|---|---|
| AH0252 | PTC | 4 | 4 | 4 | 4 |
| AH0022 | PTC | 4 | 4 | 5 | 4 |
| AH0212 | PTC | 4 | 4 | 5 | 4 |
| AH0319 | PTC | 5 | 3 | 5 | 5 |
| AH0258 | PTC | 4 | 5 | 5 | 5 |
| AH0213 | PTC | 4 | 5 | 4 | 4 |
| AH0354 | PTC | 3 | 4 | 4 | 4 |
| AH0163 | PTC | 3 | 4 | 4 | 4 |
| AH0172 | PTC | 3 | 4 | 4 | 4 |
| AH0099 | PTC | 4 | 4 | 5 | 4 |
| AH0195 | PTC | 4 | 5 | 4 | 4 |
| AH0155 | PTC | 4 | 5 | 5 | 5 |
| AH0260 | PTC | 3 | 4 | 4 | 4 |
| AH0136 | PTC | 4 | 5 | 5 | 5 |
| AH0108 | PTC | 4 | 5 | 4 | 4 |
| AH0173 | PTC | 4 | 4 | 5 | 4 |
| AH0315 | PTC | 3 | 4 | 4 | 4 |
| AH0171 | PTC | 3 | 4 | 3 | 3 |
| AH0283 | PTC | 4 | 4 | 3 | 4 |
| AH0308 | PTC | 4 | 4 | 3 | 4 |
| AH0332 | PTC | 4 | 3 | 4 | 4 |
| AH0307 | PTC | 5 | 4 | 5 | 5 |
| AH0178 | PTC | 3 | 4 | 4 | 4 |
| AH0359 | PTC | 3 | 4 | 4 | 4 |
| AH0318 | PTC | 4 | 5 | 5 | 5 |
| AH0175 | PTC | 3 | 4 | 4 | 4 |
| AH0263 | PTC | 4 | 5 | 4 | 4 |
| AH0047 | BTN | 3 | 3 | 4 | 3 |
| AH0224 | PTC | 3 | 4 | 4 | 4 |
| SH0084 | PTC | 4 | 5 | 5 | 5 |
| AH0153 | BTN | 3 | 4 | 4 | 4 |
| AH0292 | BTN | 3 | 4 | 3 | 3 |
| AH0048 | BTN | 3 | 4 | 4 | 4 |
| AH0295 | BTN | 4 | 4 | 5 | 4 |
| AH0179 | PTC | 3 | 4 | 4 | 4 |
| AH0046 | BTN | 4 | 3 | 4 | 4 |
| AH0079 | BTN | 4 | 4 | 5 | 4 |
| AH0328 | PTC | 4 | 5 | 4 | 4 |
| AH0316 | PTC | 4 | 5 | 4 | 4 |
| AH0341 | PTC | 4 | 5 | 5 | 5 |
| AH0141 | BTN | 3 | 4 | 4 | 4 |
| AH0196 | PTC | 3 | 4 | 4 | 4 |
| AH0169 | PTC | 4 | 5 | 5 | 5 |
| AH0101 | PTC | 4 | 4 | 5 | 4 |
| AH0304 | PTC | 4 | 5 | 4 | 4 |

| | | | | | |
|---|---|---|---|---|---|
| AH0350 | BTN | 4 | 3 | 3 | 3 |
| SH0081 | PTC | 4 | 5 | 5 | 5 |
| AH0364 | PTC | 3 | 4 | 4 | 4 |
| AH0211 | PTC | 4 | 4 | 5 | 4 |
| AH0353 | BTN | 5 | 4 | 4 | 4 |
| AH0144 | BTN | 3 | 4 | 3 | 3 |
| AH0003 | PTC | 3 | 4 | 4 | 4 |
| AH0185 | PTC | 4 | 4 | 5 | 4 |
| AH0265 | PTC | 4 | 3 | 4 | 4 |
| AH0131 | PTC | 4 | 3 | 4 | 4 |
| AH0323 | PTC | 4 | 3 | 4 | 4 |
| AH0177 | PTC | 5 | 4 | 4 | 4 |
| AH0287 | PTC | 4 | 4 | 3 | 4 |
| AH0266 | PTC | 4 | 5 | 5 | 5 |
| AH0306 | PTC | 4 | 5 | 5 | 5 |
| AH0284 | PTC | 3 | 4 | 4 | 4 |
| AH0351 | BTN | 3 | 4 | 4 | 4 |
| AH0314 | PTC | 3 | 4 | 4 | 4 |
| AH0360 | PTC | 4 | 5 | 5 | 5 |
| AH0025 | PTC | 4 | 5 | 4 | 4 |
| AH0362 | PTC | 5 | 5 | 4 | 5 |
| AH0329 | PTC | 5 | 5 | 4 | 5 |
| AH0024 | PTC | 3 | 4 | 4 | 4 |
| AH0339 | PTC | 3 | 4 | 4 | 4 |
| SH0031 | PTC | 5 | 5 | 5 | 5 |
| AH0125 | PTC | 3 | 4 | 4 | 4 |
| AH0093 | BTN | 3 | 4 | 4 | 4 |
| AH0301 | PTC | 3 | 4 | 4 | 4 |
| AH0208 | PTC | 4 | 3 | 4 | 4 |
| AH0268 | PTC | 4 | 4 | 4 | 4 |
| AH0215 | PTC | 4 | 3 | 4 | 4 |
| AH0239 | PTC | 3 | 2 | 3 | 3 |
| AH0162 | PTC | 3 | 4 | 4 | 4 |
| AH0078 | BTN | 4 | 4 | 5 | 4 |
| AH0007 | PTC | 4 | 4 | 4 | 4 |
| AH0180 | PTC | 3 | 4 | 4 | 4 |
| AH0229 | PTC | 3 | 4 | 4 | 4 |
| AH0034 | PTC | 4 | 4 | 5 | 4 |
| AH0282 | PTC | 4 | 3 | 4 | 4 |
| AH0352 | BTN | 4 | 3 | 4 | 4 |
| AH0037 | PTC | 3 | 4 | 4 | 4 |
| AH0247 | PTC | 4 | 4 | 4 | 4 |
| AH0075 | BTN | 3 | 3 | 4 | 3 |
| AH0105 | PTC | 4 | 4 | 4 | 4 |
| AH0158 | PTC | 3 | 4 | 4 | 4 |

| | | | | | |
|---|---|---|---|---|---|
| AH0011 | PTC | 3 | 4 | 4 | 4 |
| AH0029 | PTC | 4 | 4 | 5 | 4 |
| SH0091 | PTC | 4 | 4 | 4 | 4 |
| SH0082 | PTC | 4 | 5 | 5 | 5 |
| AH0193 | PTC | 4 | 5 | 4 | 4 |
| AH0288 | PTC | 3 | 4 | 4 | 4 |
| AH0231 | PTC | 3 | 4 | 4 | 4 |
| AH0036 | PTC | 4 | 5 | 4 | 4 |
| AH0112 | PTC | 3 | 4 | 4 | 4 |
| AH0251 | PTC | 3 | 4 | 4 | 4 |
| AH0161 | PTC | 4 | 4 | 5 | 4 |
| AH0129 | PTC | 4 | 5 | 4 | 4 |
| AH0015 | BTN | 3 | 4 | 4 | 4 |
| SH0212 | PTC | 4 | 3 | 3 | 3 |
| SH0202 | PTC | 4 | 5 | 5 | 5 |
| AH0052 | BTN | 4 | 4 | 4 | 4 |
| SH0168 | PTC | 4 | 5 | 5 | 5 |
| AH0084 | BTN | 4 | 4 | 5 | 4 |
| SH0199 | PTC | 5 | 5 | 5 | 5 |
| SH0175 | PTC | 4 | 5 | 5 | 5 |
| AH0142 | BTN | 4 | 4 | 4 | 4 |
| SH0136 | PTC | 5 | 5 | 5 | 5 |
| SH0208 | BTN | 5 | 4 | 5 | 5 |
| SH0207 | BTN | 3 | 3 | 3 | 3 |
| SH0170 | PTC | 4 | 4 | 4 | 4 |
| SH0172 | PTC | 5 | 5 | 5 | 5 |
| SH0058 | PTC | 4 | 3 | 3 | 3 |
| SH0171 | PTC | 5 | 5 | 5 | 5 |
| AH0151 | BTN | 3 | 4 | 3 | 3 |
| AH0249 | PTC | 4 | 3 | 4 | 4 |
| AH0235 | PTC | 3 | 4 | 4 | 4 |
| SH0188 | PTC | 5 | 5 | 5 | 5 |
| AH0133 | PTC | 4 | 4 | 4 | 4 |
| AH0032 | PTC | 4 | 3 | 4 | 4 |
| AH0113 | PTC | 4 | 4 | 4 | 4 |
| AH0130 | PTC | 5 | 5 | 5 | 5 |
| SH0173 | PTC | 5 | 5 | 5 | 5 |
| AH0242 | PTC | 3 | 2 | 3 | 3 |
| AH0092 | BTN | 3 | 3 | 3 | 3 |
| SH0195 | PTC | 5 | 5 | 5 | 5 |
| SH0121 | PTC | 4 | 5 | 5 | 5 |
| SH0192 | PTC | 5 | 5 | 5 | 5 |
| SH0193 | PTC | 4 | 4 | 5 | 4 |
| AH0038 | PTC | 4 | 4 | 4 | 4 |
| AH0210 | PTC | 4 | 3 | 4 | 4 |

| | | | | | |
|---|---|---|---|---|---|
| AH0416 | BTN | 3 | 4 | 3 | 3 |
| AH0274 | PTC | 4 | 4 | 4 | 4 |
| AH0204 | PTC | 3 | 4 | 4 | 4 |
| SH0077 | PTC | 5 | 5 | 4 | 5 |
| SH0189 | PTC | 4 | 5 | 5 | 5 |
| AH0355 | PTC | 4 | 3 | 4 | 4 |
| AH0027 | PTC | 4 | 4 | 4 | 4 |
| AH0241 | PTC | 4 | 4 | 4 | 4 |
| SH0197 | PTC | 5 | 5 | 5 | 5 |
| SH0190 | PTC | 4 | 5 | 5 | 5 |
| SH0174 | PTC | 3 | 4 | 4 | 4 |
| SH0100 | PTC | 5 | 5 | 5 | 5 |
| AH0157 | PTC | 4 | 5 | 5 | 5 |
| JS1019 | BTN | 3 | 4 | 4 | 4 |
| SH0198 | PTC | 5 | 5 | 4 | 5 |
| SH0086 | PTC | 5 | 5 | 5 | 5 |
| AH0117 | PTC | 4 | 4 | 4 | 4 |
| SH0169 | PTC | 4 | 3 | 4 | 4 |
| AH0008 | BTN | 3 | 4 | 4 | 4 |
| SH0196 | PTC | 4 | 4 | 4 | 4 |
| SH0095 | PTC | 4 | 4 | 4 | 4 |
| AH0035 | PTC | 3 | 4 | 4 | 4 |
| AH0132 | PTC | 4 | 4 | 4 | 4 |
| SH0187 | PTC | 3 | 4 | 4 | 4 |
| AH0232 | PTC | 4 | 5 | 4 | 4 |
| AH0356 | PTC | 5 | 5 | 5 | 5 |
| AH0198 | PTC | 4 | 4 | 5 | 4 |
| AH0368 | BTN | 4 | 4 | 4 | 4 |
| SH0069 | PTC | 3 | 4 | 4 | 4 |
| SH0027 | PTC | 5 | 4 | 5 | 5 |
| AH0363 | PTC | 4 | 5 | 5 | 5 |
| AH0289 | PTC | 4 | 4 | 4 | 4 |
| AH0005 | PTC | 5 | 5 | 5 | 5 |
| AH0209 | PTC | 4 | 3 | 4 | 4 |
| AH0338 | PTC | 3 | 4 | 4 | 4 |
| AH0004 | PTC | 4 | 4 | 4 | 4 |
| AH0228 | PTC | 4 | 3 | 4 | 4 |
| AH0128 | PTC | 3 | 4 | 4 | 4 |
| AH0030 | PTC | 4 | 3 | 4 | 4 |
| AH0343 | BTN | 3 | 3 | 3 | 3 |
| AH0367 | BTN | 4 | 4 | 5 | 4 |
| SH0067 | FTC | 2 | 2 | 4 | 2 |
| AH0333 | BTN | 3 | 3 | 3 | 3 |
| SH0060 | PTC | 5 | 5 | 5 | 5 |
| SH0096 | PTC | 5 | 5 | 5 | 5 |

| | | | | | |
|---|---|---|---|---|---|
| AH0010 | BTN | 4 | 4 | 4 | 4 |
| SH0038 | PTC | 5 | 5 | 4 | 5 |
| AH0240 | PTC | 3 | 4 | 4 | 4 |
| AH0149 | BTN | 3 | 3 | 3 | 3 |
| SH0181 | PTC | 4 | 5 | 5 | 5 |
| AH0026 | PTC | 4 | 4 | 5 | 4 |
| AH0214 | PTC | 4 | 5 | 4 | 4 |
| AH0226 | PTC | 4 | 4 | 4 | 4 |
| SH0213 | BTN | 4 | 3 | 3 | 3 |
| AH0238 | PTC | 3 | 4 | 4 | 4 |
| SH0186 | PTC | 4 | 5 | 5 | 5 |
| SH0206 | BTN | 3 | 3 | 3 | 3 |
| SH0183 | PTC | 4 | 5 | 5 | 5 |
| SH0178 | PTC | 4 | 5 | 5 | 5 |
| SH0180 | PTC | 3 | 4 | 3 | 3 |
| AH0145 | BTN | 3 | 4 | 4 | 4 |
| SH0215 | BTN | 4 | 4 | 4 | 4 |
| SH0185 | PTC | 4 | 5 | 5 | 5 |
| SH0204 | PTC | 4 | 3 | 4 | 4 |
| SH0214 | BTN | 3 | 3 | 2 | 3 |
| SH0177 | PTC | 4 | 5 | 5 | 5 |
| SH0176 | PTC | 4 | 4 | 3 | 4 |
| SH0194 | PTC | 5 | 5 | 4 | 5 |
| AH0402 | BTN | 3 | 4 | 4 | 4 |
| SH0040 | PTC | 4 | 5 | 5 | 5 |
| AH0055 | BTN | 3 | 4 | 4 | 4 |
| AH0317 | FTC | 3 | 2 | 3 | 3 |
| SH0029 | PTC | 3 | 4 | 4 | 4 |
| SH0070 | PTC | 5 | 4 | 5 | 5 |
| AH0138 | PTC | 4 | 4 | 3 | 4 |
| AH0245 | PTC | 4 | 4 | 3 | 4 |
| SH0164 | PTC | 4 | 4 | 4 | 4 |
| AH0056 | BTN | 4 | 3 | 4 | 4 |
| AH0303 | PTC | 3 | 4 | 4 | 4 |
| AH0384 | BTN | 2 | 3 | 3 | 3 |
| AH0207 | PTC | 3 | 4 | 4 | 4 |
| AH0054 | BTN | 4 | 4 | 4 | 4 |
| AH0216 | PTC | 4 | 4 | 5 | 4 |
| SH0034 | PTC | 4 | 5 | 5 | 5 |
| AH0227 | PTC | 4 | 3 | 4 | 4 |
| SH0063 | PTC | 4 | 5 | 5 | 5 |
| AH0296 | BTN | 5 | 4 | 4 | 4 |
| SH0074 | PTC | 3 | 4 | 4 | 4 |
| SH0138 | PTC | 3 | 3 | 3 | 3 |
| AH0253 | PTC | 4 | 5 | 4 | 4 |

| | | | | | |
|---|---|---|---|---|---|
| SH0071 | PTC | 5 | 4 | 5 | 5 |
| SH0075 | PTC | 5 | 4 | 5 | 5 |
| AH0250 | PTC | 4 | 5 | 4 | 4 |
| SH0042 | PTC | 4 | 5 | 5 | 5 |
| SH0107 | PTC | 4 | 5 | 5 | 5 |
| AH0188 | PTC | 3 | 4 | 4 | 4 |
| AH0028 | PTC | 4 | 4 | 4 | 4 |
| AH0230 | PTC | 4 | 5 | 4 | 4 |
| AH0031 | PTC | 3 | 4 | 4 | 4 |
| SH0083 | PTC | 4 | 5 | 4 | 4 |
| SH0130 | PTC | 3 | 4 | 4 | 4 |
| SH0073 | PTC | 4 | 5 | 4 | 4 |
| AH0049 | BTN | 4 | 3 | 4 | 4 |
| AH0041 | BTN | 2 | 3 | 3 | 3 |
| AH0347 | BTN | 3 | 4 | 4 | 4 |
| AH0009 | PTC | 4 | 4 | 5 | 4 |
| SH0039 | PTC | 4 | 4 | 5 | 4 |
| SH0150 | PTC | 5 | 4 | 5 | 5 |
| AH0186 | PTC | 5 | 4 | 4 | 4 |
| SH0166 | PTC | 3 | 3 | 4 | 3 |
| AH0221 | PTC | 3 | 4 | 4 | 4 |
| SH0152 | PTC | 5 | 5 | 5 | 5 |
| SH0014 | PTC | 4 | 4 | 4 | 4 |
| AH0110 | PTC | 4 | 4 | 4 | 4 |
| AH0121 | PTC | 3 | 4 | 3 | 3 |
| SH0003 | PTC | 4 | 4 | 3 | 4 |
| SH0149 | PTC | 4 | 4 | 3 | 4 |
| AH0225 | PTC | 3 | 4 | 4 | 4 |
| SH0135 | PTC | 4 | 5 | 5 | 5 |
| SH0112 | PTC | 4 | 5 | 5 | 5 |
| SH0131 | PTC | 3 | 4 | 4 | 4 |
| AH0104 | PTC | 4 | 3 | 4 | 4 |
| AH0233 | PTC | 4 | 3 | 4 | 4 |
| SH0025 | PTC | 4 | 4 | 3 | 4 |
| SH0153 | PTC | 4 | 4 | 3 | 4 |
| SH0016 | PTC | 3 | 4 | 4 | 4 |
| SH0051 | PTC | 4 | 5 | 5 | 5 |
| SH0094 | PTC | 4 | 5 | 5 | 5 |
| AH0244 | PTC | 3 | 4 | 4 | 4 |
| SH0142 | PTC | 4 | 5 | 5 | 5 |
| SH0102 | PTC | 3 | 4 | 4 | 4 |
| SH0122 | PTC | 4 | 4 | 5 | 4 |
| SH0064 | PTC | 4 | 5 | 4 | 4 |
| AH0218 | PTC | 3 | 4 | 4 | 4 |
| SH0151 | PTC | 4 | 5 | 5 | 5 |

| | | | | | |
|---|---|---|---|---|---|
| AH0377 | BTN | 3 | 4 | 4 | 4 |
| AH0058 | BTN | 3 | 4 | 4 | 4 |
| AH0124 | PTC | 4 | 5 | 4 | 4 |
| AH0152 | BTN | 4 | 5 | 4 | 4 |
| AH0281 | PTC | 3 | 4 | 4 | 4 |
| AH0234 | PTC | 4 | 4 | 5 | 4 |
| AH0119 | PTC | 3 | 4 | 4 | 4 |
| SH0050 | PTC | 4 | 4 | 4 | 4 |
| SH0165 | PTC | 5 | 5 | 5 | 5 |
| SH0055 | PTC | 4 | 5 | 5 | 5 |
| AH0059 | BTN | 3 | 4 | 3 | 3 |
| SH0119 | PTC | 3 | 4 | 4 | 4 |
| AH0187 | PTC | 4 | 4 | 4 | 4 |
| AH0412 | BTN | 4 | 3 | 4 | 4 |
| AH0114 | PTC | 4 | 4 | 4 | 4 |
| SH0088 | PTC | 4 | 4 | 5 | 4 |
| AH0200 | PTC | 4 | 4 | 4 | 4 |
| SH0155 | PTC | 4 | 5 | 5 | 5 |
| SH0140 | PTC | 5 | 4 | 5 | 5 |
| SH0134 | PTC | 3 | 4 | 4 | 4 |
| AH0248 | PTC | 3 | 4 | 4 | 4 |
| AH0122 | PTC | 5 | 4 | 5 | 5 |
| SH0129 | PTC | 3 | 3 | 2 | 3 |
| AH0243 | PTC | 3 | 4 | 4 | 4 |
| SH0008 | PTC | 5 | 5 | 4 | 5 |
| SH0158 | PTC | 4 | 3 | 4 | 4 |
| AH0297 | BTN | 5 | 5 | 4 | 5 |
| AH0409 | BTN | 4 | 3 | 4 | 4 |
| AH0237 | PTC | 3 | 4 | 4 | 4 |
| AH0418 | BTN | 3 | 3 | 3 | 3 |
| SH0162 | PTC | 4 | 5 | 5 | 5 |
| SH0068 | PTC | 4 | 3 | 4 | 4 |
| SH0007 | PTC | 3 | 4 | 4 | 4 |
| SH0048 | PTC | 4 | 5 | 5 | 5 |
| SH0126 | PTC | 4 | 5 | 5 | 5 |
| AH0217 | PTC | 4 | 3 | 4 | 4 |
| AH0199 | PTC | 4 | 3 | 4 | 4 |
| SH0133 | PTC | 4 | 3 | 4 | 4 |
| SH0154 | PTC | 3 | 3 | 3 | 3 |
| AH0236 | PTC | 3 | 4 | 4 | 4 |
| SH0115 | PTC | 4 | 4 | 4 | 4 |
| AH0098 | PTC | 4 | 5 | 4 | 4 |
| SH0009 | PTC | 4 | 4 | 5 | 4 |
| SH0021 | PTC | 4 | 5 | 4 | 4 |
| AH0126 | PTC | 4 | 5 | 4 | 4 |

| | | | | | |
|---|---|---|---|---|---|
| SH0141 | PTC | 4 | 5 | 4 | 4 |
| AH0116 | PTC | 4 | 5 | 4 | 4 |
| AH0051 | BTN | 3 | 4 | 4 | 4 |
| AH0205 | PTC | 4 | 5 | 4 | 4 |
| AH0033 | PTC | 4 | 5 | 4 | 4 |
| AH0102 | PTC | 5 | 4 | 5 | 5 |
| JS0001 | No Surgery | 1 | 1 | 2 | 1 |
| JS0003 | No Surgery | 1 | 1 | 1 | 1 |
| JS0009 | No Surgery | 1 | 2 | 1 | 1 |
| JS0026 | No Surgery | 1 | 2 | 2 | 2 |
| JS0031 | No Surgery | 2 | 1 | 1 | 1 |
| JS0032 | No Surgery | 1 | 1 | 1 | 1 |
| JS0060 | No Surgery | 2 | 2 | 2 | 2 |
| JS0061 | No Surgery | 1 | 1 | 2 | 1 |
| JS0101 | No Surgery | 1 | 2 | 2 | 2 |
| JS0142 | No Surgery | 2 | 2 | 1 | 2 |
| JS0253 | No Surgery | 1 | 1 | 1 | 1 |
| JS0279 | No Surgery | 2 | 1 | 2 | 2 |
| JS0284 | No Surgery | 1 | 2 | 1 | 1 |
| JS0301 | No Surgery | 1 | 1 | 1 | 1 |
| JS0894 | No Surgery | 1 | 1 | 2 | 1 |
| JS0017 | No Surgery | 2 | 2 | 1 | 2 |
| JS0018 | No Surgery | 2 | 2 | 2 | 2 |
| JS0019 | No Surgery | 2 | 2 | 1 | 2 |
| JS0028 | No Surgery | 2 | 2 | 1 | 2 |
| JS0029 | No Surgery | 2 | 3 | 2 | 2 |
| JS0035 | No Surgery | 2 | 2 | 1 | 2 |
| JS0064 | No Surgery | 3 | 2 | 2 | 2 |
| JS0069 | No Surgery | 2 | 2 | 2 | 2 |
| JS0070 | No Surgery | 3 | 2 | 2 | 2 |
| JS0072 | No Surgery | 2 | 2 | 1 | 2 |
| JS0074 | No Surgery | 2 | 1 | 2 | 2 |
| JS0078 | No Surgery | 1 | 2 | 2 | 2 |
| JS0079 | No Surgery | 1 | 2 | 2 | 2 |
| JS0086 | No Surgery | 2 | 2 | 2 | 2 |
| JS0088 | No Surgery | 2 | 3 | 2 | 2 |
| JS0089 | No Surgery | 2 | 2 | 2 | 2 |
| JS0091 | No Surgery | 2 | 2 | 2 | 2 |
| JS0106 | No Surgery | 2 | 2 | 3 | 2 |
| JS0107 | No Surgery | 2 | 2 | 2 | 2 |
| JS0108 | No Surgery | 1 | 2 | 2 | 2 |
| JS0110 | No Surgery | 2 | 2 | 3 | 2 |
| JS0112 | No Surgery | 2 | 2 | 3 | 2 |
| JS0116 | No Surgery | 2 | 2 | 3 | 2 |
| JS0118 | No Surgery | 2 | 3 | 2 | 2 |

| | | | | | |
|---|---|---|---|---|---|
| JS0119 | No Surgery | 1 | 2 | 2 | 2 |
| JS0123 | No Surgery | 2 | 2 | 2 | 2 |
| JS0124 | No Surgery | 2 | 1 | 2 | 2 |
| JS0125 | No Surgery | 2 | 2 | 2 | 2 |
| JS0127 | No Surgery | 2 | 1 | 2 | 2 |
| JS0128 | No Surgery | 1 | 2 | 2 | 2 |
| JS0129 | No Surgery | 1 | 2 | 2 | 2 |
| JS0132 | No Surgery | 1 | 2 | 2 | 2 |
| JS0137 | No Surgery | 1 | 2 | 2 | 2 |
| JS0138 | No Surgery | 2 | 2 | 2 | 2 |
| JS0145 | No Surgery | 2 | 2 | 3 | 2 |
| JS0146 | No Surgery | 2 | 2 | 3 | 2 |
| JS0150 | No Surgery | 2 | 2 | 3 | 2 |
| JS0153 | No Surgery | 2 | 3 | 2 | 2 |
| JS0167 | No Surgery | 2 | 2 | 2 | 2 |
| JS0168 | No Surgery | 1 | 2 | 2 | 2 |
| JS0169 | No Surgery | 2 | 2 | 2 | 2 |
| JS0170 | No Surgery | 2 | 3 | 2 | 2 |
| JS0171 | No Surgery | 2 | 2 | 3 | 2 |
| JS0173 | No Surgery | 2 | 2 | 3 | 2 |
| JS0174 | No Surgery | 1 | 2 | 2 | 2 |
| JS0178 | No Surgery | 1 | 2 | 2 | 2 |
| JS0180 | No Surgery | 1 | 2 | 2 | 2 |
| JS0181 | No Surgery | 2 | 2 | 3 | 2 |
| JS0189 | No Surgery | 2 | 2 | 2 | 2 |
| JS0190 | No Surgery | 2 | 2 | 2 | 2 |
| JS0191 | No Surgery | 2 | 2 | 2 | 2 |
| JS0193 | No Surgery | 2 | 3 | 2 | 2 |
| JS0195 | No Surgery | 2 | 3 | 2 | 2 |
| JS0196 | No Surgery | 2 | 3 | 2 | 2 |
| JS0198 | No Surgery | 2 | 2 | 2 | 2 |
| JS0205 | No Surgery | 2 | 1 | 2 | 2 |
| JS0206 | No Surgery | 2 | 2 | 2 | 2 |
| JS0208 | No Surgery | 1 | 2 | 2 | 2 |
| JS0209 | No Surgery | 2 | 1 | 2 | 2 |
| JS0213 | No Surgery | 1 | 2 | 2 | 2 |
| JS0214 | No Surgery | 2 | 3 | 2 | 2 |
| JS0216 | No Surgery | 2 | 2 | 3 | 2 |
| JS0219 | No Surgery | 2 | 2 | 3 | 2 |
| JS0221 | No Surgery | 1 | 2 | 2 | 2 |
| JS0222 | No Surgery | 1 | 1 | 2 | 1 |
| JS0226 | No Surgery | 1 | 2 | 2 | 2 |
| JS0227 | No Surgery | 1 | 1 | 1 | 1 |
| JS0233 | No Surgery | 2 | 1 | 2 | 2 |
| JS0246 | No Surgery | 2 | 2 | 3 | 2 |

| | | | | | |
|---|---|---|---|---|---|
| JS0255 | No Surgery | 2 | 2 | 2 | 2 |
| JS0259 | No Surgery | 1 | 2 | 2 | 2 |
| JS0261 | No Surgery | 2 | 2 | 2 | 2 |
| JS0262 | No Surgery | 2 | 2 | 1 | 2 |
| JS0266 | No Surgery | 2 | 2 | 2 | 2 |
| JS0267 | No Surgery | 2 | 2 | 3 | 2 |
| JS0268 | No Surgery | 1 | 2 | 2 | 2 |
| JS0269 | No Surgery | 1 | 2 | 2 | 2 |
| JS0270 | No Surgery | 2 | 1 | 2 | 2 |
| JS0272 | No Surgery | 2 | 2 | 2 | 2 |
| JS0273 | No Surgery | 2 | 3 | 2 | 2 |
| JS0274 | No Surgery | 2 | 3 | 2 | 2 |
| JS0275 | No Surgery | 2 | 2 | 3 | 2 |
| JS0277 | No Surgery | 1 | 1 | 2 | 1 |
| JS0278 | No Surgery | 2 | 2 | 3 | 2 |
| JS0286 | No Surgery | 1 | 2 | 2 | 2 |
| JS0294 | No Surgery | 1 | 2 | 2 | 2 |
| JS0309 | No Surgery | 2 | 1 | 2 | 2 |
| JS0311 | No Surgery | 2 | 1 | 2 | 2 |
| JS0315 | No Surgery | 1 | 2 | 2 | 2 |
| JS0317 | No Surgery | 2 | 1 | 2 | 2 |
| JS0319 | No Surgery | 2 | 1 | 2 | 2 |
| JS0321 | No Surgery | 2 | 2 | 2 | 2 |
| JS0327 | No Surgery | 2 | 2 | 2 | 2 |
| JS0330 | No Surgery | 1 | 1 | 2 | 1 |
| JS0332 | No Surgery | 2 | 2 | 3 | 2 |
| JS0333 | No Surgery | 2 | 2 | 3 | 2 |
| JS0337 | No Surgery | 1 | 2 | 2 | 2 |
| JS0339 | No Surgery | 1 | 2 | 2 | 2 |
| JS0340 | No Surgery | 2 | 2 | 2 | 2 |
| JS0004 | No Surgery | 2 | 2 | 3 | 2 |
| JS0005 | No Surgery | 2 | 3 | 2 | 2 |
| JS0006 | No Surgery | 2 | 2 | 2 | 2 |
| JS0007 | No Surgery | 2 | 2 | 2 | 2 |
| JS0008 | No Surgery | 2 | 1 | 2 | 2 |
| JS0010 | No Surgery | 1 | 2 | 1 | 1 |
| JS0011 | No Surgery | 1 | 2 | 2 | 2 |
| JS0014 | No Surgery | 1 | 1 | 2 | 1 |
| JS0015 | No Surgery | 2 | 2 | 3 | 2 |
| JS0016 | No Surgery | 1 | 2 | 2 | 2 |
| JS0020 | No Surgery | 2 | 1 | 2 | 2 |
| JS0022 | No Surgery | 2 | 1 | 1 | 1 |
| JS0023 | No Surgery | 2 | 2 | 2 | 2 |
| JS0024 | No Surgery | 2 | 3 | 2 | 2 |
| JS0025 | No Surgery | 2 | 3 | 2 | 2 |

| | | | | | |
|---|---|---|---|---|---|
| JS0030 | No Surgery | 1 | 1 | 1 | 1 |
| JS0033 | No Surgery | 2 | 3 | 2 | 2 |
| JS0034 | No Surgery | 1 | 2 | 2 | 2 |
| JS0037 | No Surgery | 1 | 2 | 2 | 2 |
| JS0041 | No Surgery | 1 | 1 | 2 | 1 |
| JS0042 | No Surgery | 2 | 2 | 2 | 2 |
| JS0043 | No Surgery | 1 | 2 | 2 | 2 |
| JS0045 | No Surgery | 2 | 2 | 3 | 2 |
| JS0046 | No Surgery | 2 | 2 | 3 | 2 |
| JS0047 | No Surgery | 2 | 2 | 3 | 2 |
| JS0048 | No Surgery | 2 | 2 | 2 | 2 |
| JS0049 | No Surgery | 2 | 1 | 1 | 1 |
| JS0050 | No Surgery | 2 | 2 | 2 | 2 |
| JS0052 | No Surgery | 1 | 1 | 1 | 1 |
| JS0053 | No Surgery | 2 | 1 | 2 | 2 |
| JS0059 | No Surgery | 1 | 2 | 2 | 2 |
| JS0065 | No Surgery | 1 | 2 | 2 | 2 |
| JS0071 | No Surgery | 2 | 2 | 2 | 2 |
| JS0073 | No Surgery | 1 | 1 | 2 | 1 |
| JS0075 | No Surgery | 2 | 1 | 2 | 2 |
| JS0077 | No Surgery | 2 | 1 | 1 | 1 |
| JS0080 | No Surgery | 2 | 2 | 2 | 2 |
| JS0081 | No Surgery | 2 | 3 | 2 | 2 |
| JS0082 | No Surgery | 1 | 2 | 1 | 1 |
| JS0083 | No Surgery | 2 | 2 | 2 | 2 |
| JS0084 | No Surgery | 1 | 1 | 1 | 1 |
| JS0087 | No Surgery | 2 | 1 | 2 | 2 |
| JS0092 | No Surgery | 1 | 2 | 2 | 2 |
| JS0093 | No Surgery | 1 | 1 | 2 | 1 |
| JS0095 | No Surgery | 2 | 2 | 2 | 2 |
| JS0097 | No Surgery | 1 | 1 | 1 | 1 |
| JS0098 | No Surgery | 1 | 2 | 1 | 1 |
| JS0100 | No Surgery | 1 | 1 | 2 | 1 |
| JS0102 | No Surgery | 2 | 2 | 3 | 2 |
| JS0104 | No Surgery | 2 | 2 | 3 | 2 |
| JS0105 | No Surgery | 2 | 2 | 3 | 2 |
| JS0109 | No Surgery | 1 | 2 | 2 | 2 |
| JS0111 | No Surgery | 2 | 2 | 3 | 2 |
| JS0113 | No Surgery | 2 | 2 | 3 | 2 |
| JS0114 | No Surgery | 1 | 2 | 2 | 2 |
| JS0115 | No Surgery | 2 | 2 | 2 | 2 |
| JS0117 | No Surgery | 2 | 1 | 2 | 2 |
| JS0120 | No Surgery | 2 | 1 | 2 | 2 |
| JS0121 | No Surgery | 2 | 1 | 2 | 2 |
| JS0122 | No Surgery | 1 | 1 | 1 | 1 |

| | | | | | |
|---|---|---|---|---|---|
| JS0126 | No Surgery | 2 | 1 | 2 | 2 |
| JS0130 | No Surgery | 1 | 1 | 2 | 1 |
| JS0131 | No Surgery | 2 | 2 | 1 | 2 |
| JS0133 | No Surgery | 2 | 2 | 2 | 2 |
| JS0134 | No Surgery | 2 | 2 | 3 | 2 |
| JS0135 | No Surgery | 1 | 1 | 1 | 1 |
| JS0136 | No Surgery | 1 | 2 | 1 | 1 |
| JS0139 | No Surgery | 2 | 2 | 2 | 2 |
| JS0140 | No Surgery | 3 | 2 | 2 | 2 |
| JS0141 | No Surgery | 1 | 2 | 1 | 1 |
| JS0143 | No Surgery | 1 | 1 | 1 | 1 |
| JS0144 | No Surgery | 2 | 1 | 2 | 2 |
| JS0147 | No Surgery | 1 | 1 | 1 | 1 |
| JS0148 | No Surgery | 1 | 1 | 2 | 1 |
| JS0154 | No Surgery | 2 | 1 | 2 | 2 |
| JS0155 | No Surgery | 1 | 1 | 1 | 1 |
| JS0172 | No Surgery | 3 | 2 | 2 | 2 |
| JS0175 | No Surgery | 2 | 1 | 1 | 1 |
| JS0176 | No Surgery | 1 | 1 | 1 | 1 |
| JS0177 | No Surgery | 2 | 2 | 1 | 2 |
| JS0179 | No Surgery | 2 | 2 | 2 | 2 |
| JS0182 | No Surgery | 1 | 2 | 2 | 2 |
| JS0183 | No Surgery | 1 | 2 | 2 | 2 |
| JS0184 | No Surgery | 2 | 1 | 2 | 2 |
| JS0185 | No Surgery | 2 | 2 | 2 | 2 |
| JS0186 | No Surgery | 2 | 1 | 2 | 2 |
| JS0187 | No Surgery | 1 | 2 | 2 | 2 |
| JS0188 | No Surgery | 1 | 1 | 1 | 1 |
| JS0192 | No Surgery | 1 | 2 | 1 | 1 |
| JS0197 | No Surgery | 1 | 1 | 1 | 1 |
| JS0199 | No Surgery | 1 | 2 | 1 | 1 |
| JS0200 | No Surgery | 1 | 2 | 1 | 1 |
| JS0201 | No Surgery | 1 | 2 | 1 | 1 |
| JS0202 | No Surgery | 1 | 1 | 2 | 1 |
| JS0203 | No Surgery | 2 | 1 | 2 | 2 |
| JS0204 | No Surgery | 2 | 1 | 1 | 1 |
| JS0207 | No Surgery | 2 | 2 | 2 | 2 |
| JS0210 | No Surgery | 2 | 3 | 2 | 2 |
| JS0211 | No Surgery | 1 | 1 | 1 | 1 |
| JS0212 | No Surgery | 2 | 2 | 1 | 2 |
| JS0215 | No Surgery | 1 | 1 | 2 | 1 |
| JS0217 | No Surgery | 1 | 1 | 1 | 1 |
| JS0218 | No Surgery | 2 | 1 | 1 | 1 |
| JS0220 | No Surgery | 2 | 3 | 2 | 2 |
| JS0224 | No Surgery | 1 | 1 | 1 | 1 |

| | | | | | |
|---|---|---|---|---|---|
| JS0225 | No Surgery | 1 | 2 | 1 | 1 |
| JS0228 | No Surgery | 1 | 2 | 1 | 1 |
| JS0229 | No Surgery | 1 | 2 | 1 | 1 |
| JS0230 | No Surgery | 1 | 1 | 1 | 1 |
| JS0231 | No Surgery | 2 | 1 | 1 | 1 |
| JS0232 | No Surgery | 2 | 1 | 1 | 1 |
| JS0234 | No Surgery | 2 | 1 | 1 | 1 |
| JS0235 | No Surgery | 1 | 1 | 1 | 1 |
| JS0238 | No Surgery | 2 | 3 | 2 | 2 |
| JS0239 | No Surgery | 1 | 2 | 1 | 1 |
| JS0240 | No Surgery | 1 | 1 | 1 | 1 |
| JS0241 | No Surgery | 1 | 2 | 1 | 1 |
| JS0244 | No Surgery | 1 | 2 | 1 | 1 |
| JS0254 | No Surgery | 1 | 2 | 2 | 2 |
| JS0257 | No Surgery | 1 | 1 | 2 | 1 |
| JS0258 | No Surgery | 1 | 2 | 2 | 2 |
| JS0263 | No Surgery | 1 | 1 | 2 | 1 |
| JS0264 | No Surgery | 1 | 2 | 2 | 2 |
| JS0265 | No Surgery | 1 | 1 | 1 | 1 |
| JS0271 | No Surgery | 2 | 1 | 2 | 2 |
| JS0276 | No Surgery | 2 | 2 | 3 | 2 |
| JS0280 | No Surgery | 2 | 2 | 3 | 2 |
| JS0281 | No Surgery | 2 | 2 | 2 | 2 |
| JS0283 | No Surgery | 2 | 2 | 3 | 2 |
| JS0291 | No Surgery | 1 | 2 | 2 | 2 |
| JS0292 | No Surgery | 1 | 1 | 2 | 1 |
| JS0295 | No Surgery | 2 | 2 | 1 | 2 |
| JS0298 | No Surgery | 2 | 2 | 2 | 2 |
| JS0300 | No Surgery | 2 | 2 | 3 | 2 |
| JS0302 | No Surgery | 2 | 3 | 2 | 2 |
| JS0304 | No Surgery | 2 | 3 | 2 | 2 |
| JS0305 | No Surgery | 2 | 2 | 2 | 2 |
| JS0306 | No Surgery | 2 | 1 | 2 | 2 |
| JS0307 | No Surgery | 1 | 1 | 2 | 1 |
| JS0310 | No Surgery | 1 | 2 | 2 | 2 |
| JS0312 | No Surgery | 2 | 2 | 2 | 2 |
| JS0313 | No Surgery | 1 | 2 | 2 | 2 |
| JS0314 | No Surgery | 1 | 2 | 2 | 2 |
| JS0322 | No Surgery | 2 | 2 | 2 | 2 |
| JS0323 | No Surgery | 1 | 2 | 2 | 2 |
| JS0324 | No Surgery | 2 | 2 | 3 | 2 |
| JS0325 | No Surgery | 2 | 2 | 3 | 2 |
| JS0326 | No Surgery | 1 | 2 | 2 | 2 |
| JS0331 | No Surgery | 2 | 2 | 2 | 2 |
| JS0336 | No Surgery | 1 | 1 | 2 | 1 |

| | | | | | |
|---|---|---|---|---|---|
| JS0338 | No Surgery | 1 | 2 | 2 | 2 |
| JS0341 | No Surgery | 1 | 1 | 1 | 1 |
| JS0342 | No Surgery | 1 | 1 | 1 | 1 |
| JS0347 | No Surgery | 1 | 2 | 2 | 2 |
| JS0348 | No Surgery | 1 | 1 | 2 | 1 |
| JS0350 | No Surgery | 1 | 1 | 1 | 1 |
| JS0893 | No Surgery | 1 | 2 | 2 | 2 |
| AH0089 | No Surgery | 1 | 1 | 1 | 1 |
| JS0013 | No Surgery | 0 | 1 | 1 | 1 |
| JS0021 | No Surgery | 1 | 2 | 1 | 1 |
| JS0027 | No Surgery | 1 | 1 | 1 | 1 |
| JS0038 | No Surgery | 1 | 2 | 1 | 1 |
| JS0040 | No Surgery | 1 | 1 | 1 | 1 |
| JS0044 | No Surgery | 1 | 1 | 2 | 1 |
| JS0051 | No Surgery | 2 | 2 | 1 | 2 |
| JS0054 | No Surgery | 2 | 2 | 2 | 2 |
| JS0055 | No Surgery | 2 | 3 | 2 | 2 |
| JS0056 | No Surgery | 2 | 2 | 2 | 2 |
| JS0062 | No Surgery | 2 | 2 | 2 | 2 |
| JS0099 | No Surgery | 1 | 2 | 2 | 2 |
| JS0194 | No Surgery | 2 | 2 | 1 | 2 |
| JS0236 | No Surgery | 2 | 2 | 2 | 2 |
| JS0247 | No Surgery | 1 | 2 | 1 | 1 |
| JS0251 | No Surgery | 1 | 1 | 1 | 1 |
| JS0252 | No Surgery | 2 | 2 | 1 | 2 |
| JS0287 | No Surgery | 2 | 2 | 2 | 2 |
| JS0288 | No Surgery | 2 | 2 | 2 | 2 |
| JS0289 | No Surgery | 2 | 2 | 2 | 2 |
| JS0290 | No Surgery | 2 | 2 | 2 | 2 |
| JS0293 | No Surgery | 1 | 2 | 2 | 2 |
| JS0297 | No Surgery | 1 | 2 | 2 | 2 |
| JS0299 | No Surgery | 2 | 2 | 3 | 2 |
| JS0303 | No Surgery | 1 | 2 | 2 | 2 |
| JS0308 | No Surgery | 1 | 2 | 2 | 2 |
| JS0335 | No Surgery | 2 | 3 | 2 | 2 |
| JS0344 | No Surgery | 1 | 2 | 2 | 2 |
| JS0349 | No Surgery | 2 | 2 | 2 | 2 |
| JS0351 | No Surgery | 2 | 3 | 2 | 2 |
| JS0002 | No Surgery | 2 | 2 | 3 | 2 |
| JS0012 | No Surgery | 2 | 2 | 2 | 2 |
| JS4001 | FTA | 2 | 2 | 3 | 2 |
| JS4002 | FTA | 3 | 3 | 2 | 3 |
| JS4003 | TFND | 3 | 4 | 4 | 4 |
| JS4004 | FTA | 3 | 4 | 4 | 4 |
| JS4005 | TFND | 3 | 2 | 3 | 3 |

| | | | | | |
|---|---|---|---|---|---|
| JS4006 | FTA | 2 | 3 | 3 | 3 |
| JS4007 | TFND | 2 | 3 | 3 | 3 |
| JS4008 | FTA | 3 | 3 | 4 | 3 |
| JS4009 | FTA | 4 | 3 | 3 | 3 |
| JS4010 | TFND | 3 | 2 | 2 | 2 |
| JS4011 | TFND | 2 | 2 | 3 | 2 |
| JS4012 | TFND | 3 | 2 | 2 | 2 |
| JS4013 | TFND | 2 | 3 | 2 | 2 |
| JS4014 | FTA | 3 | 2 | 3 | 3 |
| JS4015 | FTA | 3 | 3 | 4 | 3 |
| JS4016 | FTA | 2 | 3 | 2 | 2 |
| JS4017 | TFND | 3 | 3 | 4 | 3 |
| JS4018 | FTA | 3 | 3 | 3 | 3 |
| JS4019 | TFND | 2 | 2 | 3 | 2 |
| JS4020 | FTA | 2 | 2 | 2 | 2 |
| JS4021 | TFND | 2 | 2 | 3 | 2 |
| JS4022 | FTA | 3 | 2 | 2 | 2 |
| JS4023 | FTA | 3 | 4 | 3 | 3 |
| JS4024 | FTA | 4 | 3 | 3 | 3 |
| JS4025 | FTA | 4 | 3 | 3 | 3 |
| JS4026 | FTA | 3 | 3 | 2 | 3 |
| JS4027 | TFND | 2 | 3 | 2 | 2 |
| JS4028 | FTA | 3 | 3 | 2 | 3 |
| JS4029 | TFND | 2 | 2 | 3 | 2 |
| JS4030 | FTA | 4 | 3 | 4 | 4 |
| JS4031 | FTA | 2 | 3 | 2 | 2 |
| JS4032 | FTA | 3 | 2 | 3 | 3 |
| JS4033 | TFND | 4 | 2 | 4 | 4 |
| JS4034 | FTA | 3 | 4 | 3 | 3 |
| JS4035 | FTA | 2 | 3 | 2 | 2 |
| JS4036 | TFND | 3 | 3 | 2 | 3 |
| AH0140 | PTC | 4 | 4 | 3 | 4 |
| AH0268 | PTC | 4 | 3 | 4 | 4 |
| AH0374 | PTC | 4 | 5 | 4 | 4 |
| AH0397 | PTC | 5 | 4 | 4 | 4 |
| AH0408 | PTC | 4 | 4 | 3 | 4 |
| AH0410 | PTC | 4 | 4 | 4 | 4 |
| AH0258 | PTC | 5 | 4 | 4 | 4 |
| AH0381 | PTC | 5 | 4 | 4 | 4 |

### Table S9. Characteristics of Thyroid nodules

| | The model training cohort | | The model validation cohort | | The second validation cohort | |
|---|---|---|---|---|---|---|
| Variables | DTC | BTN | DTC | BTN | DTC | BTN |
| n | 454 | 418 | 195 | 197 | 300 | 99 |
| Tumor Size (mm) | | | | | | |
| Length (IQR) | 8.0 (5.0-14.0) | 17.5 (8.1-31.5) | 7.0 (5.0-10.0) | 20.0 (5.0-40.0) | 4.5 (3.0-9.2) | 22.5 (6.0-26.2) |
| Width (IQR) | 7.0 (5.0-10.0) | 11.5 (6.5-25.0) | 7.0 (5.0-10.0) | 15.0 (4.0-25.0) | 3.8 (2.6-10.0) | 15..0 (4.0-20.0) |
| TNM stage[#] (n) | | | | | | |
| T | 450 | | 144 | | 291 | |
| T1 | 393 (87.3%) | - | 127 (88.2%) | - | 250 (85.9%) | |
| T2 | 32 (7.1%) | - | 6 (4.2%) | - | 17 (5.8%) | |
| T3 | 16 (3.6%) | - | 8 (5.6%) | - | 6 (2.1%) | |
| T4 | 9 (2.0%) | - | 3 (2.1%) | - | 18 (6.2%) | |
| N | | | | | | |
| N0 | 232 (51.6%) | - | 73 (50.7%) | - | 147 (50.5%) | |
| N1 | 212 (47.1%) | - | 71 (49.3%) | - | 143 (49.1%) | |
| NX | 6 (1.3%) | | 0 (0.0%) | | 1 (0.3%) | |
| M | | | | | | |
| M0 | 450 (100.0%) | | 144 (100.0%) | - | 289 (99.7%) | |
| M1 | 0 (0.0%) | - | 0 (0.0%) | | 1 (0.3%) | |
| DTC Subtypes (n) | | | | | | |
| PTC | 435 (95.8%) | - | 187 (95.9) | - | 296 (98.7%) | - |
| PTC with OCA | 2 (0.4%) | - | 2 (1.0%) | - | 0 (0.0%) | - |
| FTC | 11 (2.4%) | - | 5 (2.5%) | - | 3 (1.0%) | - |

| | | | | | | |
|---|---|---|---|---|---|---|
| OCC | 6 (1.3%) | - | 1 (0.5%) | - | 1(0.3%) | - |
| BTN Subtypes* (n) | | | | | | |
| FTA | - | 42 (41.6%) | - | 50 (25.4%) | | 36 (36.4%) |
| TFND | - | 55 (54.5%) | - | 146 (74.1%) | | 63 (63.6%) |
| OCA | - | 2 (2.0%) | - | 0 (0.0%) | | 0 (0.0%) |
| HT | - | 2 (2.0%) | - | 1 (0.5%) | | 0 (0.0%) |
| ACR TI-RADS | | | | | | |
| 1 | 0 (0.0%) | 10 (2.4%) | 0 (0.0%) | 0 (0.0%) | 0 (0.0%) | 0 (0.0%) |
| 2 | 2 (0.4%) | 87 (20.8%) | 0 (0.0%) | 4 (2.0%) | 0 (0.0%) | 2 (2.0%) |
| 3 | 50 (11.0%) | 220 (52.6%) | 21 (10.8%) | 109 (55.3%) | 11 (3.7%) | 49 (49.5%) |
| 4 | 249 (54.8%) | 89 (21.3%) | 119 (61.0%) | 75 (38.1%) | 228 (76.0%) | 47 (47.5%) |
| 5 | 153 (33.7%) | 12 (2.9%) | 55 (28.2%) | 9 (4.6%) | 61 (20.3%) | 1 (1.0%) |
| FNAB[&] | | | | | | |
| II | 9 (3.6%) | 68 (60.7%) | 3 (2.7%) | 105 (56.1%) | 5 (5.1%) | 10 (41.7%) |
| III | 36 (14.5%) | 17 (15.2%) | 15 (13.5%) | 37 (19.8%) | 12 (12.2%) | 7 (29.2%) |
| IV | 45 (18.1%) | 20 (17.9%) | 25 (22.5%) | 22 (11.8%) | 6 (6.1%) | 2 (8.3%) |
| V | 62 (25.0%) | 2 (1.8%) | 25 (22.5%) | 17 (9.1%) | 25 (25.5%) | 4 (16.7%) |
| VI | 96 (38.7%) | 5 (4.5%) | 43 (38.7%) | 6 (3.2%) | 50 (51.0%) | 1 (4.2%) |

DTC, Differentiated thyroid carcinoma
BTN, Benign thyroid nodules
PTC, Papillary thyroid carcinoma
FTC, Follicular thyroid carcinoma
OCC, oncocytic thyroid carcinoma
OCA, oncocytic thyroid adenoma
TFND, Thyroid follicular nodular disease.
FTA, follicular thyroid adenoma.
HT, Hashimoto's thyroiditis.
ACR TI-RADS=American College of Radiology Thyroid Imaging Reporting and Data System
FNAB= Fine needle aspiration biopsy
[&]658 subjects had FNAB results
[#]594 DTC subjects had a record of TNM stage.
*101 subjects in BTN of training group had histopathological diagnosis

**Supplementary Table S10.** Clinical characteristics of healthy volunteers

| Variables | Healthy volunteers |
| --- | --- |
| n | 340 |
| Patients Sex | |
| Female | 145 |
| Male | 195 |
| Patients Age (years) | |
| Median | 38 |
| IQR | 32.0~50.0 |
| BMI (Kg/m$^2$) | |
| Median | 23.7 |
| IQR | 21.4~25.7 |
| TT3 (ng/mL) | |
| Median | ~ |
| IQR | ~ |
| TT4 (μg/dL) | |
| Median | ~ |
| IQR | ~ |
| FT3 (pmol/L) | |
| Median | 4.3 |
| IQR | 3.9~4.9 |
| FT4 (pmol/L) | |
| Median | 12.9 |
| IQR | 12.1~14.0 |
| TSH (μIU/mL) | |
| Median | 1.7 |
| IQR | 1.3~2.5 |
| TgAb (IU/mL) | |
| Median | 1.1 |
| IQR | 0.7~1.8 |

**Supplementary Table S11.** The MRM method parameters for the detection of thyroid hormone metabolites

| No. | Compounds | Precursor ions | Fragment ions | Declust ering potenti | Collision energy (V) | ESI (+/-) |
|---|---|---|---|---|---|---|
| 1 | $T_0$ | 274.0628 | 215.0876 | 50 | 25 | + |
| 2 | 3-$T_1$AM | 355.9024 | 212.0184 | 50 | 25 | + |
| 3 | 3-$T_1$ | 400.0297 | 256.1404 | 50 | 25 | + |
| 4 | 3'-$T_1$ | 400.0297 | 341.0414 | 50 | 30 | + |
| 5 | 3,3'-$T_2$ | 525.7338 | 381.9307 | 50 | 30 | + |
| 6 | 3',5'-$T_2$ | 525.8000 | 466.800 | 50 | 35 | + |
| 7 | 3,5-$T_2$ | 525.8000 | 353.000 | 50 | 45 | + |
| 8 | $T_3$ | 651.7057 | 605.6000 | 50 | 30 | + |
| 9 | r$T_3$ | 651.7057 | 478.9687 | 50 | 50 | + |
| 10 | $T_4$ | 777.6000 | 731.6000 | 50 | 30 | + |

IS: Internal Standard

**Supplementary Table S12.** The quantitative linearity of ten thyroid hormone metabolite compounds

| No. | Compounds | Regression | r | Range (ng/mL) | LLOQ (ng/mL) |
|---|---|---|---|---|---|
| 1 | $T_0$ | y = 290807x +411.33 | 0.9999 | 0.001-2 | 0.001 |
| 2 | 3-$T_1$AM | y = 0.7087x -0.0016 | 0.9987 | 0.002-0.2 | 0.002 |
| 3 | 3-$T_1$ | y = 0.3247x + 0.002 | 0.9999 | 0.01-1 | 0.01 |
| 4 | 3'-$T_1$ | y = 41598x + 22.161 | 0.9988 | 0.002-1 | 0.002 |
| 5 | 3,3'-$T_2$ | y = 54412x - 204.01 | 0.9997 | 0.002-0.2 | 0.002 |
| 6 | 3.5-$T_2$ | y = 9299.7x - 217.79 | 1 | 0.01-0.2 | 0.01 |
| 7 | 3',5'-$T_2$ | y = 85013x - 2336.3 | 0.9991 | 0.01-0.2 | 0.01 |
| 8 | $T_3$ | y = 0.5603x + 0.0086 | 0.9989 | 0.2-10 | 0.2 |
| 9 | r$T_3$ | y = 0.1184x + 0.0018 | 0.999 | 0.1-10 | 0.1 |
| 10 | $T_4$ | y = 1.5286x - 13.867 | 0.999 | 20-200 | 20 |

**Supplementary Table S13.** The inter-day consistency pattern of the LC-MS/MS method

| THMs | Theoretical concentration (pg/mL) | Detected concentration (pg/mL) | | | | | | | | | | | SD (pg/mL) | RSD (%) |
|---|---|---|---|---|---|---|---|---|---|---|---|---|---|---|
| | | Day 1 | Day 2 | Day 3 | Day 4 | Day 5 | Day 6 | Day 7 | Day 8 | Day 9 | Day 10 | Average | | |
| $T_0$ | 10.0 | 10.0 | 10.8 | 10.5 | 9.7 | 9.9 | 11.0 | 10.6 | 11.2 | 10.5 | 10.3 | 10.5 | 0.5 | 4.4 |
| | 100.0 | 97.6 | 99.1 | 108.0 | 109.3 | 100.1 | 105.3 | 101.2 | 96.3 | 96.7 | 101.7 | 101.5 | 4.4 | 4.3 |
| | 1000.0 | 994.1 | 967.5 | 1035.9 | 991.4 | 993.9 | 968.6 | 993.5 | 962.4 | 981.0 | 996.0 | 988.4 | 19.9 | 2.0 |
| $T_1AM$ | 5.0 | 4.9 | 4.6 | 5.0 | 5.3 | 5.0 | 4.8 | 5.4 | 5.0 | 4.8 | 5.1 | 5.0 | 0.2 | 4.4 |
| | 20.0 | 20.5 | 20.7 | 20.4 | 19.3 | 20.2 | 20.1 | 20.1 | 18.2 | 17.8 | 20.4 | 19.8 | 1.0 | 4.9 |
| | 100.0 | 94.1 | 96.0 | 102.9 | 93.0 | 97.7 | 95.4 | 96.2 | 89.5 | 94.0 | 90.9 | 95.0 | 3.5 | 3.7 |
| 3-$T_1$ | 10.0 | 10.3 | 10.9 | 10.2 | 10.5 | 10.0 | 10.7 | 9.4 | 10.5 | 9.4 | 10.0 | 10.2 | 0.5 | 4.7 |
| | 100.0 | 98.9 | 110.2 | 105.0 | 95.8 | 99.1 | 102.6 | 106.3 | 94.9 | 96.5 | 97.9 | 100.7 | 4.8 | 4.8 |
| | 1000.0 | 992.4 | 972.6 | 1017.1 | 1026.1 | 983.0 | 1032.5 | 997.8 | 996.9 | 997.4 | 1007.3 | 1002.3 | 17.8 | 1.8 |
| 3’-$T_1$ | 10.0 | 9.9 | 10.5 | 10.2 | 9.4 | 10.1 | 9.3 | 10.1 | 10.5 | 9.3 | 10.5 | 10.0 | 0.5 | 4.6 |
| | 100.0 | 107.3 | 104.7 | 101.9 | 96.5 | 90.9 | 100.6 | 100.1 | 100.1 | 100.3 | 108.4 | 101.1 | 4.8 | 4.8 |
| | 1000.0 | 988.1 | 998.8 | 1003.7 | 997.4 | 992.0 | 990.5 | 958.9 | 993.4 | 958.3 | 1019.1 | 990.0 | 17.8 | 1.8 |
| 3,3’-$T_2$ | 10.0 | 9.6 | 10.0 | 10.0 | 10.4 | 10.6 | 9.6 | 10.6 | 9.9 | 9.5 | 10.8 | 10.1 | 0.4 | 4.3 |
| | 50.0 | 50.9 | 49.9 | 47.7 | 47.4 | 51.9 | 50.7 | 50.6 | 52.5 | 51.7 | 55.6 | 50.9 | 2.22 | 4.4 |
| | 200.0 | 197.3 | 215.3 | 202.8 | 191.0 | 195.7 | 203.4 | 193.9 | 210.1 | 189.9 | 186.7 | 198.6 | 8.7 | 4.4 |
| 3,5-$T_2$ | 10.0 | 9.8 | 10.1 | 9.7 | 10.0 | 9.5 | 10.1 | 10.3 | 11.0 | 10.7 | 10.5 | 10.2 | 0.4 | 4.3 |
| | 50.0 | 49.0 | 48.2 | 52.2 | 49.0 | 49.5 | 51.5 | 54.1 | 55.3 | 52.5 | 52.4 | 51.4 | 2.2 | 4.3 |
| | 200.0 | 200.9 | 189.4 | 203.0 | 217.6 | 202.6 | 192.9 | 199.4 | 189.8 | 183.0 | 196.8 | 197.5 | 9.1 | 4.6 |

| | | | | | | | | | | | | | | |
|---|---|---|---|---|---|---|---|---|---|---|---|---|---|---|
| | 10.0 | 9.8 | 10.4 | 10.4 | 9.9 | 10.7 | 10.1 | 10.6 | 10.6 | 10.7 | 10.5 | 10.34 | 0.3 | 3.2 |
| 3’,5’-$T_2$ | 50.0 | 54.1 | 47.8 | 48.6 | 48.0 | 53.4 | 55.0 | 50.2 | 46.6 | 48.3 | 46.4 | 49.8 | 3.0 | 6.0 |
| | 200.0 | 191.0 | 200.1 | 213.0 | 189.5 | 194.5 | 195.9 | 192.9 | 200.5 | 188.1 | 181.5 | 194.7 | 8.1 | 4.2 |
| | 200.0 | 194.1 | 209.1 | 209.6 | 198.4 | 207.0 | 196.9 | 192.9 | 214.3 | 206.4 | 228.4 | 205.7 | 10.2 | 4.9 |
| 3,3’,5-$T_3$ | 1000.0 | 1041.8 | 995.9 | 1034.7 | 971.1 | 1005.0 | 992.4 | 1020.9 | 983.3 | 999.9 | 998.8 | 1004.4 | 21.0 | 2.1 |
| | 5000.0 | 4997.3 | 4835.9 | 5266.3 | 5001.7 | 4857.8 | 4906.8 | 4866.3 | 4809.5 | 4952.5 | 4959.4 | 4945.4 | 124.5 | 2.5 |
| | 200.0 | 215.8 | 209.1 | 204.0 | 217.2 | 196.2 | 182.0 | 208.5 | 200.1 | 208.2 | 209.5 | 205.1 | 9.8 | 4.8 |
| 3,3’,5’-$T_3$ | 1000.0 | 1022.3 | 1042.4 | 1088.4 | 1019.4 | 975.4 | 1032.0 | 958.9 | 964.5 | 993.0 | 1030.8 | 1013.3 | 37.7 | 3.7 |
| | 5000.0 | 4877.4 | 4968.9 | 5164.5 | 4593.8 | 4979.3 | 4998.7 | 4766.9 | 4801.2 | 4592.9 | 4475.9 | 4822.0 | 206.3 | 4.3 |
| | Theoretical concentration (ng/mL) | Detected concentration (ng/mL) | | | | | | | | | | | | |
| | 50.0 | 51.8 | 51.6 | 50.0 | 51.4 | 49.2 | 50.2 | 49.1 | 49.9 | 48.8 | 49.4 | 50.2 | 1.1 | 2.1 |
| $T_4$ | 100.0 | 98.6 | 98.7 | 98.8 | 99.7 | 98.7 | 101.4 | 102.0 | 101.2 | 100.7 | 101.5 | 100.1 | 1.3 | 1.3 |
| | 150.0 | 150.0 | 151.8 | 148.1 | 152.6 | 151.6 | 152.1 | 150.0 | 147.6 | 148.1 | 150.1 | 150.2 | 1.7 | 1.1 |

**Supplementary Table S14.** The concentrations of serum THMs from the same DTC patient in different tests: demonstration of the consistency and stability of the test.

| No. | Detected concentration (pg/mL) | | | | | | | | | |
|---|---|---|---|---|---|---|---|---|---|---|
| | $T_0$ | 3-$T_1$AM | 3-$T_1$ | 3'-$T_1$ | 3,3'-$T_2$ | 3,5-$T_2$ | 3',5'-$T_2$ | 3,3',5-$T_3$ | 3,3',5'-$T_3$ | T4 (ng/mL) |
| Test 1 | 44.3 | 11.7 | 7.6 | 2.4 | 7.1 | 4.8 | 2.7 | 472.8 | 147.6 | 68.9 |
| Test 2 | 41.7 | 11.4 | 5.6 | 2.3 | 7.0 | 7.2 | 2.3 | 370.3 | 168.0 | 72.5 |
| Test 3 | 56.7 | 13.7 | 5.4 | 2.1 | 7.3 | 7.8 | 2.5 | 392.3 | 134.8 | 69.4 |
| Test 4 | 54.9 | 9.4 | 6.8 | 2.2 | 6.7 | 6.2 | 2.5 | 394.0 | 131.1 | 62.4 |
| Test 5 | 54.3 | 9.8 | 6.2 | 1.7 | 5.5 | 7.1 | 2.4 | 469.2 | 164.4 | 60.0 |
| Test 6 | 43.8 | 11.8 | 6.4 | 2.3 | 7.1 | 5.8 | 2.0 | 537.2 | 174.5 | 71.6 |
| Test 7 | 53.2 | 11.7 | 7.1 | 2.1 | 9.5 | 4.6 | 2.3 | 494.5 | 177.7 | 68.3 |
| Test 8 | 46.0 | 10.8 | 6.4 | 1.5 | 8.9 | 7.7 | 2.3 | 473.4 | 174.7 | 62.4 |
| Test 9 | 55.0 | 12.4 | 7.3 | 2.2 | 6.8 | 6.6 | 2.5 | 423.4 | 177.4 | 72.5 |
| Test 10 | 55.3 | 13.9 | 7.5 | 2.5 | 7.8 | 5.9 | 2.5 | 469.6 | 145.7 | 77.8 |
| Test 11 | 58.6 | 9.3 | 6.3 | 2.6 | 8.9 | 6.1 | 2.1 | 480.2 | 161.7 | 58.7 |
| Test 12 | 52.3 | 9.4 | 6.0 | 2.2 | 9.3 | 6.2 | 2.6 | 511.8 | 150.1 | 68.7 |
| Test 13 | 57.3 | 12.3 | 7.0 | 2.4 | 7.6 | 6.9 | 2.8 | 521.2 | 154.2 | 60.3 |
| Average | 51.8 | 11.4 | 6.6 | 2.2 | 7.6 | 6.4 | 2.4 | 462.3 | 158.6 | 67.2 |
| SD | 5.5 | 1.5 | 0.7 | 0.3 | 1.2 | 1.0 | 0.2 | 50.2 | 15.4 | 5.7 |
| RSD (%) | 10.7 | 13.2 | 10.1 | 13.4 | 15.4 | 15.1 | 8.9 | 10.9 | 9.7 | 8.4 |

**Supplementary Table S15.** The diagnostic performance of the logistic model training, the model validation, and the second validation of the test of three THMs for thyroid nodules

| | | | |
|---|---|---|---|
| **Performance in the Model Training Cohort (n=872, malignancy rate 52.1%)** | | | |
| Pathological Diagnosis | Malignant | Benign | Test Performance, % (95% CI) |
| Test Positive | 422 | 54 | Sensitivity, 93.0 (90.2 to 95.0%) |
| Test Negative | 32 | 364 | Specificity, 87.1 (83.5 to 90.0%) |
| | | | PPV, 88.7 (85.5 to 91.2%) |
| | | | NPV, 91.9 (88.8 to 94.2%) |
| | | | Accuracy, 90.1 (87.0 to 92.9%) |
| **Performance in the Validation Cohort (n=392, malignant rate 49.7%)** | | | |
| Pathological Diagnosis | Malignant | Benign | Test Performance, % (95% CI) |
| Test Positive | 180 | 1 | Sensitivity, 92.3 (87.7 to 95.3%) |
| Test Negative | 15 | 196 | Specificity, 99.5 (97.2 to 99.9%) |
| | | | PPV, 99.4 (96.9 to 99.9%) |
| | | | NPV, 92.9 (88.6 to 95.6%) |
| | | | Accuracy, 95.9 (90.8 to 100.0%) |
| **Performance in Second Validation Cohort (n=399, malignant rate 75.2%)** | | | |
| Pathological Diagnosis | Malignant | Benign | Test Performance, % (95% CI) |
| Test Positive | 276 | 0 | Sensitivity 92.0, (88.4 to 94.6%) |
| Test Negative | 24 | 99 | Specificity 100.0, (96.3 to 100.0%) |
| | | | PPV 100.0, (98.6 to 100.0%) |
| | | | NPV 80.5, (72.6 to 86.5%) |
| | | | Accuracy 94.0, (89.5 to 97.6%) |
| **Performance in the Combined Two Validation Cohorts (n=791, malignant rate 62.6%)** | | | |
| Pathological Diagnosis | Malignant | Benign | Test Performance, % (95% CI) |
| Test Positive | 456 | 1 | Sensitivity 92.1, (89.4 to 94.2%) |
| Test Negative | 39 | 295 | Specificity 99.7, (98.1 to 99.9%) |
| | | | PPV 99.8, (98.8 to 100.0%) |
| | | | NPV 88.3, (84.4 to 91.3%) |
| | | | Accuracy 94.9, (91.5 to 98.0%) |

**Supplementary Table S16.** Diagnostic performance of the test of serum THMs for thyroid nodules in various Bethesda cytological categories with histopathological confirmation

| **Performance in Bethesda II Nodules (n=138, malignant rate 12.3%)** | | | |
|---|---|---|---|
| Pathological Diagnosis | Malignant | Benign | Test Performance, % (95% CI) |
| Test Positive | 17 | 0 | Sensitivity, 100.0 (81.6 to 100.0%) |
| Test Negative | 0 | 121 | Specificity, 100.0 (96.9 to 100.0%) |
| | | | PPV, 100.0 (81.6 to 100.0%) |
| | | | NPV, 100.0 (96.9 to 100.0%) |
| | | | Accuracy, 100.0 (93.1 to 100.0%) |
| **Performance in Bethesda III Nodules (n=117, malignant rate 53.8%)** | | | |
| Pathological Diagnosis | Malignant | Benign | Test Performance, % (95% CI) |
| Test Positive | 58 | 0 | Sensitivity, 92.0 (82.7 to 96.6%) |
| Test Negative | 5 | 54 | Specificity, 100.0 (93.4 to 100.0%) |
| | | | PPV, 100.0 (93.8to 100.0%) |
| | | | NPV, 91.5 (81.6 to 96.3%) |
| | | | Accuracy, 95.7 (85.7 to 100.0%) |
| **Performance in Bethesda IV Nodules (n=109, malignant rate 69.7%)** | | | |
| Pathological Diagnosis | Malignant | Benign | Test Performance, % (95% CI) |
| Test Positive | 72 | 0 | Sensitivity, 94.7 (87.2 to 97.9%) |
| Test Negative | 4 | 33 | Specificity, 100.0 (89.6 to 100.0%) |
| | | | PPV, 100.0 (94.9 to 100.0%) |
| | | | NPV, 89.2 (75.3 to 95.7%) |
| | | | Accuracy, 96.3 (86.5 to 100.0%) |
| **Performance in Bethesda V Nodules (n=134, malignant rate 83.6%)** | | | |
| Pathological Diagnosis | Malignant | Benign | Test Performance, % (95% CI) |
| Test Positive | 96 | 0 | Sensitivity, 85.7 (78.0 to 91.0%) |
| Test Negative | 16 | 22 | Specificity, 100.0 (89.6 to 100.0%) |
| | | | PPV, 100.0 (96.2 to 100.0%) |
| | | | NPV, 67.3 (53.4 to 78.8%) |
| | | | Accuracy, 89.0 (81.7 to 94.2%) |
| **Performance in Bethesda VI Nodules (n=200, malignant rate 94.5%)** | | | |
| Pathological Diagnosis | Malignant | Benign | Test Performance, % (95% CI) |
| Test Positive | 177 | 0 | Sensitivity, 93.6 (89.2 to 96.3%) |
| Test Negative | 12 | 11 | Specificity, 100.0 (85.1 to 100.0%) |
| | | | PPV, 100.0 (97.9 to 100.0%) |
| | | | NPV, 64.7 (47.9 to 78.5%) |
| | | | Accuracy, 94.3 (89.5 to 97.5%) |
| **Performance in Bethesda III & IV Nodules (n=226, malignant rate 61.5%)** | | | |
| Pathological Diagnosis | Malignant | Benign | Test Performance, % (95% CI) |
| Test Positive | 130 | 0 | Sensitivity, 93.5 (88.2 to 96.6%) |
| Test Negative | 9 | 87 | Specificity, 100.0 (95.8 to 100.0%) |

| | | | |
|---|---|---|---|
| | | | PPV, 100.0 (97.1 to 100.0%) |
| | | | NPV, 90.6 (83.1 to 95.0%) |
| | | | Accuracy, 96.0 (89.2 to 100.0%) |
| **Performance in Bethesda III-V Nodules (n=360, malignant rate 69.7%)** | | | |
| Pathological Diagnosis | Malignant | Benign | Test Performance, % (95% CI) |
| Test Positive | 226 | 0 | Sensitivity, 90.0 (85.7 to 93.1%) |
| Test Negative | 25 | 109 | Specificity, 100.0 (96.6 to 100.0%) |
| | | | PPV, 100.0 (98.3 to 100.0%) |
| | | | NPV, 81.3 (73.9 to 87.0%) |
| | | | Accuracy, 93.1 (88.1 to 97.1%) |
| **Performance in all the Bethesda Categories of Nodules Combined (n=698, malignant rate 65.5%)** | | | |
| Pathological Diagnosis | Malignant | Benign | Test Performance, % (95% CI) |
| Test Positive | 420 | 0 | Sensitivity, 91.9 (89.0 to 94.1%) |
| Test Negative | 37 | 241 | Specificity, 100.0 (98.4 to 100.0%) |
| | | | PPV, 100.0 (99.1 to 100.0%) |
| | | | NPV, 86.7 (82.2 to 90.2%) |
| | | | Accuracy, 94.7 (91.1 to 97.8%) |

**Supplementary Table S17.** The diagnostic performance of the serum THM test in various ACR TI-RADS categories of thyroid nodules in the subjects with histopathologically confirmed diagnosis

| **Performance in ACR-TIRADS 2 Nodules (n=7, malignant rate 14.3%)** | | | |
|---|---|---|---|
| Pathological Diagnosis | Malignant | Benign | Test Performance, % (95% CI) |
| Test Positive | 1 | 0 | Sensitivity, 100.0 (20.7 to 99.2%) |
| Test Negative | 0 | 6 | Specificity, 100.0 (61.0 to 100.0%) |
| | | | PPV, 100.0 (20.7 to 100.0%) |
| | | | NPV, 100.0 (61.0 to 98.8%) |
| | | | Accuracy, 100.0 (57.9 to 100.0%) |
| **Performance in ACR-TIRADS 3 Nodules (n=190, malignant rate 31.6%)** | | | |
| Pathological Diagnosis | Malignant | Benign | Test Performance, % (95% CI) |
| Test Positive | 56 | 0 | Sensitivity, 93.3 (84.1 to 97.3%) |
| Test Negative | 4 | 130 | Specificity, 100.0 (97.1 to 100.0%) |
| | | | PPV, 100.0 (93.6 to 100.0%) |
| | | | NPV, 97.0 (92.6 to 98.8%) |
| | | | Accuracy, 97.9 (90.6 to 100.0%) |
| **Performance in ACR-TIRADS 4 Nodules (n=348, malignant rate 73.3%)** | | | |
| Pathological Diagnosis | Malignant | Benign | Test Performance, % (95% CI) |
| Test Positive | 233 | 0 | Sensitivity, 91.3 (87.3 to 94.2%) |
| Test Negative | 22 | 93 | Specificity, 100.0 (96.0 to100.0%) |
| | | | PPV, 100.0 (98.4 to 100.0%) |
| | | | NPV, 80.9 (72.7 to 87.0%) |
| | | | Accuracy, 93.7 (88.8 to 97.6%) |
| **Performance in ACR-TIRADS 5 Nodules (n=153, malignant rate 92.1%)** | | | |
| Pathological Diagnosis | Malignant | Benign | Test Performance, % (95% CI) |
| Test Positive | 133 | 0 | Sensitivity, 94.3 (89.2 to 97.1%) |
| Test Negative | 8 | 12 | Specificity, 100.0 (75.8 to 100.0%) |
| | | | PPV, 100.0 (97.2 to 100.0%) |
| | | | NPV, 60.0 (38.7 to 78.1%) |
| | | | Accuracy, 94.8 (89.5 to 97.9%) |

**Supplementary Table S18.** Concentrations of $T_0$, 3-$T_1$AM, 3-$T_1$, and $T_4$ in DTC, BTN, and matched adjacent normal tissues.

| No. | Tissue weight (mg) | Concentrations of THMs in Tissues | | | |
|---|---|---|---|---|---|
| | | $T_0$ (ng/g) | 3-$T_1$AM (ng/g) | 3-$T_1$(ng/g) | $T_4$ (μg/g) |
| C1[&] | 102.3 | 36.6 | 166.0 | 36.4 | 46.9 |
| C2 | 100.9 | 91.5 | 122.5 | 46.5 | 161.5 |
| C3 | 100.8 | 280.5 | 465.5 | 45.7 | 92.7 |
| C4 | 100.5 | 249.0 | 595.0 | 35.4 | 207.1 |
| C5 | 100.5 | 148.1 | 530.1 | 35.4 | 108.0 |
| C6 | 103.0 | 203.5 | 830.2 | 45.8 | 98.5 |
| C7 | 100.0 | 241.0 | 358.5 | 26.2 | 60.5 |
| C8 | 99.9 | 158.2 | 181.5 | 48.0 | 62.9 |
| C9 | 100.9 | 223.3 | 645.0 | 36.5 | 63.2 |
| C10 | 102.1 | 340.2 | 530.3 | 45.9 | 62.9 |
| C11 | 100.7 | 149.9 | 475.6 | 46.7 | 42.8 |
| C12 | 99.5 | 492.3 | 830.5 | 35.7 | 123.0 |
| C13 | 13.51 | 126.3 | 190.6 | 36.8 | 67.1 |
| C14 | 10.93 | 224.7 | 556.0 | 26.8 | 47.5 |
| C15 | 25.12 | 316.0 | 508.2 | 25.1 | 133.7 |
| C16 | 19.88 | 324.2 | 277.1 | 28.4 | 223.9 |
| C17 | 20.20 | 364.4 | 297.2 | 46.2 | 96.1 |
| C18 | 14.02 | 105.2 | 654.9 | 33.8 | 147.0 |
| C19 | 14.12 | 163.5 | 112.2 | 29.5 | 41.7 |
| C20 | 15.84 | 280.2 | 1015.6 | 35.9 | 92.6 |
| C21 | 23.06 | 317.8 | 333.1 | 28.1 | 87.7 |
| C22 | 4.73 | 138.0 | 381.9 | 28.8 | 76.6 |
| C23 | 10.21 | 285.2 | 569.4 | 31.2 | 51.1 |
| C24 | 13.30 | 265.6 | 535.2 | 45.3 | 81.6 |
| C25 | 16.05 | 182.3 | 1122.0 | 41.7 | 63.6 |
| C26 | 16.81 | 345.0 | 481.0 | 41.7 | 70.5 |
| C27 | 10.39 | 291.2 | 697.9 | 32.1 | 61.2 |
| C28 | 3.71 | 392.2 | 624.6 | 34.0 | 58.9 |
| C1P[#] | 100.5 | 30.1 | 203.5 | 45.6 | 177.5 |
| C2P | 100.1 | 118.5 | 312.8 | 46.1 | 33.3 |
| C3P | 100.9 | 57.5 | 535.7 | 25.9 | 48.6 |
| C4P | 100.3 | 42.5 | 286.6 | 45.8 | 84.1 |
| C5P | 100.4 | 122.4 | 183.5 | 45.9 | 91.5 |
| C6P | 100.2 | 178.5 | 202.5 | 37.1 | 147.1 |
| C7P | 100.1 | 139.4 | 383.5 | 46.3 | 119.0 |
| C8P | 98.0 | 16.1 | 64.0 | 45.6 | 81.6 |
| C9P | 103.7 | 119.0 | 272.2 | 46.3 | 91.5 |
| C10P | 92.6 | 167.6 | 439.5 | 36.9 | 75.5 |
| C11P | 100.8 | 65.5 | 299.3 | 46.1 | 134.0 |
| C12P | 99.8 | 228.8 | 184.5 | 37.0 | 30.6 |
| C13P | 25.58 | 188.0 | 27.0 | 78.6 | 188.0 |
| C14P | 7.56 | 170.8 | 37.7 | 72.4 | 70.8 |

| | | | | | |
|---|---|---|---|---|---|
| C15P | 13.88 | 216.7 | 32.5 | 64.2 | 116.7 |
| C16P | 16.10 | 117.7 | 43.6 | 132.6 | 117.7 |
| C17P | 143.3 | 127.9 | 28.2 | 66.6 | 127.9 |
| C18P | 20.41 | 206.2 | 35.5 | 75.1 | 106.2 |
| C19P | 16.92 | 107.2 | 28.9 | 70.2 | 107.2 |
| C20P | 15.45 | 134.3 | 30.5 | 84.0 | 64.3 |
| C21P | 24.74 | 286.6 | 44.5 | 97.7 | 86.6 |
| C22P | 19.12 | 75.9 | 35.6 | 44.7 | 75.9 |
| C23P | 18.41 | 198.5 | 30.1 | 147.2 | 98.5 |
| C24P | 17.91 | 207.3 | 34.5 | 50.3 | 47.3 |
| C25P | 11.61 | 150.7 | 50.2 | 58.8 | 45.7 |
| C26P | 23.25 | 62.3 | 40.1 | 114.3 | 56.3 |
| C27P | 6.61 | 40.8 | 48.4 | 105.3 | 40.8 |
| C28P | 17.03 | 66.7 | 33.7 | 55.1 | 66.7 |
| B1$^{\alpha}$ | 99.8 | 39.8 | 137.1 | 37.7 | 35.0 |
| B2 | 100.1 | 117.3 | 57.2 | 56.2 | 114.1 |
| B3 | 100.2 | 73.1 | 113.4 | 61.4 | 59.9 |
| B4 | 100.1 | 85.9 | 156.5 | 62.7 | 133.5 |
| B5 | 99.9 | 119.1 | 198.8 | 26.1 | 45.1 |
| B6 | 98.9 | 122.4 | 146.1 | 77.9 | 72.0 |
| B7 | 98.5 | 233.3 | 60.2 | 32.7 | 124.3 |
| B8 | 100.5 | 152.6 | 143.8 | 91.2 | 106.2 |
| B9 | 101.0 | 168.9 | 138.1 | 55.3 | 145.0 |
| B10 | 100.6 | 104.5 | 60.9 | 58.9 | 133.8 |
| B11 | 100.2 | 119.8 | 337.3 | 91.1 | 41.2 |
| B12 | 99.3 | 70.0 | 184.1 | 49.6 | 41.1 |
| B1P$^{\beta}$ | 97.2 | 59.1 | 91.1 | 47.7 | 115.5 |
| B2P | 102.3 | 62.2 | 161.1 | 93.8 | 52.3 |
| B3P | 100.5 | 226.1 | 146.7 | 72.0 | 86.2 |
| B4P | 100.9 | 58.8 | 92.1 | 41.4 | 85.1 |
| B5P | 101.0 | 98.3 | 30.9 | 40.2 | 102.5 |
| B6P | 100.4 | 204.8 | 144.5 | 84.5 | 73.5 |
| B7P | 100.1 | 28.4 | 86.7 | 65.1 | 162 |
| B8P | 99.0 | 37.17 | 68.1 | 43 | 60.5 |
| B9P | 99.2 | 43.8 | 322.5 | 56.2 | 206.5 |
| B10P | 100.2 | 126.1 | 177.5 | 43.1 | 54.5 |
| B11P | 99.3 | 139.3 | 112.4 | 29.8 | 117.5 |
| B12P | 99.5 | 161.2 | 113.2 | 85.9 | 38.9 |

**Footnotes**:

$^{\&}$DTC tissues    $^{\#}$Adjacent normal tissues of DTC

$^{\alpha}$BTN tissues    $^{\beta}$Adjacent normal tissues of BTN

**Supplementary Table S19.** Clinical characteristics of the study subjects

| | The model training cohort | | | The model validation cohort | | | The second validation cohort | | |
|---|---|---|---|---|---|---|---|---|---|
| Variables | DTC | BTN | *P* | DTC | BTN | *P* | DTC | BTN | *P* |
| n | 454 | 418 | | 195 | 197 | | 300 | 99 | |
| Patient Sex | | | <0.001 | | | 0.48 | | | 0.06 |
| Female | 335 | 257 | | 140 | 149 | | 230 | 85 | |
| Male | 119 | 161 | | 55 | 48 | | 70 | 14 | |
| Patient Age (years) | | | 0.003 | | | <0.001 | | | <0.001 |
| Median (IQR) | 47.0 (35.0-55.0) | 49.0 (39.0-56.0) | | 44.0 (34.0-53.0) | 53.0 (45.0-59.0) | | 46.5 (37.5-57.0) | 55.0 (45.0-62.0) | |
| BMI (Kg/m$^2$) | | | 0.41 | | | 0.36 | | | 0.03 |
| Median (IQR) | 23.7 (21.3-26.2) | 23.8 (21.7-25.5) | | 23.8 (22.0-26.7) | 24.7 (22.4-26.8) | | 25.1 (22.8-27.5) | 23.9 (22.1-26.6) | |
| TSH (μIU/mL) | | | 0.94 | | | 0.34 | | | 0.9 |
| Median (IQR) | 1.4 (0.9-2.3) | 1.2 (0.7-2.3) | | 1.1 (0.7-2.3) | 1.0 (0.7-2.7) | | 1.7 (1.1-2.5) | 1.6 (0.8-2.5) | |
| FT4 (pmol/L) | | | 0.004 | | | 0.08 | | | 0.307 |
| Median (IQR) | 12.5 (11.7-13.6) | 12.8 (12.3-14.2) | | 12.6 (12.1-13.4) | 12.9 (11.9-13.9) | | 12.4 (11.7-13.4) | 12.4 (11.5-13.1) | |
| FT3 (pmol/L) | | | <0.001 | | | 0.01 | | | 0.459 |
| Median (IQR) | 4.2 (3.6-4.8) | 4.3 (3.8-4.7) | | 3.7 (3.0-4.1) | 4.2 (4.0-4.5) | | 4.4 (4.1-4.7) | 4.3 (4.0-4.6) | |
| TgAb (IU/mL) | | | 0.14 | | | 0.59 | | | 0.125 |
| Median (IQR) | 3.8 (1.0-33.1) | 2.9 (0.9-15.6) | | 4.1 (1.0-29.4) | 2.9 (0.4-19.9) | | 1.9 (0.8-11.7) | 1.2 (0.8-4.0) | |

DTC= Differentiated thyroid carcinoma; BTN= Benign thyroid nodules

**Supplementary Table S20.** Input Performance by Cohort and Adjusted PPV/NPV

| | Training cohort | Validation cohort 1 | Validation cohort 2 | Whole cohort | Combined validation cohorts |
|---|---|---|---|---|---|
| **TP** | 422 | 180 | 276 | 878 | 456 |
| **FP** | 54 | 1 | 0 | 55 | 1 |
| **FN** | 32 | 15 | 24 | 71 | 39 |
| **TN** | 364 | 196 | 99 | 659 | 295 |
| **Sensitivity (%)** | 92.95 | 92.31 | 92.00 | 92.52 | 92.12 |
| **Specificity (%)** | 87.08 | 99.49 | 100.00 | 92.30 | 99.66 |
| **PPV at 49/100,000** | 0.351 | 8.185 | 100.000 | 0.585 | 11.792 |
| **PPV at 5%** | 27.5 | 90.5 | 100.0 | 38.7 | 93.5 |
| **PPV at 10%** | 44.4 | 95.3 | 100.0 | 57.2 | 96.8 |
| **PPV at 15%** | 55.9 | 97.0 | 100.0 | 67.9 | 98.0 |
| **NPV at 49/100,000** | 99.996 | 99.996 | 99.996 | 99.996 | 99.996 |
| **NPV at 5%** | 99.6 | 99.6 | 99.6 | 99.6 | 99.6 |
| **NPV at 10%** | 99.1 | 99.1 | 99.1 | 99.1 | 99.1 |
| **NPV at 15%** | 98.6 | 98.7 | 98.6 | 98.6 | 98.6 |
